# PRAM: post-hoc retrieval augmentation adapts intensive care prediction models without retraining and can be shared across hospitals

**DOI:** 10.64898/2026.04.03.26350132

**Authors:** Inyong Jeong, Taeyeong Lee, Byeongsu Kim, Jin-Hyun Park, Yeongmin Kim, Hwamin Lee

## Abstract

A hospital given a prediction model it did not build has one resource, its local patients with known outcomes. We compared eight strategies across 116,010 intensive care stays in MIMIC-IV, MIMIC-III and eICU-CRD, for acute kidney injury and 168 hour mortality, and at 67 and 59 eICU-CRD hospitals. Ranking the eight by discrimination and by net benefit named a different winner at 78 percent of hospitals for acute kidney injury and 68 percent for mortality. The criterion can therefore decide whether the installed option helps at the threshold where it acts. Retraining on local data alone cost up to 0.095 AUROC below 1,000 patients. Post-hoc retrieval augmentation (PRAM) leaves the model frozen and adapts by replacing a bank of local records. It cost at most 0.0029 AUROC in the main comparison, improved the calibration of both frozen models where the shift was largest, and its effect on the frozen logistic model varied little between hospitals (heterogeneity 0 percent against 39 to 76 percent for local retraining). Across hospitals it added 0.020 AUROC for mortality and 0.009 for acute kidney injury. Because similarity is computed outside the model, retrieval attached to every base model we tried. The retrieval estimate is a ratio of two sums, so hospitals can exchange 16 bytes per query instead of records. A hospital holding no labelled outcomes can use none of the eight strategies, but querying a network of hospitals that hold them added 0.017 AUROC at 84 to 88 percent of sites. Hospitals should choose an adaptation strategy on calibration, net benefit and the number of labelled local patients they hold, not on discrimination alone.

## Introduction

Prediction models built in one hospital lose accuracy in another.^1–6^ This has been shown many times in intensive care, where a model developed at one centre discriminates less well at the next.^7–9^ That a model which performs best in development can lose that advantage on external data has recently been named a portability paradox.^10^ The usual response is to retrain or fine-tune on data from the receiving hospital.^11,12^ That response assumes staff who can run the training, enough labelled local outcomes, and, for a regulated model, willingness to resubmit it for review.^13–15^ Hospitals without an informatics team meet none of these conditions, and even hospitals that do must decide whether the effort is worth it.

These mechanisms all modify model parameters: domain generalization, domain adaptation, transfer learning and anchor regression.^16–21^ A separate line leaves the model untouched and supplies information at inference time from an external store, so the same mechanism attaches to a model however that model was produced.^22–25^ For tabular data this has been built into network architectures, where the retrieval representation is learned during training and fixed at shipping time, and applied to electronic health records by retrieving concept-level knowledge.^26–28^ Neither attaches to a finished model as a layer whose only moving part is the bank.

These are usually presented as competing methods and compared on discrimination. That is not the question a deploying hospital faces. The hospital has a fixed resource, namely a number of local patients with known outcomes, and must decide what to do with it. Recalibrating the output, retrieving from a bank of those patients, and retraining a model on them all consume the same resource and differ in what they change, what they cost, and what they require in regulatory terms. Whether one dominates the others is an empirical question, and it may have different answers at different local sample sizes.

A model that ranks patients well can still produce probabilities that are systematically too extreme, and a decision made at a fixed probability threshold depends on both,^29,30^ so discrimination is not the right single criterion. Calibration and net benefit are routinely reported for new models but are rarely used to compare adaptation strategies, which leaves open the possibility that the strategy that ranks best also acts worst. The same concern has been raised for continuous acute kidney injury monitoring, where the architecture with the highest area under the curve carried the heaviest alert burden.^31^

A second question follows once the model is already deployed. Collaboration between hospitals is usually arranged at training time: federated learning lets several sites fit one model without moving records, but a site that was not in the consortium cannot join afterwards.^32,33^ Whether anything comparable exists for a layer that sits on top of a finished model is, as far as we are aware, unexamined, and the answer depends on whether the quantity being computed can be assembled from parts held at different sites.

We therefore asked three questions. First, what can a hospital do with a model it did not build, and how do the available strategies compare when discrimination, calibration and net benefit are all measured. Second, what should go into the bank of local patients that retrieval uses. Third, whether retrieval can be run across hospitals in the way federated learning is run across hospitals, and what leaves the building if it is. We ran the comparison across three intensive care databases, two outcomes and local sample sizes from 500 patients to full accumulation, and then repeated it at individual eICU-CRD hospitals. One of the eight strategies is the post-hoc retrieval augmentation module (PRAM), which mixes the frozen model output with outcomes retrieved from the local bank and adapts by bank replacement alone.

## Results

### Cohorts and the size of the shift

The study covered 116,010 first intensive care stays: 33,455 in MIMIC-IV, 14,514 in MIMIC-III and 68,041 in eICU-CRD (Table 1). Acute kidney injury occurred in 14.7, 16.1 and 6.1 percent of stays and 168 hour in-hospital mortality in 4.4, 4.7 and 4.6 percent. All 42 features were present in all three databases (Supplementary Table S1), but nine exceeded 30 percent missingness in MIMIC-III or eICU-CRD and none did in MIMIC-IV.

**Table 1.**
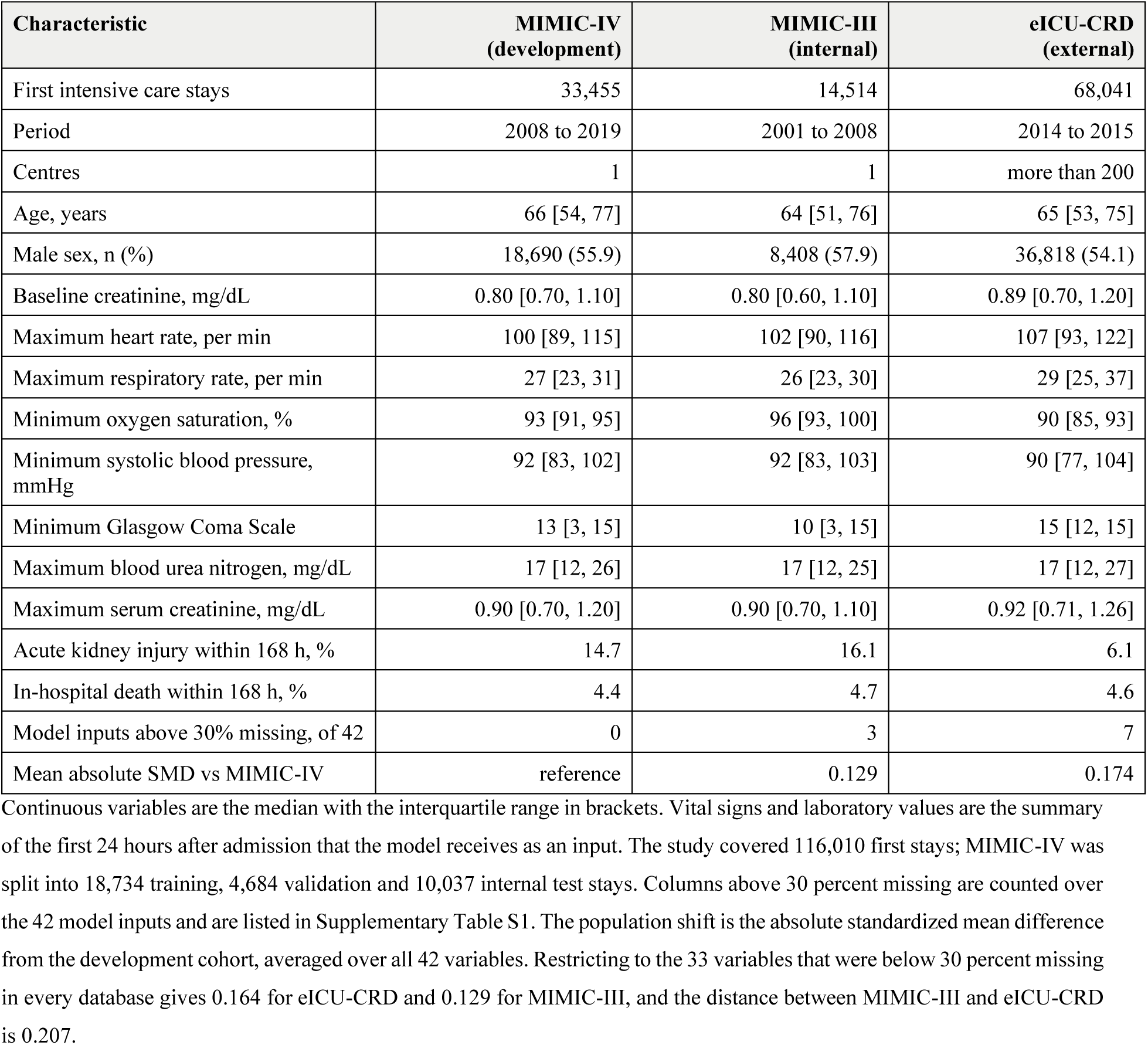
Cohort characteristics, missingness and population shift.

| Characteristic | MIMIC-IV<br>(development) | MIMIC-III<br>(internal) | eICU-CRD<br>(external) |
| --- | --- | --- | --- |
| First intensive care stays | 33,455 | 14,514 | 68,041 |
| Period | 2008 to 2019 | 2001 to 2008 | 2014 to 2015 |
| Centres | 1 | 1 | more than 200 |
| Age, years | 66 [54, 77] | 64 [51, 76] | 65 [53, 75] |
| Male sex, n (%) | 18,690 (55.9) | 8,408 (57.9) | 36,818 (54.1) |
| Baseline creatinine, mg/dL | 0.80 [0.70, 1.10] | 0.80 [0.60, 1.10] | 0.89 [0.70, 1.20] |
| Maximum heart rate, per min | 100 [89, 115] | 102 [90, 116] | 107 [93, 122] |
| Maximum respiratory rate, per min | 27 [23, 31] | 26 [23, 30] | 29 [25, 37] |
| Minimum oxygen saturation, % | 93 [91, 95] | 96 [93, 100] | 90 [85, 93] |
| Minimum systolic blood pressure, mmHg | 92 [83, 102] | 92 [83, 103] | 90 [77, 104] |
| Minimum Glasgow Coma Scale | 13 [3, 15] | 10 [3, 15] | 15 [12, 15] |
| Maximum blood urea nitrogen, mg/dL | 17 [12, 26] | 17 [12, 25] | 17 [12, 27] |
| Maximum serum creatinine, mg/dL | 0.90 [0.70, 1.20] | 0.90 [0.70, 1.10] | 0.92 [0.71, 1.26] |
| Acute kidney injury within 168 h, % | 14.7 | 16.1 | 6.1 |
| In-hospital death within 168 h, % | 4.4 | 4.7 | 4.6 |
| Model inputs above 30% missing, of 42 | 0 | 3 | 7 |
| Mean absolute SMD vs MIMIC-IV | reference | 0.129 | 0.174 |

Standardized mean differences placed eICU-CRD furthest from the development cohort, at 0.174 averaged over all 42 variables. Restricting to the 33 less sparse variables gave 0.164. The same measure between MIMIC-IV and MIMIC-III was 0.129. Values for every variable and every pair of databases are in Supplementary Table S2.

### Eight strategies across local sample size

Figure 1 shows every strategy as a function of bank size in all six cohort and outcome combinations, and Supplementary Table S3 gives the underlying AUROC and AUPRC values.

**Figure 1.**
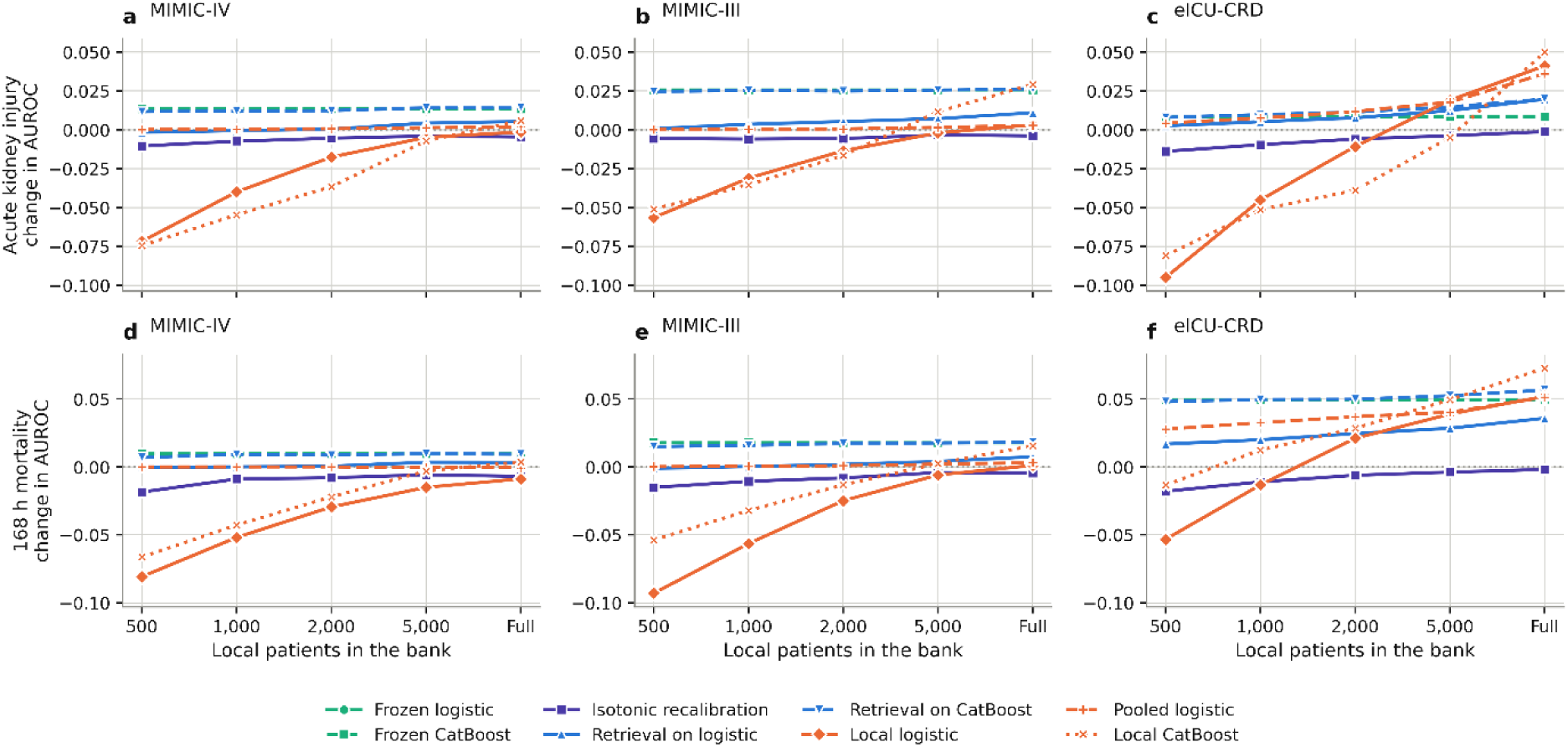
Eight adaptation strategies as a function of local bank size. Each panel is one target cohort and one outcome. The vertical axis is the change in AUROC against the frozen logistic model, so the horizontal line at zero is the do-nothing strategy. The horizontal axis is the number of local patients held, from 500 to full accumulation, on an ordinal scale. Colour distinguishes the family of strategy and line style distinguishes variants within a family. Points are the median over ten independent bank draws, and a single draw at the full pool.

With 500 local patients, a locally fitted logistic model lost 0.095 AUROC against the frozen logistic model in eICU-CRD for acute kidney injury, 0.093 in MIMIC-III for mortality and 0.081 in MIMIC-IV for mortality. A locally fitted CatBoost model lost 0.081 against the same frozen logistic model in eICU-CRD for acute kidney injury at the same size. Local-only retraining overtook the frozen model in eICU-CRD between 1,000 and 2,000 patients for mortality and between 2,000 and 5,000 for acute kidney injury, and in MIMIC-III only at the full pool. In MIMIC-IV it did not overtake at any size we examined, staying 0.0014 below the frozen model for acute kidney injury and 0.0088 below it for mortality with the whole pool in the bank.

Across all 30 combinations of cohort, outcome and bank size, a logistic model fitted on the bank together with the development data was never more than 0.0005 AUROC below the frozen model. With 500 patients in eICU-CRD for mortality it gained 0.028 where the local-only model lost 0.053, and at 1,000 patients it gained 0.033.

At the bank sizes of the main analysis, the largest loss from retrieval on either base model was 0.0029 AUROC, and once the bank reached 2,000 patients no loss exceeded 0.0011. Its gains were concentrated where the shift was largest: on the frozen logistic model it added 0.020 for acute kidney injury and 0.036 for mortality in eICU-CRD at the full bank, against 0.005 and 0.003 in MIMIC-IV. Isotonic recalibration cannot change the ranking of patients, and it lowered AUROC slightly at every bank size, by 0.001 to 0.019, through the ties that its step function introduces.

The dose response between bank size and retrieval performance was significant in every setting (Table 2). AUROC rose by 0.0021 to 0.0130 for each tenfold increase in bank size, with all six p values below 0.0001. Single bank testing was more conservative than the pooled analysis: after correction, improvement over the frozen model survived only for mortality in eICU-CRD at the full bank (q = 0.015). The proportion of individual draws reaching significance was 87 percent for mortality in eICU-CRD at 2,000 patients and 100 percent at 5,000, and never rose above zero for acute kidney injury in MIMIC-IV. The proportion at every bank size is in Supplementary Table S4.

**Table 2.** Dose-response between bank size and retrieval performance, and single-bank significance.

| Outcome | Target | Slope per tenfold increase | 95% CI | p | q at the full bank |
| --- | --- | --- | --- | --- | --- |
| Acute kidney injury | MIMIC-IV | +0.0025 | 0.0022 to 0.0029 | <0.0001 | 0.631 |
| Acute kidney injury | MIMIC-III | +0.0045 | 0.0041 to 0.0048 | <0.0001 | 0.161 |
| Acute kidney injury | eICU-CRD | +0.0061 | 0.0056 to 0.0066 | <0.0001 | 0.129 |
| 168 h mortality | MIMIC-IV | +0.0021 | 0.0017 to 0.0025 | <0.0001 | 0.829 |
| 168 h mortality | MIMIC-III | +0.0041 | 0.0036 to 0.0047 | <0.0001 | 0.701 |
| 168 h mortality | eICU-CRD | +0.0130 | 0.0125 to 0.0136 | <0.0001 | 0.015 |

The slope comes from a linear mixed model of AUROC on log10 bank size with a fold-level random intercept. The last column is the Benjamini-Hochberg corrected p value of a paired DeLong test at the full bank, computed on single bank draws rather than on averaged predictions. The proportion of individual bank draws reaching significance was 87 percent at 2,000 patients and 100 percent at 5,000 for mortality in eICU-CRD, and zero at every size for acute kidney injury in MIMIC-IV.

Local logistic retraining overtook retrieval on the frozen logistic model at about 2,800 patients for mortality and about 4,200 for acute kidney injury in eICU-CRD. In MIMIC-III and MIMIC-IV it had not overtaken retrieval at the largest bank we could form, which was the whole pool of 11,600 and 8,000 patients.

### Discrimination and clinical utility rank the strategies differently

Table 3 gives discrimination, calibration and net benefit for all eight strategies at the full bank in eICU-CRD, the cohort with the largest shift. Net benefit across the whole threshold range is shown in Supplementary Figure S1, and the calibration and net benefit values for MIMIC-IV and MIMIC-III are in Supplementary Table S5, with the AUROC values in Supplementary Table S3.

**Table 3.** Discrimination, calibration and net benefit for the eight strategies at the full bank in eICU-CRD, the cohort with the largest population shift.

| Strategy | AKI AUROC | AKI slope | AKI net benefit | Mortality AUROC | Mortality slope | Mortality net benefit |
| --- | --- | --- | --- | --- | --- | --- |
| Frozen logistic | 0.690 | 0.83 | -0.0015 | 0.774 | 0.41 | -0.0069 |
| Frozen CatBoost | 0.699 | 0.84 | -0.0059 | 0.824 | 0.75 | +0.0013 |
| Isotonic recalibration | 0.689 | 0.98 | +0.0015 | 0.773 | 0.98 | -0.0003 |
| Retrieval on logistic | 0.710 | 1.10 | +0.0018 | 0.810 | 0.78 | -0.0014 |
| Retrieval on CatBoost | 0.710 | 1.01 | -0.0014 | 0.831 | 0.91 | +0.0034 |
| Local logistic | 0.732 | 0.99 | +0.0045 | 0.826 | 0.99 | +0.0047 |
| Pooled logistic | 0.726 | 1.05 | +0.0042 | 0.826 | 0.96 | +0.0045 |
| Local CatBoost | 0.740 | 1.13 | +0.0022 | 0.847 | 1.05 | +0.0067 |

Values are the median over bank draws. A calibration slope of 1.00 is ideal. Net benefit is computed at a threshold probability of 20 percent, and a value below zero means the strategy is worse than treating nobody at that threshold. Expected calibration error for mortality fell from 0.043 to 0.023 under retrieval on the logistic model and from 0.014 to 0.008 under retrieval on CatBoost, and isotonic recalibration reached 0.001. Retrieval placed on top of the local logistic model reached 0.736 for acute kidney injury and 0.833 for mortality, the highest of any strategy built on a logistic model.

For acute kidney injury, the frozen CatBoost model had higher AUROC than the frozen logistic model, 0.699 against 0.690, and the worst net benefit of all eight strategies at a 20 percent threshold, -0.0059 against -0.0015. Both were worse than treating nobody at that threshold. Isotonic recalibration had the lowest AUROC of any strategy at 0.689 and a positive net benefit of +0.0015. Retrieval on the frozen logistic model gained 0.020 AUROC and turned net benefit positive at +0.0018. The three retraining variants led on both measures, and their internal ordering also differed, with local CatBoost first on AUROC at 0.740 and local logistic first on net benefit at +0.0045.

For mortality, frozen CatBoost led the frozen logistic model by 0.049 AUROC and by 0.008 net benefit at a 20 percent threshold, and local CatBoost led on both measures. Isotonic recalibration again produced the lowest AUROC of any strategy, at 0.773, and moved net benefit from -0.0069 to -0.0003. Here the two rankings agreed on which strategy was best, but not on which was worst.

In eICU-CRD for mortality the frozen logistic model had a calibration slope of 0.41, indicating that its probabilities were far too extreme, and the frozen CatBoost model was also miscalibrated at 0.75. Every strategy that touched the local outcomes improved the slope. Retrieval raised it to 0.78 on the logistic model and to 0.91 on CatBoost, isotonic recalibration reached 0.98, and pooled retraining reached 0.96. Expected calibration error followed the same ordering, falling from 0.043 to 0.023 under retrieval on the logistic model and from 0.014 to 0.008 under retrieval on CatBoost. Isotonic recalibration gave the lowest value of any strategy at 0.001. Supplementary Table S6 repeats the discrimination and calibration comparison for the frozen logistic model and retrieval on it in four age bands; net benefit was not computed within bands.

### What to put in the bank

Balancing outcome prevalence was the strongest of the six bank rules (Figure 2, values in Supplementary Table S7). For acute kidney injury in eICU-CRD a balanced bank of 1,000 patients gained 0.019 AUROC, more than a random bank of 5,000, which gained 0.018. For mortality a balanced bank of 300 gained 0.026 against 0.019 for a random bank of 1,000. Storing cluster representatives was the next best on average. Banking the patients about which the frozen model was least certain was harmful at small sizes, costing 0.125 AUROC for mortality in MIMIC-III at 300 patients.

**Figure 2.**
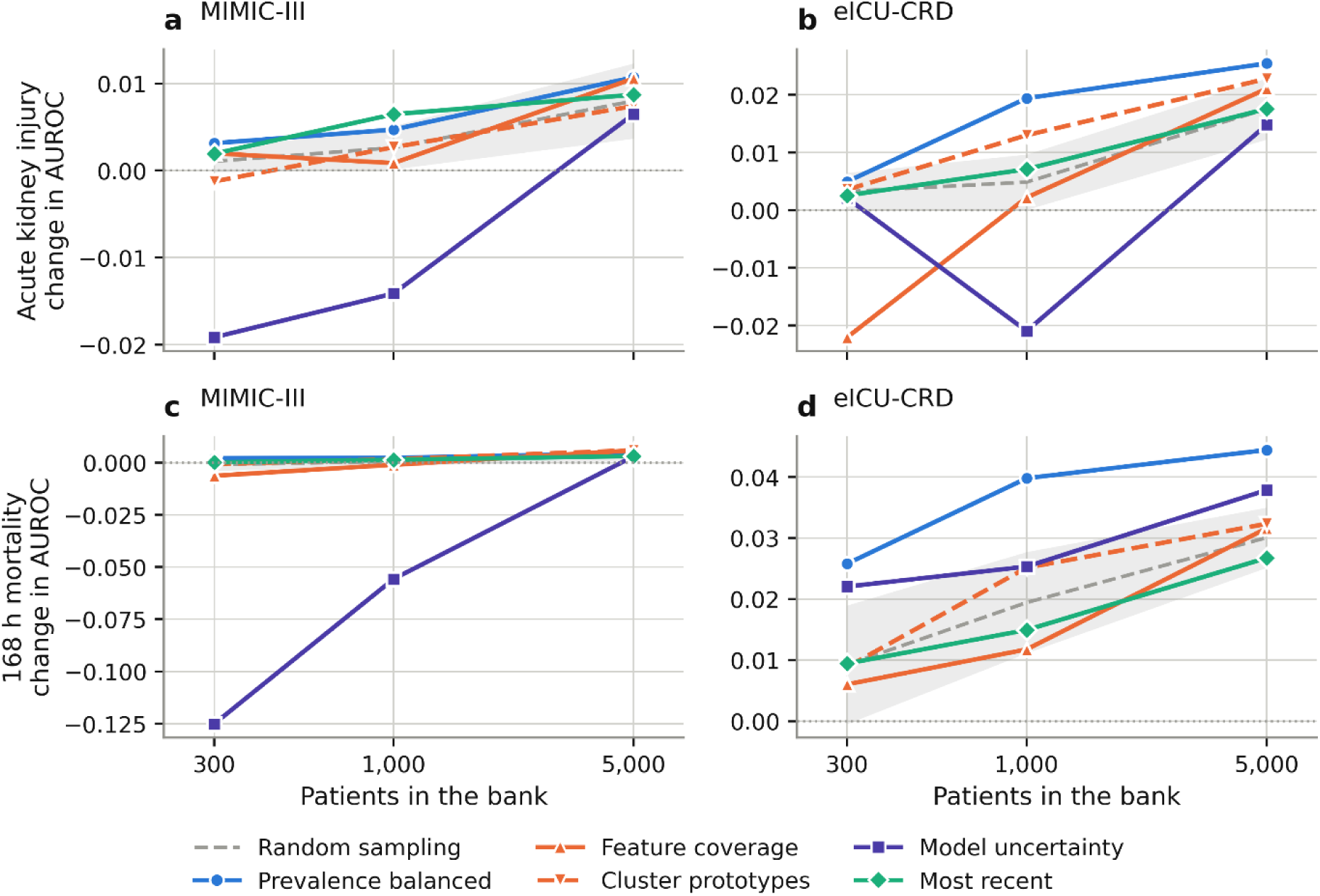
Bank composition at fixed size. The vertical axis is the change in AUROC against the frozen logistic model when the bank of the stated size is filled by each rule instead of at random. The shaded band is the mean and standard deviation of random sampling at the same sizes, so a curve above the band means the rule beats random sampling with the same number of patients.

At 1,000 patients, filling the bank from the most recent end of the pool gained 0.016 for acute kidney injury in eICU-CRD against 0.006 from the oldest end, and the ordering held in MIMIC-III (Supplementary Table S8).

Tripling the weight of the renal variable family improved retrieval for acute kidney injury in MIMIC-III, from 0.011 to 0.014 at the full bank, and reduced the gain for mortality. Every reduction of the variable set lost ground against uniform weighting, and no restriction of neighbours to the same sex, age band or creatinine tertile helped consistently (Supplementary Table S9).

Applied to the frozen CatBoost model, retrieval added 0.011 AUROC for acute kidney injury and 0.007 for mortality in eICU-CRD, and between -0.0003 and +0.001 in the two lower shift cohorts. Applied to a locally retrained logistic model it added 0.004 and 0.007 in eICU-CRD at the full bank, and 0.023 and 0.035 at 500 patients. Across five base model families and three retrieval measures in the development cohort, the rank correlation between base model AUROC and retrieval gain was -0.69 for acute kidney injury and -0.84 for mortality, and was negative in every split (Supplementary Table S10). The same ordering appeared when MIMIC-III was used as the development source (Supplementary Table S11). Retrieval was the only strategy that could be placed on top of another without a loss.

Making the mixing weight depend on prediction uncertainty and neighbour agreement did not improve discrimination by more than 0.001 AUROC in 17 of 18 settings, although it lowered the proportion of patients whose prediction was made worse (Supplementary Table S12). Learning the distance metric by neighbourhood components analysis was worse than plain cosine distance (Supplementary Table S13). Handling of missing data changed which strategy won: dropping the nine sparse variables let retrieval on the logistic model exceed frozen CatBoost for both outcomes in eICU-CRD (Supplementary Table S14).

Using the development split as a starting bank produced a gain before any local patient had been collected. In eICU-CRD for acute kidney injury this was 0.011 AUROC with no local data at all, close to what 5,000 local patients achieved without it. The advantage over a locally built bank of the same size shrank as local patients accumulated and was within 0.0011 AUROC at the full bank in every setting (Supplementary Tables S3 and S15). Sending such a bank moves development records to the receiving site, so we report it as a bound rather than as an option.

### The comparison at individual hospitals

Retrieval improved mortality prediction at 88 percent of 382 simulated hospitals of 300 to 3,000 patients, with a median change of 0.02, and acute kidney injury at 52 percent of 394, with a median change of 0.00 (Supplementary Figure S2). Varying the degree of case mix heterogeneity between those hospitals moved neither figure. Using the eICU-CRD hospital identifiers, the comparison was repeated site by site in 67 hospitals for acute kidney injury and 59 for mortality (Figure 3).

**Figure 3.**
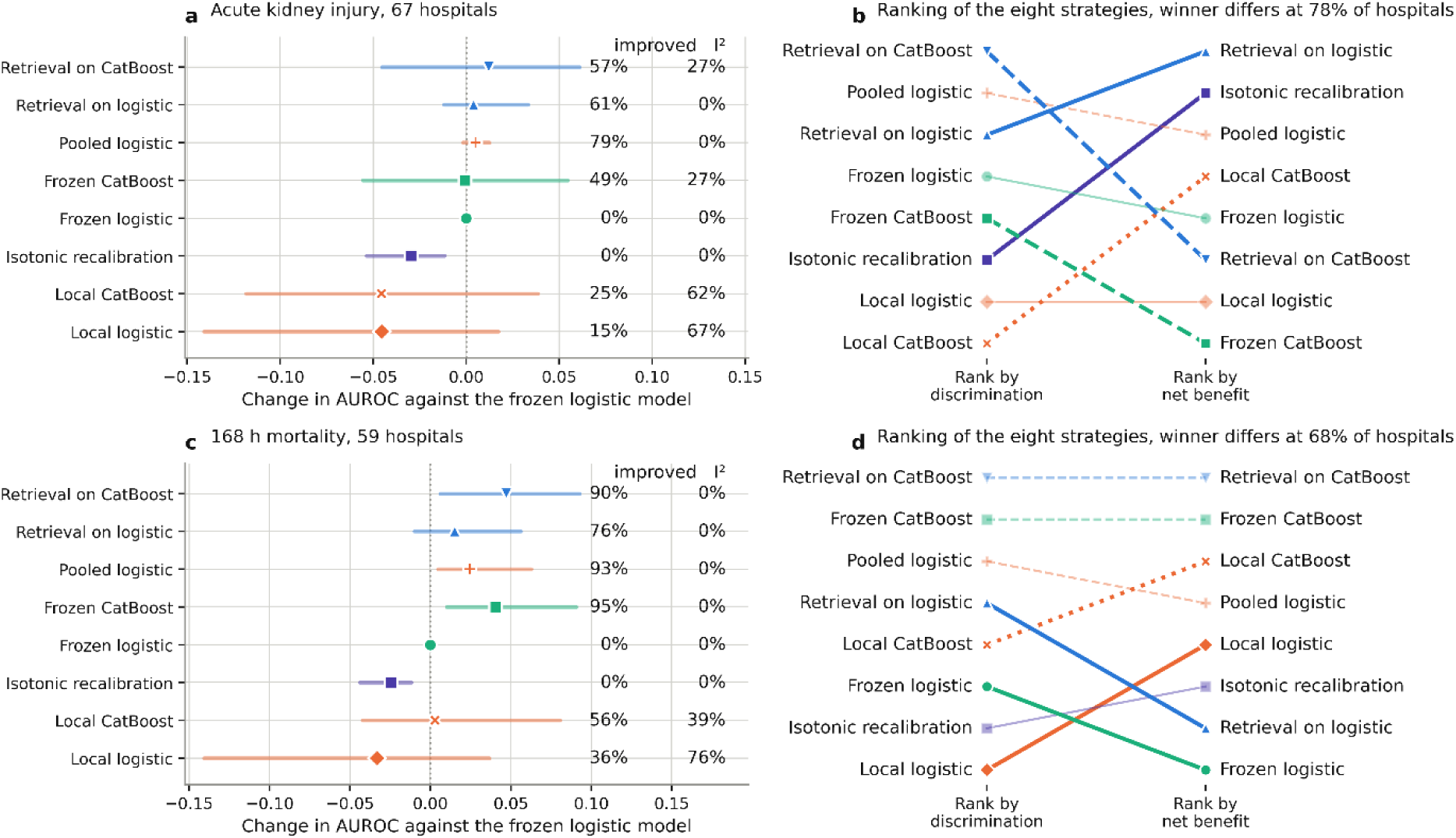
The same comparison at individual eICU-CRD hospitals, 67 for acute kidney injury and 59 for 168 hour mortality. **a**, Change in AUROC against the frozen logistic model at each hospital. The marker is the median across hospitals and the bar spans the 10th to 90th percentile of the hospital-level distribution. It is not a confidence interval, and it is wider than the interval on the pooled estimate quoted in the text and in Supplementary Table S16, because it carries the sampling error of each hospital rather than the precision of their average. The two right-hand columns give the percentage of hospitals that improved and the heterogeneity statistic I^2^. Strategies that leave the model frozen have I^2^ of 0 to 27 percent, while local retraining ranges from 39 to 76 percent. **b**, Rank of each strategy by discrimination against its rank by net benefit at a threshold probability of 20 percent, at every hospital. Lines drawn in full colour move at least two places between the two criteria. The title of each panel gives the percentage of hospitals at which the two criteria name a different winner.

Retrieval on the frozen logistic model gained a pooled 0.0092 AUROC (95 percent interval 0.0040 to 0.0144) for acute kidney injury and 0.0201 (0.0141 to 0.0262) for mortality, and in both cases the heterogeneity statistic I^2^ was 0 percent. Pooling with the development data behaved the same way, at 0.0068 (0.0054 to 0.0082) and 0.0302 (0.0247 to 0.0356), also with I^2^ of 0 percent. A locally fitted logistic model lost 0.0499 (-0.0677 to -0.0320) for acute kidney injury with I^2^ of 67 percent, and 0.0272 for mortality with I^2^ of 76 percent. A locally fitted CatBoost model lost 0.0368 with I^2^ of 62 percent for acute kidney injury and gained 0.0146 with I^2^ of 39 percent for mortality. Among the strategies that leave the model frozen the highest value was 27 percent, so heterogeneity above that was confined to the strategies that refit the model.

Retrieval on the frozen logistic model helped 61 percent of hospitals for acute kidney injury and 76 percent for mortality, and retrieval on CatBoost helped 57 and 90 percent. Pooled retraining helped more hospitals than either, 79 and 93 percent, and it needs the development records to be present at the deploying site. Local logistic retraining helped 15 and 36 percent. Isotonic recalibration lowered AUROC at every hospital in both outcomes. Across hospitals the frozen gradient boosted model gained a pooled 0.0077 AUROC for acute kidney injury and 0.0486 for mortality, and retrieval on it reached 0.0140 and 0.0484. These hospitals hold a median of 375 to 1,139 patients for acute kidney injury and 405 to 1,249 for mortality, which is at the low end of the dose response in Table 2.

Ranking the eight strategies by AUROC and by net benefit at a 20 percent threshold gave a different winner at 78 percent of hospitals for acute kidney injury and 68 percent for mortality (Fig. 3b). Isotonic recalibration was best on net benefit at 13 of 67 hospitals for acute kidney injury, more often than any other strategy, and it never improved AUROC at any hospital. The frozen CatBoost model led on AUROC at 27 of 59 hospitals for mortality and on net benefit at 10. The pooled estimate for every strategy, with the winner counts, is in Supplementary Table S16.

### Operating cost and exposure

Retrieval added 0.70 ms per patient at a 5,000 patient bank and 1.97 ms at 50,000, using exact rather than approximate nearest neighbour search, and needed 8.6 MB of memory at the larger size (Table 4). A batch of 100 patients took 4.9 ms.

**Table 4.** Cost of running retrieval and exposure of the bank.

**Panel A. Latency and memory**
| Bank size | Single patient (ms) | Batch of 100 (ms) | Memory (MB, float32) |
| --- | --- | --- | --- |
| 1,000 | 3.0 | 3.7 | 0.17 |
| 5,000 | 0.70 | 2.0 | 0.86 |
| 20,000 | 1.0 | 3.1 | 3.4 |
| 50,000 | 2.0 | 4.9 | 8.6 |

**Panel B. Re-identification risk and defences**
| Bank form | Attack AUC | Retrieval gain | Gain retained |
| --- | --- | --- | --- |
| Raw patient records | 1.00 | +0.0294 | 100% |
| Features coarsened to deciles<sup>a</sup> | 0.99 | +0.0290 | 99% |
| k-anonymity, k = 10 | 0.83 | +0.0272 | 93% |
| Cluster prototypes, 1,000 | 0.78 | +0.0307 | 104% |
| Noise on stored records, epsilon = 4 | 0.59 | +0.0260 | 88% |
| Prototypes with noise, epsilon = 4 | 0.55 | +0.0264 | 90% |
| Noise on the output probability, epsilon = 4 | 1.00 | +0.0291 | 99% |
| Refusal of the closest 5% of queries | 1.00 | +0.0293 | 100% |

Panel B reports a membership inference attack that scores each patient by distance to the nearest bank record, with the retrieval gain the same bank delivers. Values are for 168 hour mortality in eICU-CRD at a bank of 5,000 patients, except the prototype rows, which hold 1,000 prototypes. Only the defences that alter the stored records lower the attack. Those applied after retrieval leave it at 1.00. <sup>a</sup>each value replaced by the midpoint of its decile in the original scale

The bank is a store of real records, so we measured what an attacker with access to it could learn. A membership inference attack identified bank members perfectly, with an area under the curve of 1.00. Decile coarsening, k-anonymity at k = 10 and cluster prototypes lowered the attack only to 0.99, 0.81 to 0.83 and 0.77 to 0.78. Calibrated noise at epsilon = 4 lowered it to 0.59 and prototypes with noise to 0.55, retaining 88 and 90 percent of the raw-bank gain for mortality in eICU-CRD (Supplementary Figure S3). Perturbing the output probability, or refusing close queries, left the attack at 1.00. All eight bank forms are in Supplementary Table S17 for eICU-CRD, with three of them repeated in MIMIC-III.

### Retrieval decomposes into per-site sufficient statistics

The retrieval estimate is a weighted mean of bank outcomes, so its numerator and denominator are sums. Each site returns sum(w y) and sum(w) over its own neighbours, and the querying site adds them, which reproduces the number a single pooled bank would have produced.

We confirmed this on the eICU-CRD hospitals with a network of 20 partners. Reconstructing the estimate from the top neighbours reported by each site reproduced the pooled raw bank to within 3e-07 AUROC, which is floating point error rather than approximation (Supplementary Figure S4). Sending only the two scalars is an approximation rather than an identity, and it agreed with the exact version to a median absolute difference of 0.027, with a correlation of 0.795. Each participating site transmits two floating point numbers per query, which is 16 bytes, in a single round, and holds no state between queries.

Against a hospital’s own bank, joining the network was neutral for acute kidney injury (median 0.000) and slightly positive for mortality (median 0.0034, with 56 percent of hospitals improving). Most of the available gain arrived with the first one or two partners and then flattened for both outcomes. Adding Laplace noise to the two transmitted scalars, with a per-query privacy budget of epsilon of 1 or 4, changed the result by less than 0.001 AUROC. All eight schemes are in Supplementary Table S18. Federated learning was fitted across the same hospitals as a reference point rather than as a comparator. Federated model averaging and federated model stacking gained a median 0.0071 and 0.0111 AUROC over the frozen logistic model for acute kidney injury, and 0.0400 and 0.0357 for mortality. Adding retrieval on top of the federated stack changed the median by 0.0000 and 0.0009, and the selected mixing weight fell from 0.40 to 0.25 for acute kidney injury and from 0.45 to 0.20 for mortality. Table 5 lists these schemes by what has to leave the institution and what the participants have to agree on.

**Table 5.** What leaves the institution and what the participants must agree on.

| Level | What leaves the institution | What has to be agreed | Timing | Evidence here |
| --- | --- | --- | --- | --- |
| 0 | Nothing | Nothing | After deployment | Own bank (Fig. 1) |
| 1 | One probability per query | Outcome definition and query fields | After deployment | Retrieval network (Fig. 4) |
| 2 | Two scalars per query (16 bytes) | The above, plus the weight scale | After deployment | Aggregate scheme (Supplementary Fig. S4) |
| 3 | Distance and outcome of the k nearest | The above, plus distance function and k | After deployment | Exact scheme (Supplementary Fig. S4) |
| 4 | Prototypes, not real patients | The above, plus the feature space | After deployment | Prototype network (Supplementary Fig. S4, Supplementary Table S18) |
| 5 | Model parameters or gradients | Architecture, feature space, training protocol | During training | Federated learning [32, 33] |
| 6 | Raw patient records | A data use agreement | Either | Pooled retraining (Fig. 1) |

Levels 0 to 4 can be added after a model is deployed. Level 5 requires the site to have taken part while the model was being trained. The level that reproduces a single pooled bank exactly is 3, and the level that requires the least agreement is 1, where the network never learns how a site defines a similar patient. The exact scheme at level 3 reproduced a single pooled bank to within $3e-07$ AUROC. Levels 1 and 2 send one round per query and hold no state between queries.

### A network that shares only probabilities

We tested the level that needs the least agreement, in which each of a median of 20 partner hospitals answers a query with one probability and the querying hospital combines the answers (Figure 4). Against the frozen logistic model, the network gained a median 0.0155 AUROC for acute kidney injury with 80 percent of hospitals improving, and 0.0234 for mortality with 85 percent improving. A hospital using only its own bank gained 0.0080 and 0.0202. Expected calibration error fell from 0.044 to 0.035 for acute kidney injury and from 0.053 to 0.037 for mortality. The calibration slope moved from 0.48 to 0.89 for mortality. For acute kidney injury it moved from 0.86 to 1.14, which replaces under-confidence with over-confidence of the same size rather than removing it. Nine ways of combining the partner probabilities were not distinguishable from one another (Supplementary Table S19). When one partner was corrupted, the largest loss for acute kidney injury was the locally fitted stack at 0.025 AUROC, and the median, the trimmed mean and the mean in logit space lost no more than 0.001. Losses were smaller for mortality, where no rule lost more than 0.010.

**Figure 4.**
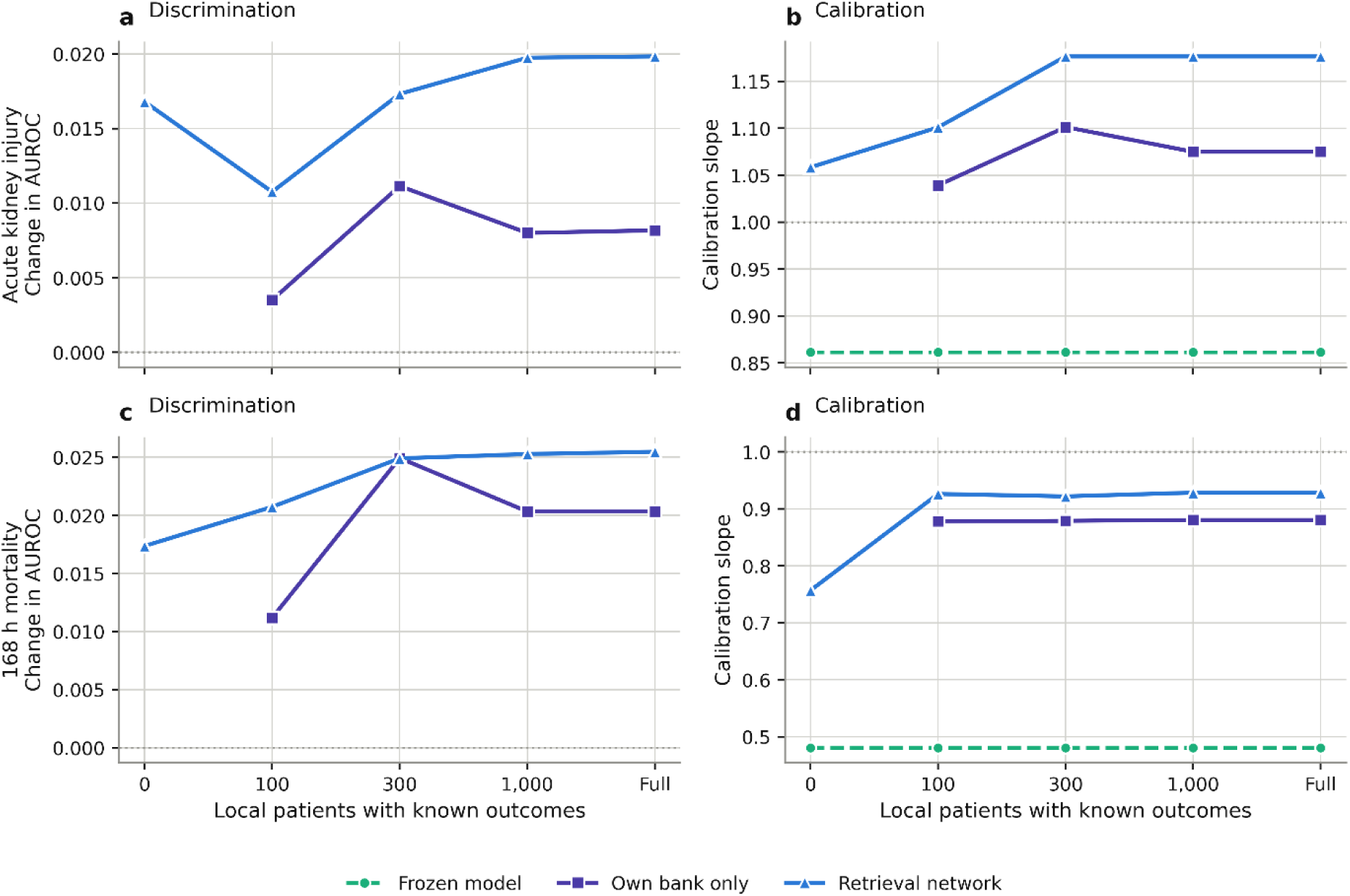
A retrieval network in which each partner hospital returns a single probability. The horizontal axis is the number of labelled local patients the querying hospital holds, starting from none. The left column is the change in AUROC against the frozen logistic model and the right column is the calibration slope, where 1.00 is ideal. The network line is the mean of the partner probabilities, the comparison line uses the hospital’s own bank alone, and the dashed line is the frozen model. At zero local labels the hospital can use no other strategy in this study, and the network still moves both measures.

A hospital with no labelled local outcomes cannot recalibrate, cannot retrain and cannot build a bank, so none of the strategies in Table 3 is open to it. It can still send queries. Doing so gained a median 0.0168 AUROC for acute kidney injury, with 84 percent of hospitals improving, and 0.0174 for mortality, with 88 percent improving. The calibration slope moved from 0.86 to 1.06 and from 0.48 to 0.76. For acute kidney injury this is about twice what the same hospitals obtained from their own full bank without the network, and for mortality it is most of it (Fig. 4). The curve is close to flat from 300 patients upward. The nine combination rules at the full bank, and the mean rule at every local bank size, are in Supplementary Table S19. A hospital that can reach no network at all can average the frozen model output over its own neighbours, which needs no outcomes and recovered much of the labelled gain where the shift was large (Supplementary Table S20).

### How much is still open

Five choices fixed in advance were compared with the best choice in hindsight, which cannot be used at deployment (Supplementary Figure S5). Picking the best of the eight strategies per hospital in hindsight would have added 0.0255 AUROC over the pre-specified retrieval. Choosing which partners to query was next at 0.0209, and the hindsight-optimal network was a median of one or two of the 20 available partners. The combination rule was worth 0.0063, the bank composition rule 0.0034, and the mixing weight 0.0009 (Supplementary Table S21).

Nine ways of defining the reference class were compared as a sensitivity analysis (Supplementary Figure S6). None dominated. Random forest proximity was the best estimator for acute kidney injury in MIMIC-III, adding 0.016 to 0.017 AUROC over the pre-specified cosine rule, and among the worst for mortality in eICU-CRD, losing 0.008. Predicted risk band alone was selected out entirely, with a mixing weight of zero in every setting. Cost differed by four orders of magnitude, from 0.001 to 4.86 seconds per 2,000 queries (Supplementary Table S22). Assigning different definitions to different partners lowered the correlation between their answers, from 0.32 to 0.60 down to 0.23 to 0.40, but the heterogeneous networks stayed within the spread of the homogeneous ones (Supplementary Figure S7, Supplementary Table S23). All main analyses use the pre-specified cosine rule.

## Discussion

Both frozen models were miscalibrated on the new population, and net benefit at a fixed threshold depends on the probability scale rather than on the ranking of patients. A site that compares adaptation strategies on discrimination alone can therefore install the option that helps its patients least. Pooling local patients with the development data avoids the loss that local retraining alone incurs, at the cost of moving the development records to the deploying site. The hospital-level analysis adds a property the averages hide: heterogeneity above 27 percent was confined to the strategies that refit the model. A site is therefore choosing both an expected gain and how much that gain depends on which site it is.

Retrieval lost almost no discrimination in any cohort, and at individual hospitals the tenth percentile for retrieval on the frozen logistic model was -0.0119 for acute kidney injury and -0.0098 for mortality (Fig. 3a). We attached it to four kinds of base model and to five model families in the development cohort. The gain grew with the size of the population shift and fell as the base model became stronger. The layer can therefore be attached whatever method produced the model, and how good that model already is sets how much the layer adds. A site with a strong frozen model and thousands of labelled patients has other options and gains little. A site with no labelled outcomes has none of the eight strategies open to it. The layer’s most consistent effect was on calibration.

In eICU-CRD a balanced bank outperformed a larger random one and a recent bank outperformed an older one of the same size, so a site can improve retrieval by curating the bank rather than by enlarging it, without any change to the model. Because the retrieval estimate is a ratio of two sums, it decomposes into quantities that each site can compute over its own records, and the reconstruction matched a pooled raw bank to floating point precision. In practice this is 16 bytes per site per query, one round and no state, or a single probability if the participants do not want to agree on a distance function. Table 5 sets out the levels of disclosure this creates. The level that reproduces a single pooled bank exactly is the one that sends the nearest neighbours; the level that sends only a probability gives up some accuracy and in exchange never reveals how a site defines a similar patient. Federated learning sits one level higher, since it moves parameters and is applied while the model is being built.

A hospital outside a federated consortium cannot join it once the model has shipped, but it can still attach a retrieval layer to the model that consortium produced, so the two combine rather than compete. A hospital that holds no labelled outcomes has nothing to contribute to a collaboration arranged at training time, and querying a network after deployment asks for no contribution. Such a site sends queries, receives probabilities, and gained discrimination and calibration at most of the sites we tested, with the calibration slope moving from 0.86 to 1.06 and from 0.48 to 0.76. The sites that most need a working model are often the ones with too little data to earn a place in its development, and a network that admits consumers lets them benefit on the day the tool arrives. A consumer does not improve the network, so how contribution should be rewarded is a question our data cannot answer.

The same disagreement has been reported between architectures. In a multi-centre study of continuous acute kidney injury monitoring, the architecture with the highest AUROC had the worst alert burden, while the architecture ranked third had the best.^31^ That comparison is made when a model is built; ours is between the strategies a hospital can apply to a model it already has. That study also used post-hoc isotonic recalibration, fitted on each site’s own data, to adapt models without retraining. It is one of the eight strategies we compare, and at the full bank in eICU-CRD it gave the lowest expected calibration error of any strategy while lowering discrimination at every hospital.

Our results agree with reports that multi-centre training is the most reliable way to obtain transportable intensive care models^7^ and with the three-regime account of external data utility from anchor regression,^21^ to which our bank size axis adds where those regimes begin. A recent perioperative network study across 23 hospitals found half of the multicentre gain already present at four hospitals,^33^ and the same diminishing return appears in a network assembled after deployment rather than during training: our network size curve flattens after one or two partners. What differs is where the collaboration sits, which is why a site with no labelled outcomes can join ours. Retrieval built into a network architecture has instead been reported to go stale under temporal shift, because the distance representation is learned once;^34,35^ a network of that type performed worse than retrieval on a frozen logistic model in eICU-CRD (Supplementary Table S24).

A bank holds records the hospital already has, under the same access controls, so an attack that succeeds against it restates that intensive care records are identifiable. Exposure arises when the bank leaves the institution, which the lower levels in Table 5 avoid. Where a bank has to be shared, prototypes and calibrated noise brought the attack close to chance while keeping most of the gain. Noise on the transmitted probability did not lower the attack at all, because it does not touch the records. Common data models such as OMOP already provide the harmonization we did by hand as infrastructure and already standardize the query fields a network has to agree on,^36^ and secure aggregation would let the two scalars be summed without any site seeing the individual terms.^37^

Retrieved neighbours are patients of the deploying hospital, so a prediction arrives with records the attending clinician can open, which feature attribution does not offer^38,39^ and which is close to how clinicians reason from remembered cases.^40,41^ It also raises a question the model does not ask: who counts as a similar patient. This is the reference class problem, and the answer depends on what is being decided.^42^ Of the nine definitions we compared, including proximity in a random forest,^43^ none was best across cohorts and outcomes. Similarity is computed outside the model, so a site can weight, restrict or replace the input space, and could use fields the model never sees. On the lowest disclosure level all of this stays local.

Three features support these conclusions: the comparison was run at individual hospitals rather than at database level, we ran a membership inference attack against our own bank and report which defences fail, and we measured the distance between each pre-specified choice and the best choice in hindsight. No model in this study was installed at a site and used on a patient. The comparison is a simulation run on recorded outcomes, so it is not evidence about deployed care. The feature set is 42 static variables from the first 24 hours, without medications, time series or notes, and all three databases hold North American intensive care populations. Hospital-level results come from eICU-CRD alone. Banks were formed by sampling from a pool rather than by sequential arrival. We make no claim at the level of the individual patient, since any probability adjustment moves some patients in the wrong direction and our design cannot separate that from benefit. Net benefit was computed at fixed thresholds, and a site using another threshold may reach a different ordering. We do not claim that a bank replacement is exempt from regulatory review, only that the change is confined to a data artefact rather than to model parameters, which is the kind of change a predetermined change control plan can specify in advance.

A hospital installing a model it did not build should compare its options on calibration and net benefit as well as discrimination, and against the number of labelled local patients it holds. Below a few thousand local patients, retrieval is the safe option. Local retraining takes over above that, at sites where the population has moved a long way. Because its estimate is a sum of parts held at different sites, hospitals can improve each other’s predictions by exchanging probabilities rather than patients, including hospitals that hold no labelled outcomes of their own. What remains to be settled is how large such a network has to be, how a site should define a similar patient, and how the answers from several sites should be combined.

## Methods

### Data and cohort

Three de-identified intensive care databases hosted on PhysioNet were used:^44^ MIMIC-IV v2.2,^45^ MIMIC-III v1.4^46^ and the eICU Collaborative Research Database v2.0.^47^ MIMIC-IV and MIMIC-III come from a single centre over non-overlapping periods, 2008 to 2019 and 2001 to 2008. eICU-CRD covers more than 200 United States hospitals in 2014 and 2015.

Adults aged 18 years or older with a first intensive care stay of at least 24 hours were included. Only the first stay of each patient was used, so that every row is one independent admission and no patient contributes both a bank record and a test record.

The prediction anchor was 24 hours after admission. Two binary outcomes were defined within 168 hours of the anchor. The first is acute kidney injury identified here from the rise in serum creatinine defined by the Kidney Disease Improving Global Outcomes criteria, using the minimum creatinine of the first 24 hours as the baseline.^48,49^ The second is in-hospital death from any cause. Both are acted on at a fixed probability threshold and are ascertainable from the same fields in all three databases.

### Features and preprocessing

Forty-two features were taken from the first 24 hours: age and sex, baseline creatinine, seven vital signs with weight, and five laboratory measurements (Supplementary Table S1). The list was assembled from earlier work with clinical teams, from variables that are recorded routinely in intensive care, are established predictors of acute kidney injury and death, and are present in all three databases. It was fixed before any model was fitted and no data-driven selection was applied at any stage. Each variable was reduced to its minimum, maximum and last value across the 24 hour window, giving a single row per patient.

Harmonization mapped the item identifiers of MIMIC-III and eICU-CRD onto the MIMIC-IV template by clinical concept and unit, converting units where they differed and applying the same physiological range filter to all three databases. The mapping uses only variable identity and units, never the outcome. Numeric features were imputed by multiple imputation by chained equations^50^ and standardized; categorical features were mode imputed and one-hot encoded. Every imputation and scaling parameter was estimated on the MIMIC-IV training split alone and applied unchanged everywhere else. The main analysis imputes every variable regardless of how much of it is missing, because laboratory values in intensive care are missing not at random and the fact that a test was not ordered carries information. Five alternative handlings were run alongside it and are reported in Supplementary Table S14.

### Strategies compared

MIMIC-IV served as the development cohort, split into 56 percent training, 14 percent validation and 30 percent internal test. Two frozen base models were trained on the training split: a logistic regression and a CatBoost model.^51^ Eight strategies were then applied at each deployment site.

*Frozen logistic* and *frozen CatBoost* apply the development model unchanged. *Isotonic recalibration* fits a monotone transform of the frozen logistic output on the local bank. *Retrieval on logistic* and *retrieval on CatBoost* apply PRAM to the corresponding frozen model. *Local logistic* fits a new model on the bank alone. *Pooled logistic* fits a new model on the bank together with the MIMIC-IV training split, which requires the development records at the deploying site and is therefore an upper bound rather than a routine option. *Local CatBoost* fits a new gradient boosted model on the bank alone. Two further variants were evaluated alongside the eight: a label-free version that averages the frozen model output over the same neighbours, and a cold start in which the development split serves as the bank.

PRAM forms the augmented prediction as p_mix_ = (1 - a) p_base_(x) + a p_retr_(x). Here p_retr_ is the weighted mean of the observed outcomes y of the 50 nearest patients in the bank, each weighted by w = 1 / (d + 1e-6), where d is that patient’s distance to the query. The scalar a, the mixing weight, is chosen from eleven values from 0 to 0.50 in steps of 0.05. Only the bank changes when the model moves. Model settings, the neighbour count and the grid searched for a are listed in Supplementary Table S25. Three distance metrics were compared on the development cohort; because they were not distinguishable (Supplementary Table S26), cosine distance was pre-specified for all deployment analyses.

### Deployment simulation

The simulation was run once for every combination of target cohort, outcome and local bank size N. Each target cohort was divided into five stratified folds. In each fold in turn, that fold is the test set and the other four form the pool of local patients the site is imagined to hold. A bank of size N is drawn at random from the pool and every strategy is fitted using only that bank, so one pass over the five folds gives every patient a prediction from a bank that did not hold them. Bank sizes were 500, 1,000, 2,000, 5,000 and the whole pool, and at each intermediate size the pass was repeated ten times with a new draw. Each pass is scored once, and we report the median across the ten. The significance testing below uses thirty passes at each intermediate size.

The mixing weight was selected by three-fold cross-validation inside the bank, so no test patient contributed to it. Each draw was scored on its own rather than pooled with other draws; pooling first inflates the apparent gain by about 0.004 AUROC at a bank of 5,000 and by up to 0.008 in eICU-CRD. Reversing the roles of MIMIC-III and MIMIC-IV gives a second, smaller and older development source and brings the total to 12 deployment scenarios.

Bank composition was varied at fixed size using six rules: random sampling, greedy coverage of the feature space, model uncertainty, prevalence balancing, recency, and cluster representatives. Bank recency was also tested by filling the bank from the oldest or the newest end of the pool. Retrieval similarity was varied by weighting variable families relevant to each outcome and by restricting neighbours to the same sex, age band or baseline creatinine tertile.

### Hospital-level analysis

The eICU-CRD hospital identifier was used to repeat the whole comparison at real sites. Hospitals with at least 300 stays and at least 15 events and 15 non-events were retained, which gave 67 hospitals for acute kidney injury and 59 for mortality. Within each hospital, three-fold cross-validation provided the local bank and the test patients, so that no patient contributed to both. Strategy effects were pooled across hospitals as a mean weighted by the number of stays at each hospital, with a 95 percent interval from the standard error of that mean across hospitals, and heterogeneity was summarized by an I^2^ statistic computed from the same weighted Q.^52^ Simulated hospitals of 300 to 3,000 patients drawn from eICU-CRD under two settings of case mix heterogeneity are reported as a sensitivity analysis.

### Federated retrieval and the retrieval network

Eight federated schemes were implemented over networks of up to 20 partner hospitals (Table 5). In the exact scheme each partner returns the distance and outcome of its k nearest bank patients and the querying site merges the lists, which reproduces a single pooled bank by construction. In the aggregate scheme each partner returns only sum(w y) and sum(w), with a differentially private variant adding Laplace noise to those two scalars at epsilon = 1 or 4.^53^ In the probability-only network each partner returns a single mixed probability, and nine combination rules were compared against the local bank alone (Supplementary Table S19). Local bank sizes from 0, meaning a hospital with no labelled outcomes, to the whole local pool were crossed with every rule, and robustness was tested by permuting the outcome labels at one partner. Federated averaging and federated stacking were fitted across the same hospitals as a coordinate check, with retrieval applied on top of the federated stack.

### Privacy analysis

Re-identification risk was assessed by a membership inference attack^54^ that scores each patient by distance to the nearest bank record. Seven defences were applied to the bank and each is reported as a pair of the attack area under the curve and the retained AUROC gain (Supplementary Table S17).

### Oracle bounds and reference class estimators

Five pre-specified choices were compared with the best choice in hindsight, chosen on the test data and therefore not available at deployment: the mixing weight, the strategy, the bank composition rule, the combination rule and the partner subset. Nine estimators of the reference class, including random forest proximity,^43^ were compared as a sensitivity analysis, both within and across partners; they are listed in Supplementary Table S22 and none was allowed to change the main analysis, which uses the pre-specified cosine rule throughout.

### Outcomes of the comparison and statistical analysis

Discrimination was measured by AUROC and AUPRC. Calibration was measured by expected calibration error and by the slope and intercept of the recalibration line. Clinical utility was measured by net benefit from decision curve analysis at thresholds from 2 to 30 percent. Retrieval latency and memory were profiled on a single processor with exact nearest neighbour search.

The relationship between bank size and performance was modelled with a linear mixed model of AUROC on log10 bank size with a fold-level random intercept. Paired DeLong tests^55^ compared strategies with the frozen model on pooled out-of-fold predictions, computed separately for each of thirty bank draws at every intermediate size; we report the median p value across draws and the proportion of draws reaching significance. Benjamini-Hochberg correction^56^ was applied across the six cohort and outcome combinations within each family. Analyses used Python 3.11 with scikit-learn 1.3, CatBoost 1.2 and SciPy 1.10. Reporting follows TRIPOD+AI;^57^ the completed checklist is Supplementary Note 1.

## Data Availability

The datasets analyzed in this study are publicly available through PhysioNet (https://physionet.org/) under credentialed data use agreements: MIMIC-IV v2.2, MIMIC-III v1.4, and eICU-CRD v2.0.

## Statements

## Data availability

MIMIC-IV v2.2, MIMIC-III v1.4 and eICU-CRD v2.0 are available from PhysioNet under credentialed data use agreements.

## Code availability

The analysis code is available from the corresponding author on reasonable request.

## Ethics

All databases contain de-identified data and are exempt from institutional review board approval.

## Acknowledgements

This study used de-identified data from MIMIC-IV, MIMIC-III and eICU-CRD, accessed through PhysioNet. The authors thank the PhysioNet team and the contributing institutions for making these data publicly available. No patients or members of the public were involved in the design, conduct, reporting or dissemination of this study.

## Author contributions

Inyong Jeong: conceptualization, methodology, software, formal analysis, investigation, data curation, writing of the original draft, visualization. Taeyeong Lee: conceptualization, data curation. Byeongsu Kim: conceptualization, data curation. Jin-Hyun Park: conceptualization, writing, review and editing. Yeongmin Kim: conceptualization, writing, review and editing. Hwamin Lee: conceptualization, supervision, writing, review and editing, project administration.

## Use of generative artificial intelligence

Generative artificial intelligence tools were used for language editing during the preparation of this manuscript. The authors reviewed and edited all such content and take full responsibility for the final text.

---

**Supplementary Table S1.** Cohort characteristics, features and missingness by database.

Panel A. Model inputs by variable family
| Family | Variable | Derived columns | MIMIC-IV | MIMIC-III | eICU-CRD |
| --- | --- | --- | --- | --- | --- |
| Demographics | Age | 1 | 0.0 | 0.0 | 0.0 |
| Demographics | Sex | 1 | 0.0 | 0.0 | 0.0 |
| Baseline | Serum creatinine, first 24 h minimum | 1 | 0.0 | 0.0 | 0.0 |
| Vital signs | Heart rate | 3 | 0.1 | 0.7 | 1.4 |
| Vital signs | Respiratory rate | 3 | 0.3 | 0.8 | 8.2 |
| Vital signs | Oxygen saturation | 3 | 0.1 | 0.7 | 2.2 |
| Vital signs | Temperature | 3 | 2.4 | 0.7 | 88.9 |
| Vital signs | Systolic blood pressure | 3 | 0.4 | 0.7 | 3.3 |
| Vital signs | Diastolic blood pressure | 3 | 0.4 | 0.7 | 3.3 |
| Vital signs | Glasgow Coma Scale total | 3 | 0.2 | 0.8 | 4.6 |
| Vital signs | Weight | 3 | 0.6 | 41.1 | 0.3 to 74.1 |
| Laboratory | Blood urea nitrogen | 3 | 0.0 | 0.1 | 0.7 |
| Laboratory | Calcium | 3 | 8.6 | 17.5 | 3.6 |
| Laboratory | International normalized ratio | 3 | 12.5 | 7.0 | 60.9 |
| Laboratory | Serum creatinine | 3 | 0.0 | 0.0 | 0.0 |
| Laboratory | Blood urea nitrogen to creatinine ratio | 3 | 0.0 | 0.1 | 0.7 |
| **Total** |  | **42** | **>30% in 0 of 42** | **>30% in 3 of 42** | **>30% in 7 of 42** |

Panel B. Descriptive statistics for every column in every database
| Variable | Database | Missing (%) | Median | Q1 | Q3 | Mean | SD |
| --- | --- | --- | --- | --- | --- | --- | --- |
| age | MIMIC-III | 0.0 | 64.31 | 51.36 | 76.3 | 62.33 | 16.8 |
| age | MIMIC-IV | 0.0 | 66.0 | 54.0 | 77.0 | 63.84 | 16.91 |
| age | eICU-CRD | 0.0 | 65.0 | 53.0 | 75.0 | 62.67 | 16.46 |
| baseline_scr | MIMIC-III | 0.0 | 0.8 | 0.6 | 1.1 | 0.94 | 0.51 |
| baseline_scr | MIMIC-IV | 0.0 | 0.8 | 0.7 | 1.1 | 0.97 | 0.52 |
| baseline_scr | eICU-CRD | 0.0 | 0.89 | 0.7 | 1.2 | 1.05 | 0.59 |
| bun_cr_ratio_last | MIMIC-III | 0.1 | 18.57 | 14.29 | 24.29 | 20.74 | 10.23 |
| bun_cr_ratio_last | MIMIC-IV | 0.0 | 18.57 | 14.38 | 24.29 | 20.54 | 9.67 |
| bun_cr_ratio_last | eICU-CRD | 0.7 | 18.18 | 13.75 | 24.24 | 20.33 | 10.43 |
| bun_cr_ratio_max | MIMIC-III | 0.1 | 20.0 | 15.45 | 25.83 | 22.05 | 10.65 |
| bun_cr_ratio_max | MIMIC-IV | 0.0 | 20.0 | 15.56 | 25.71 | 21.9 | 10.13 |
| bun_cr_ratio_max | eICU-CRD | 0.7 | 18.18 | 13.86 | 24.24 | 20.43 | 10.37 |
| bun_cr_ratio_min | MIMIC-III | 0.1 | 18.0 | 14.0 | 23.33 | 19.91 | 9.59 |
| bun_cr_ratio_min | MIMIC-IV | 0.0 | 17.78 | 13.75 | 23.04 | 19.58 | 9.13 |
| bun_cr_ratio_min | eICU-CRD | 0.7 | 18.0 | 13.64 | 24.12 | 20.24 | 10.45 |
| bun_last | MIMIC-III | 0.1 | 16.0 | 11.0 | 23.0 | 20.02 | 15.08 |
| bun_last | MIMIC-IV | 0.0 | 16.0 | 12.0 | 24.0 | 20.6 | 15.58 |
| bun_last | eICU-CRD | 0.7 | 17.0 | 11.0 | 26.0 | 21.4 | 16.08 |
| bun_max | MIMIC-III | 0.1 | 17.0 | 12.0 | 25.0 | 21.41 | 16.22 |
| bun_max | MIMIC-IV | 0.0 | 17.0 | 12.0 | 26.0 | 22.21 | 16.89 |
| bun_max | eICU-CRD | 0.7 | 17.0 | 12.0 | 27.0 | 22.55 | 17.37 |
| bun_min | MIMIC-III | 0.1 | 15.0 | 11.0 | 23.0 | 19.51 | 14.91 |
| bun_min | MIMIC-IV | 0.0 | 16.0 | 11.0 | 23.0 | 19.96 | 15.28 |
| bun_min | eICU-CRD | 0.7 | 16.0 | 11.0 | 25.0 | 21.03 | 15.96 |
| calcium_last | MIMIC-III | 17.5 | 8.3 | 7.8 | 8.8 | 8.31 | 0.74 |
| calcium_last | MIMIC-IV | 8.6 | 8.4 | 7.9 | 8.8 | 8.36 | 0.69 |
| calcium_last | eICU-CRD | 3.6 | 8.3 | 7.8 | 8.8 | 8.28 | 0.73 |
| calcium_max | MIMIC-III | 17.5 | 8.4 | 7.9 | 8.9 | 8.42 | 0.75 |
| calcium_max | MIMIC-IV | 8.6 | 8.5 | 8.0 | 8.9 | 8.5 | 0.71 |
| calcium_max | eICU-CRD | 3.6 | 8.4 | 7.9 | 8.8 | 8.38 | 0.74 |
| calcium_min | MIMIC-III | 17.5 | 8.2 | 7.7 | 8.7 | 8.17 | 0.82 |
| calcium_min | MIMIC-IV | 8.6 | 8.2 | 7.8 | 8.7 | 8.22 | 0.77 |
| calcium_min | eICU-CRD | 3.6 | 8.2 | 7.7 | 8.7 | 8.21 | 0.77 |
| dbp_last | MIMIC-III | 0.7 | 58.0 | 50.0 | 68.0 | 59.58 | 13.38 |
| dbp_last | MIMIC-IV | 0.4 | 62.0 | 53.0 | 72.0 | 63.57 | 14.66 |
| dbp_last | eICU-CRD | 3.3 | 65.0 | 56.0 | 75.0 | 66.32 | 14.86 |
| dbp_max | MIMIC-III | 0.7 | 80.0 | 71.0 | 90.0 | 81.34 | 15.46 |
| dbp_max | MIMIC-IV | 0.4 | 86.0 | 75.0 | 99.0 | 88.48 | 19.32 |
| dbp_max | eICU-CRD | 3.3 | 93.0 | 80.0 | 108.0 | 96.54 | 23.21 |
| dbp_min | MIMIC-III | 0.7 | 43.0 | 37.0 | 50.0 | 43.63 | 10.39 |
| dbp_min | MIMIC-IV | 0.4 | 47.0 | 40.0 | 54.0 | 47.38 | 10.73 |
| dbp_min | eICU-CRD | 3.3 | 47.0 | 39.0 | 55.0 | 47.3 | 12.27 |
| gcs_total_last | MIMIC-III | 0.8 | 15.0 | 11.0 | 15.0 | 12.96 | 3.32 |
| gcs_total_last | MIMIC-IV | 0.2 | 15.0 | 13.0 | 15.0 | 13.24 | 3.12 |
| gcs_total_last | eICU-CRD | 4.6 | 15.0 | 12.0 | 15.0 | 12.87 | 3.47 |
| gcs_total_max | MIMIC-III | 0.8 | 15.0 | 14.0 | 15.0 | 13.58 | 2.65 |
| gcs_total_max | MIMIC-IV | 0.2 | 15.0 | 14.0 | 15.0 | 13.76 | 2.55 |
| gcs_total_max | eICU-CRD | 4.6 | 15.0 | 12.0 | 15.0 | 12.87 | 3.47 |
| gcs_total_min | MIMIC-III | 0.8 | 10.0 | 3.0 | 15.0 | 9.65 | 4.94 |
| gcs_total_min | MIMIC-IV | 0.2 | 13.0 | 3.0 | 15.0 | 10.06 | 4.97 |
| gcs_total_min | eICU-CRD | 4.6 | 15.0 | 12.0 | 15.0 | 12.87 | 3.47 |
| hr_last | MIMIC-III | 0.7 | 84.0 | 73.0 | 96.0 | 85.15 | 16.75 |
| hr_last | MIMIC-IV | 0.1 | 82.0 | 71.0 | 94.0 | 83.64 | 17.34 |
| hr_last | eICU-CRD | 1.4 | 83.0 | 71.0 | 96.0 | 84.26 | 17.88 |
| hr_max | MIMIC-III | 0.7 | 102.0 | 90.0 | 116.0 | 103.97 | 20.02 |
| hr_max | MIMIC-IV | 0.1 | 100.0 | 89.0 | 115.0 | 102.99 | 20.1 |
| hr_max | eICU-CRD | 1.4 | 107.0 | 93.0 | 122.0 | 108.82 | 21.73 |
| hr_min | MIMIC-III | 0.7 | 70.0 | 61.0 | 80.0 | 71.52 | 14.35 |
| hr_min | MIMIC-IV | 0.1 | 68.0 | 60.0 | 79.0 | 69.66 | 14.63 |
| hr_min | eICU-CRD | 1.4 | 68.0 | 59.0 | 79.0 | 69.05 | 15.25 |
| inr_last | MIMIC-III | 7.0 | 1.2 | 1.1 | 1.4 | 1.35 | 0.56 |
| inr_last | MIMIC-IV | 12.5 | 1.2 | 1.1 | 1.4 | 1.34 | 0.61 |
| inr_last | eICU-CRD | 60.9 | 1.2 | 1.1 | 1.5 | 1.46 | 0.8 |
| inr_max | MIMIC-III | 7.0 | 1.3 | 1.1 | 1.5 | 1.46 | 0.77 |
| inr_max | MIMIC-IV | 12.5 | 1.3 | 1.1 | 1.5 | 1.44 | 0.77 |
| inr_max | eICU-CRD | 60.9 | 1.2 | 1.1 | 1.5 | 1.53 | 0.95 |
| inr_min | MIMIC-III | 7.0 | 1.2 | 1.1 | 1.4 | 1.32 | 0.5 |
| inr_min | MIMIC-IV | 12.5 | 1.2 | 1.1 | 1.3 | 1.31 | 0.55 |
| inr_min | eICU-CRD | 60.9 | 1.2 | 1.1 | 1.44 | 1.44 | 0.77 |
| rr_last | MIMIC-III | 0.8 | 19.0 | 15.0 | 22.0 | 19.18 | 5.41 |
| rr_last | MIMIC-IV | 0.3 | 19.0 | 16.0 | 22.0 | 19.38 | 5.33 |
| rr_last | eICU-CRD | 8.2 | 19.0 | 16.0 | 23.0 | 19.97 | 6.5 |
| rr_max | MIMIC-III | 0.8 | 26.0 | 23.0 | 30.0 | 27.04 | 6.59 |
| rr_max | MIMIC-IV | 0.3 | 27.0 | 23.0 | 31.0 | 27.65 | 6.13 |
| rr_max | eICU-CRD | 8.2 | 29.0 | 25.0 | 37.0 | 32.36 | 11.21 |
| rr_min | MIMIC-III | 0.8 | 12.0 | 10.0 | 14.0 | 12.11 | 3.57 |
| rr_min | MIMIC-IV | 0.3 | 12.0 | 10.0 | 14.0 | 12.2 | 3.39 |
| rr_min | eICU-CRD | 8.2 | 11.0 | 8.0 | 14.0 | 11.08 | 4.28 |
| sbp_last | MIMIC-III | 0.7 | 119.0 | 106.0 | 134.0 | 120.83 | 20.72 |
| sbp_last | MIMIC-IV | 0.4 | 118.0 | 105.0 | 132.0 | 119.68 | 20.29 |
| sbp_last | eICU-CRD | 3.3 | 120.0 | 106.0 | 136.0 | 122.23 | 22.01 |
| sbp_max | MIMIC-III | 0.7 | 147.0 | 134.0 | 163.0 | 150.08 | 23.11 |
| sbp_max | MIMIC-IV | 0.4 | 146.0 | 133.0 | 161.0 | 148.52 | 21.85 |
| sbp_max | eICU-CRD | 3.3 | 151.0 | 135.0 | 170.0 | 153.7 | 26.7 |
| sbp_min | MIMIC-III | 0.7 | 92.0 | 83.0 | 103.0 | 93.24 | 16.3 |
| sbp_min | MIMIC-IV | 0.4 | 92.0 | 83.0 | 102.0 | 93.07 | 15.89 |
| sbp_min | eICU-CRD | 3.3 | 90.0 | 77.0 | 104.0 | 90.72 | 20.0 |
| scr_feat_last | MIMIC-III | 0.0 | 0.8 | 0.7 | 1.1 | 0.97 | 0.51 |
| scr_feat_last | MIMIC-IV | 0.0 | 0.9 | 0.7 | 1.1 | 1.0 | 0.53 |
| scr_feat_last | eICU-CRD | 0.0 | 0.9 | 0.7 | 1.2 | 1.06 | 0.59 |
| scr_feat_max | MIMIC-III | 0.0 | 0.9 | 0.7 | 1.1 | 1.02 | 0.56 |
| scr_feat_max | MIMIC-IV | 0.0 | 0.9 | 0.7 | 1.2 | 1.07 | 0.64 |
| scr_feat_max | eICU-CRD | 0.0 | 0.92 | 0.71 | 1.26 | 1.12 | 0.69 |
| scr_feat_min | MIMIC-III | 0.0 | 0.8 | 0.6 | 1.1 | 0.94 | 0.51 |
| scr_feat_min | MIMIC-IV | 0.0 | 0.8 | 0.7 | 1.1 | 0.97 | 0.52 |
| scr_feat_min | eICU-CRD | 0.0 | 0.89 | 0.7 | 1.2 | 1.05 | 0.59 |
| sex_male | MIMIC-III | 0.0 | not applicable | not applicable | not applicable | 0.58 | 0.49 |
| sex_male | MIMIC-IV | 0.0 | not applicable | not applicable | not applicable | 0.56 | 0.5 |
| sex_male | eICU-CRD | 0.0 | not applicable | not applicable | not applicable | 0.54 | 0.5 |
| spo2_last | MIMIC-III | 0.7 | 100.0 | 97.0 | 100.0 | 98.46 | 2.6 |
| spo2_last | MIMIC-IV | 0.1 | 97.0 | 95.0 | 99.0 | 96.47 | 2.92 |
| spo2_last | eICU-CRD | 2.2 | 97.0 | 95.0 | 99.0 | 96.42 | 3.54 |
| spo2_max | MIMIC-III | 0.7 | 100.0 | 100.0 | 100.0 | 99.83 | 0.79 |
| spo2_max | MIMIC-IV | 0.1 | 100.0 | 99.0 | 100.0 | 99.51 | 1.02 |
| spo2_max | eICU-CRD | 2.2 | 100.0 | 100.0 | 100.0 | 99.63 | 0.87 |
| spo2_min | MIMIC-III | 0.7 | 96.0 | 93.0 | 100.0 | 94.83 | 5.88 |
| spo2_min | MIMIC-IV | 0.1 | 93.0 | 91.0 | 95.0 | 92.2 | 4.69 |
| spo2_min | eICU-CRD | 2.2 | 90.0 | 85.0 | 93.0 | 87.7 | 8.41 |
| temp_c_last | MIMIC-III | 0.7 | 37.06 | 36.61 | 37.5 | 37.03 | 0.77 |
| temp_c_last | MIMIC-IV | 2.4 | 36.89 | 36.67 | 37.22 | 36.93 | 0.59 |
| temp_c_last | eICU-CRD | 88.9 | 37.25 | 36.8 | 37.7 | 37.05 | 1.22 |
| temp_c_max | MIMIC-III | 0.7 | 37.6 | 37.11 | 38.1 | 37.65 | 0.72 |
| temp_c_max | MIMIC-IV | 2.4 | 37.22 | 36.94 | 37.72 | 37.39 | 0.7 |
| temp_c_max | eICU-CRD | 88.9 | 37.8 | 37.3 | 38.3 | 37.78 | 0.96 |
| temp_c_min | MIMIC-III | 0.7 | 36.11 | 35.61 | 36.61 | 36.05 | 0.86 |
| temp_c_min | MIMIC-IV | 2.4 | 36.44 | 36.11 | 36.67 | 36.32 | 0.65 |
| temp_c_min | eICU-CRD | 88.9 | 36.0 | 35.2 | 36.7 | 35.63 | 1.64 |
| weight_kg_last | MIMIC-III | 41.1 | 81.0 | 68.0 | 95.7 | 83.9 | 23.94 |
| weight_kg_last | MIMIC-IV | 0.6 | 79.7 | 66.7 | 94.8 | 82.38 | 22.96 |
| weight_kg_last | eICU-CRD | 0.3 | 80.6 | 66.8 | 97.1 | 83.95 | 26.37 |
| weight_kg_max | MIMIC-III | 41.1 | 81.1 | 68.0 | 95.9 | 84.11 | 24.22 |
| weight_kg_max | MIMIC-IV | 0.6 | 79.83 | 67.0 | 95.0 | 82.77 | 23.15 |
| weight_kg_max | eICU-CRD | 1.8 | 81.64 | 68.04 | 98.6 | 85.67 | 26.71 |
| weight_kg_min | MIMIC-III | 41.1 | 80.5 | 67.8 | 95.0 | 83.43 | 23.75 |
| weight_kg_min | MIMIC-IV | 0.6 | 78.1 | 65.68 | 92.7 | 80.95 | 22.52 |
| weight_kg_min | eICU-CRD | 74.1 | 80.0 | 66.5 | 96.7 | 83.85 | 26.03 |

Median, first and third quartile, mean and standard deviation for every model input in every database, with the percentage missing before imputation. No feature exceeded 13 percent missingness in MIMIC-IV. Median and quartiles are not applicable to the binary sex indicator; its mean is the proportion male.

**Supplementary Table S2.** Standardized mean differences by variable.

Panel A. Summary over all 42 variables
| Pair | Variables | Mean | Median | Maximum |
| --- | --- | --- | --- | --- |
| MIMIC-IV vs MIMIC-III | 42 | 0.129 | 0.068 | 0.718 |
| MIMIC-IV vs eICU-CRD | 42 | 0.174 | 0.120 | 0.660 |
| MIMIC-III vs eICU-CRD | 42 | 0.207 | 0.149 | 0.982 |

Panel B. Every variable
| Variable | MIMIC-IV vs MIMIC-III | MIMIC-IV vs eICU-CRD | MIMIC-III vs eICU-CRD |
| --- | --- | --- | --- |
| spo2_min | 0.495 | 0.660 | 0.982 |
| gcs_total_min | 0.082 | 0.657 | 0.755 |
| temp_c_min | 0.358 | 0.556 | 0.320 |
| rr_max | 0.096 | 0.521 | 0.578 |
| temp_c_max | 0.364 | 0.466 | 0.157 |
| dbp_max | 0.408 | 0.377 | 0.770 |
| gcs_total_max | 0.072 | 0.292 | 0.228 |
| rr_min | 0.026 | 0.291 | 0.263 |
| hr_max | 0.049 | 0.279 | 0.232 |
| sbp_max | 0.070 | 0.213 | 0.145 |
| inr_min | 0.012 | 0.186 | 0.183 |
| dbp_last | 0.284 | 0.186 | 0.476 |
| inr_last | 0.016 | 0.166 | 0.158 |
| calcium_max | 0.115 | 0.166 | 0.048 |
| baseline_scr | 0.060 | 0.143 | 0.202 |
| scr_feat_min | 0.060 | 0.143 | 0.201 |
| bun_cr_ratio_max | 0.014 | 0.143 | 0.154 |
| temp_c_last | 0.145 | 0.134 | 0.028 |
| sbp_min | 0.011 | 0.130 | 0.138 |
| spo2_max | 0.352 | 0.125 | 0.243 |
| sbp_last | 0.056 | 0.121 | 0.065 |
| weight_kg_min | 0.107 | 0.119 | 0.017 |
| weight_kg_max | 0.057 | 0.116 | 0.061 |
| scr_feat_last | 0.064 | 0.112 | 0.174 |
| calcium_last | 0.072 | 0.111 | 0.038 |
| gcs_total_last | 0.086 | 0.110 | 0.025 |
| inr_max | 0.034 | 0.101 | 0.071 |
| rr_last | 0.037 | 0.099 | 0.131 |
| scr_feat_max | 0.084 | 0.079 | 0.163 |
| age | 0.089 | 0.070 | 0.020 |
| bun_min | 0.030 | 0.068 | 0.098 |
| bun_cr_ratio_min | 0.035 | 0.067 | 0.033 |
| weight_kg_last | 0.065 | 0.064 | 0.002 |
| bun_last | 0.038 | 0.051 | 0.088 |
| hr_min | 0.129 | 0.041 | 0.167 |
| hr_last | 0.089 | 0.035 | 0.051 |
| sex_male | 0.042 | 0.035 | 0.077 |
| calcium_min | 0.066 | 0.023 | 0.044 |
| bun_cr_ratio_last | 0.020 | 0.021 | 0.039 |
| bun_max | 0.049 | 0.020 | 0.068 |
| spo2_last | 0.718 | 0.016 | 0.657 |
| dbp_min | 0.356 | 0.007 | 0.323 |

**Supplementary Table S3.**
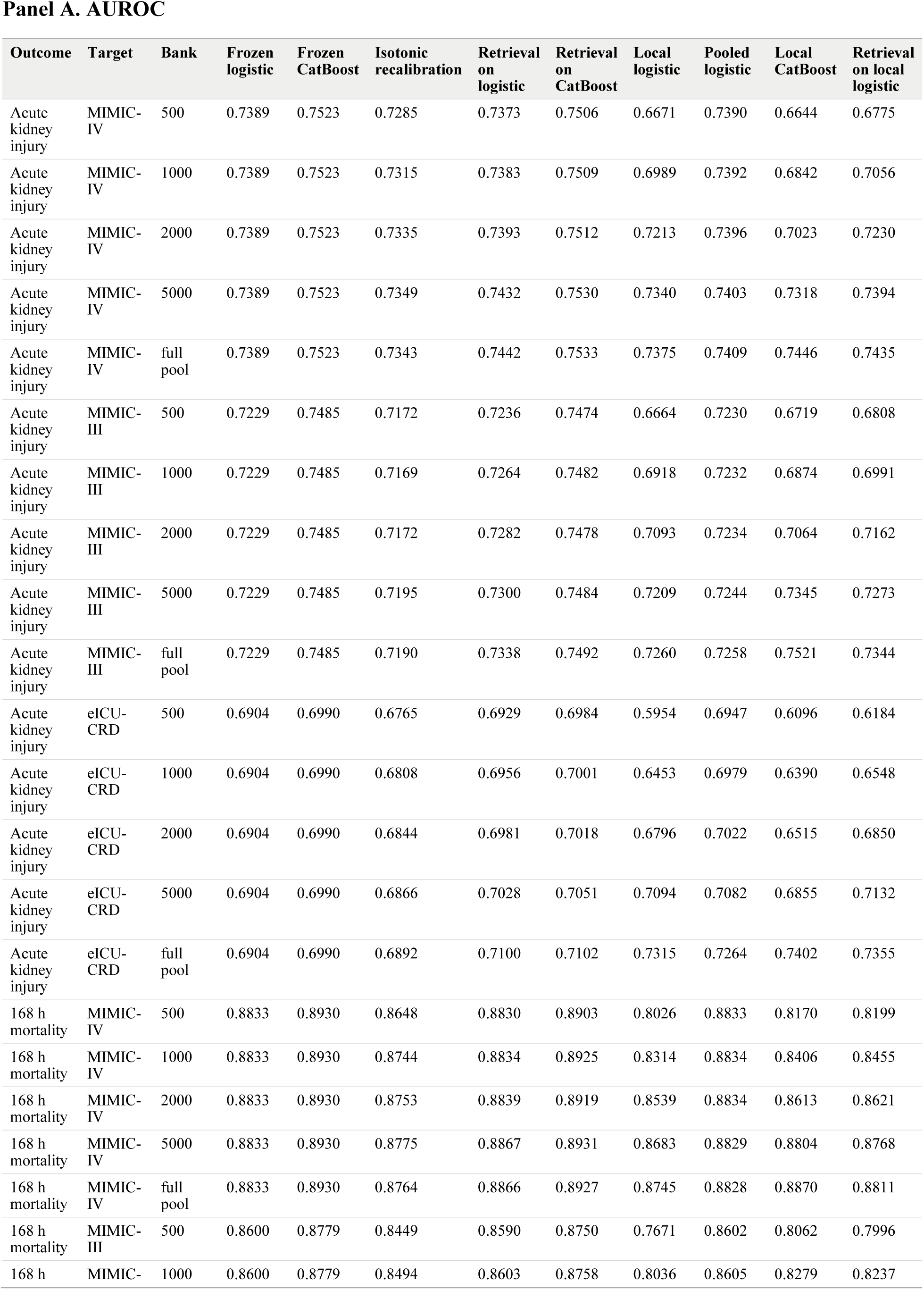
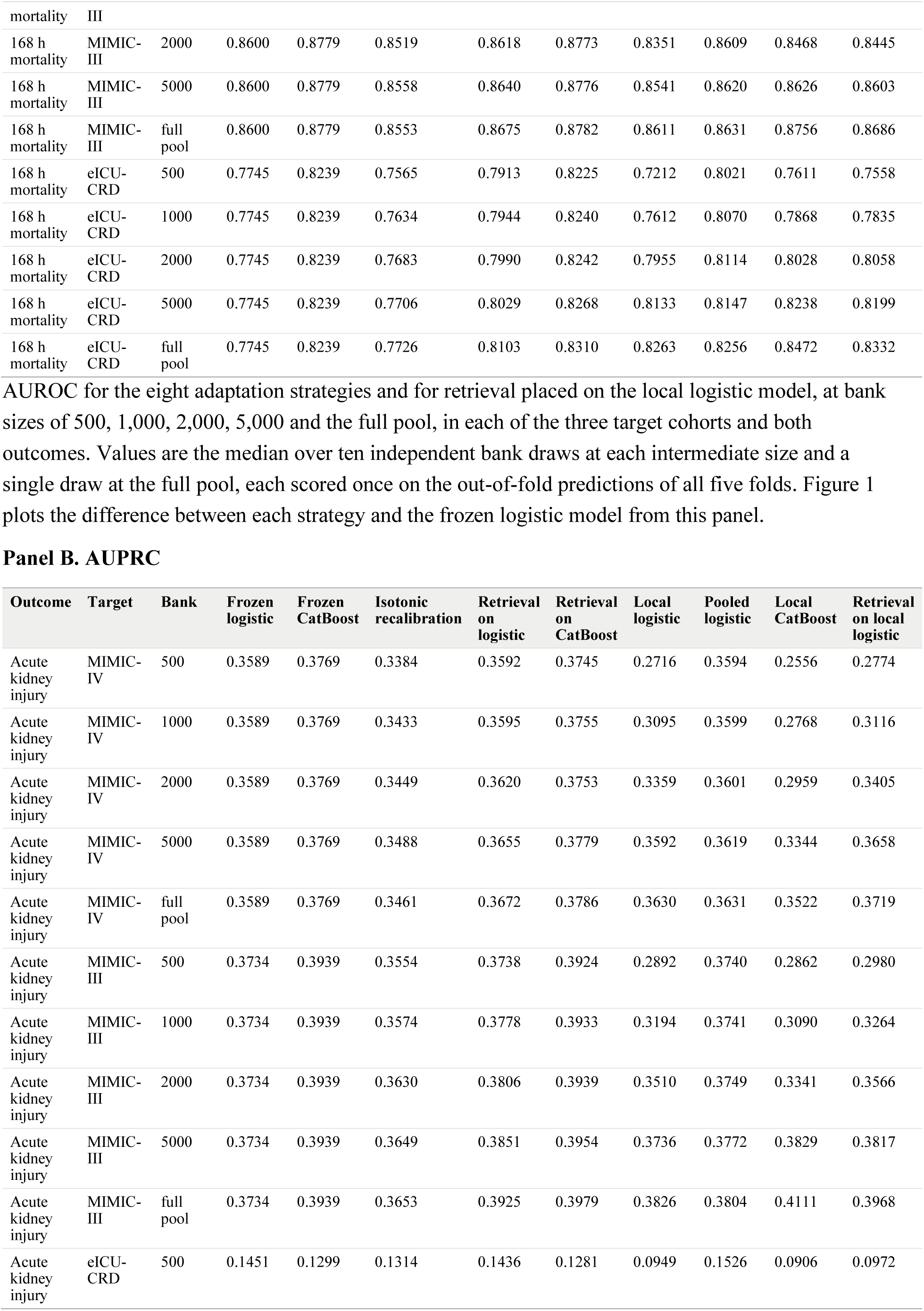
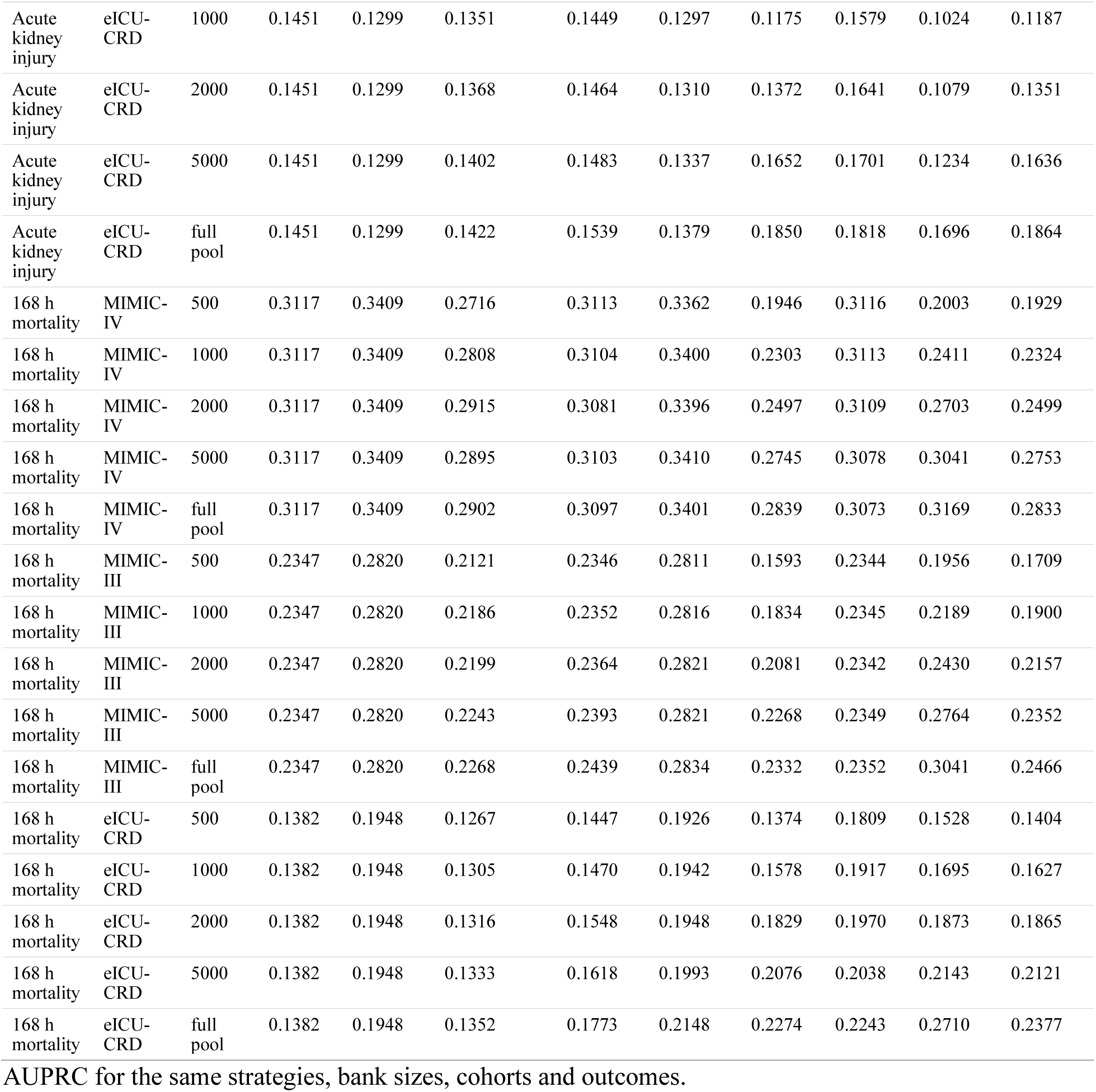
All eight strategies, and retrieval on the local logistic model, at every bank size.

**Supplementary Table S4.**
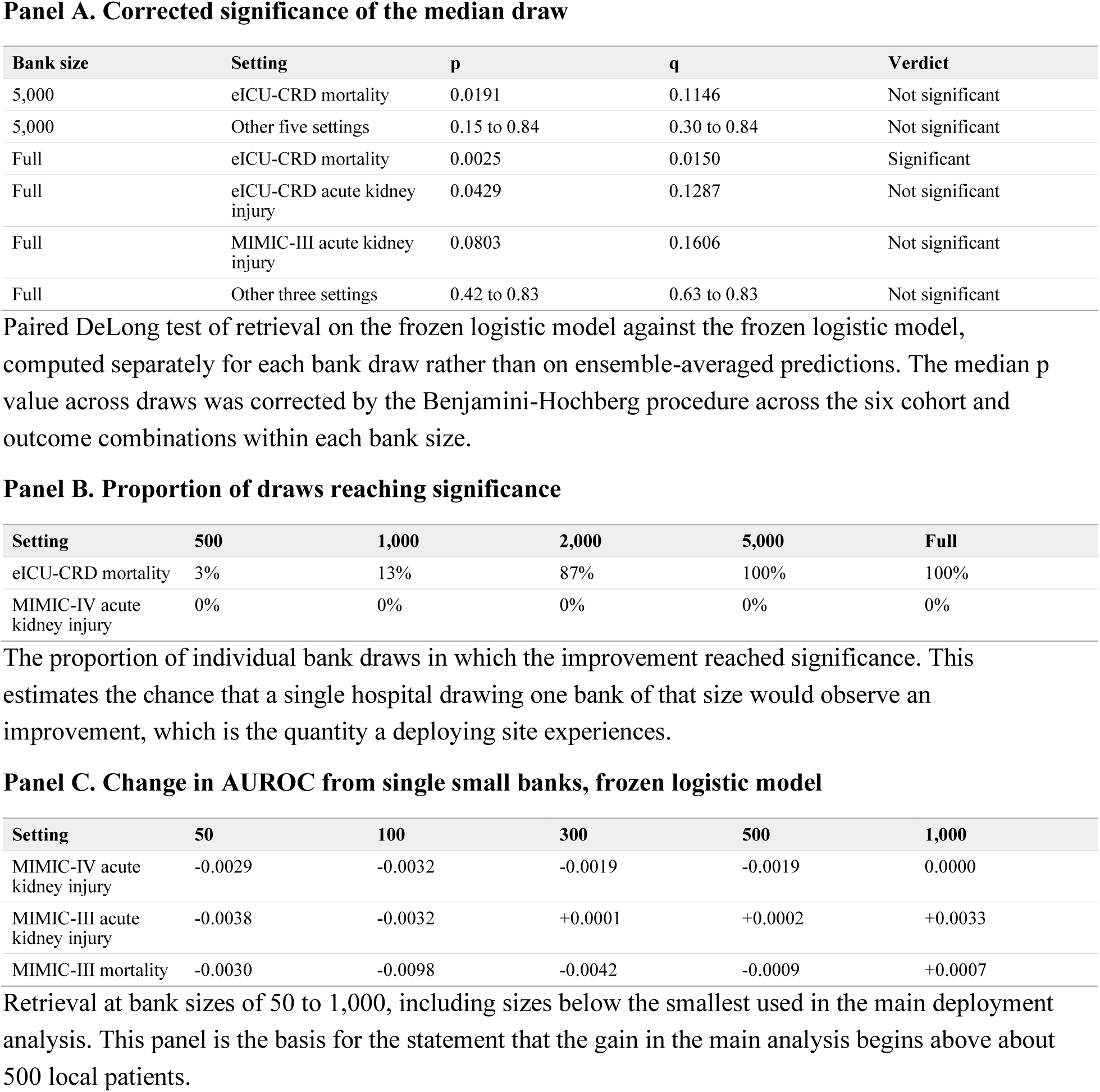
Proportion of bank draws with significant improvement.

**Supplementary Figure S1.**
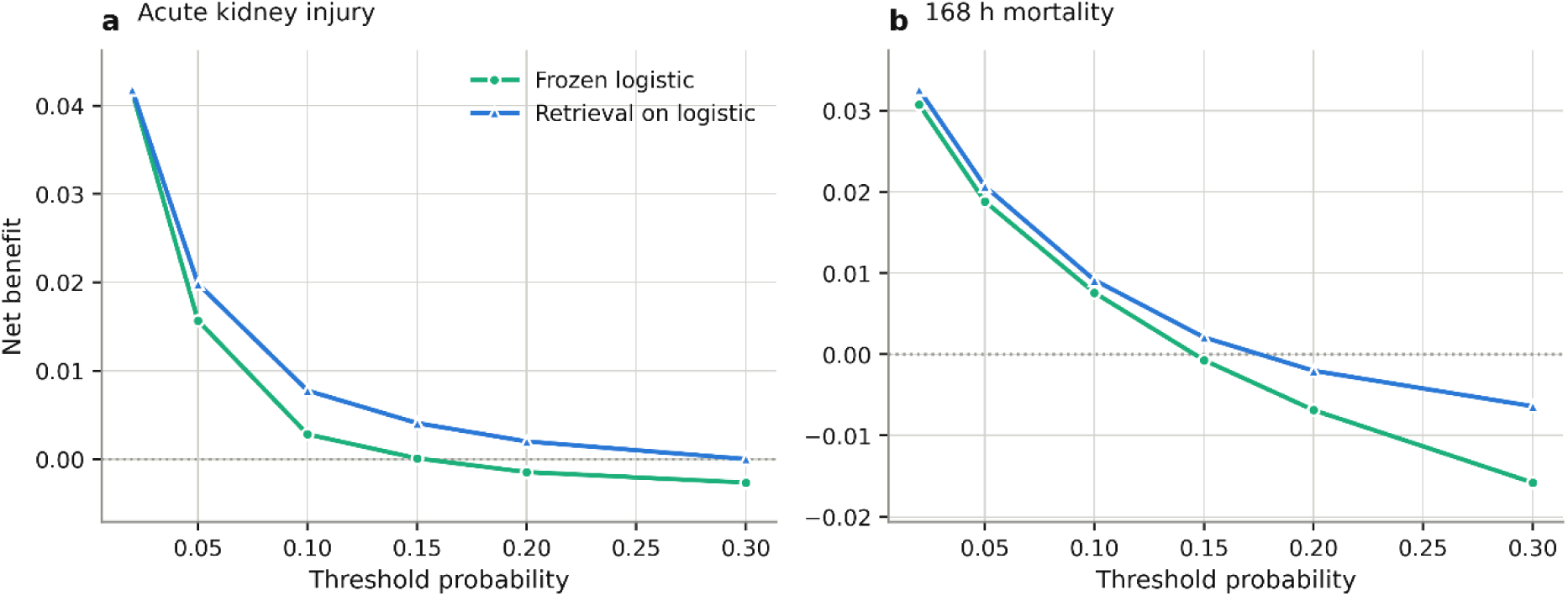
Decision curves, eICU-CRD. Net benefit against threshold probability from 2 to 30 percent for the frozen logistic model and for retrieval on it, at a bank of 5,000 patients in eICU-CRD, for acute kidney injury and 168 hour mortality. The two reference lines are treat all and treat none. A strategy is useful at a given threshold only where its curve lies above both reference lines. The frozen model falls below the treat none line above a threshold of roughly 0.15 for acute kidney injury, and retrieval keeps it at or above that line up to a threshold of 0.30. At thresholds of 0.10, 0.15, 0.20 and 0.30 the frozen logistic model reached +0.0028, +0.0001, -0.0015 and -0.0027 for acute kidney injury, and retrieval on it reached +0.0078, +0.0041, +0.0020 and 0.0000. Net benefit for all eight strategies at thresholds of 0.10 and 0.20 is in Table 3 and Supplementary Table S5.

**Supplementary Table S5.** Calibration and net benefit, MIMIC-IV and MIMIC-III.

Panel A. Calibration slope, where 1.00 is ideal
| Outcome | Target | Bank | Frozen logistic | Frozen CatBoost | Isotonic recalibration | Retrieval on logistic | Retrieval on CatBoost | Local logistic | Pooled logistic | Local CatBoost | Retrieval on local logistic |
| --- | --- | --- | --- | --- | --- | --- | --- | --- | --- | --- | --- |
| Acute kidney injury | MIMIC-IV | full pool | 1.034 | 0.984 | 0.952 | 1.196 | 1.053 | 0.983 | 1.031 | 1.035 | 1.179 |
| Acute kidney injury | MIMIC-III | full pool | 1.018 | 0.978 | 0.959 | 1.240 | 1.062 | 0.982 | 1.004 | 1.074 | 1.199 |
| 168 h mortality | MIMIC-IV | full pool | 1.006 | 0.981 | 0.932 | 1.094 | 1.003 | 0.916 | 0.994 | 1.028 | 1.053 |
| 168 h mortality | MIMIC-III | full pool | 0.907 | 0.894 | 0.945 | 1.030 | 0.920 | 0.961 | 0.949 | 1.045 | 1.116 |

Panel B. Calibration intercept, where 0.00 is ideal
| Outcome | Target | Bank | Frozen logistic | Frozen CatBoost | Isotonic recalibration | Retrieval on logistic | Retrieval on CatBoost | Local logistic | Pooled logistic | Local CatBoost | Retrieval on local logistic |
| --- | --- | --- | --- | --- | --- | --- | --- | --- | --- | --- | --- |
| Acute kidney injury | MIMIC-IV | full pool | 0.060 | 0.009 | -0.073 | 0.312 | 0.110 | -0.027 | 0.053 | 0.113 | 0.277 |
| Acute kidney injury | MIMIC-III | full pool | 0.114 | 0.044 | -0.061 | 0.441 | 0.164 | -0.028 | 0.055 | 0.141 | 0.310 |
| 168 h mortality | MIMIC-IV | full pool | 0.017 | 0.068 | -0.145 | 0.192 | 0.105 | -0.183 | -0.012 | 0.251 | 0.097 |
| 168 h mortality | MIMIC-III | full pool | 0.050 | 0.066 | -0.119 | 0.294 | 0.112 | -0.090 | 0.003 | 0.209 | 0.265 |

Panel C. Expected calibration error
| Outcome | Target | Bank | Frozen logistic | Frozen CatBoost | Isotonic recalibration | Retrieval on logistic | Retrieval on CatBoost | Local logistic | Pooled logistic | Local CatBoost | Retrieval on local logistic |
| --- | --- | --- | --- | --- | --- | --- | --- | --- | --- | --- | --- |
| Acute kidney injury | MIMIC-IV | full pool | 0.0105 | 0.0045 | 0.0044 | 0.0197 | 0.0053 | 0.0073 | 0.0093 | 0.0073 | 0.0165 |
| Acute kidney injury | MIMIC-III | full pool | 0.0112 | 0.0093 | 0.0034 | 0.0204 | 0.0085 | 0.0098 | 0.0082 | 0.0093 | 0.0172 |
| 168 h mortality | MIMIC-IV | full pool | 0.0063 | 0.0051 | 0.0032 | 0.0061 | 0.0045 | 0.0063 | 0.0060 | 0.0075 | 0.0060 |
| 168 h mortality | MIMIC-III | full pool | 0.0144 | 0.0126 | 0.0033 | 0.0109 | 0.0120 | 0.0069 | 0.0101 | 0.0048 | 0.0087 |

Panel D. Net benefit at a threshold probability of 10 percent
| Outcome | Target | Bank | Frozen logistic | Frozen CatBoost | Isotonic recalibration | Retrieval on logistic | Retrieval on CatBoost | Local logistic | Pooled logistic | Local CatBoost | Retrieval on local logistic |
| --- | --- | --- | --- | --- | --- | --- | --- | --- | --- | --- | --- |
| Acute kidney injury | MIMIC-IV | full pool | 0.0702 | 0.0729 | 0.0709 | 0.0702 | 0.0735 | 0.0698 | 0.0716 | 0.0727 | 0.0708 |
| Acute kidney injury | MIMIC-III | full pool | 0.0792 | 0.0855 | 0.0791 | 0.0814 | 0.0859 | 0.0795 | 0.0796 | 0.0871 | 0.0818 |
| 168 h mortality | MIMIC-IV | full pool | 0.0173 | 0.0175 | 0.0171 | 0.0172 | 0.0173 | 0.0160 | 0.0171 | 0.0162 | 0.0162 |
| 168 h mortality | MIMIC-III | full pool | 0.0152 | 0.0166 | 0.0151 | 0.0162 | 0.0166 | 0.0162 | 0.0160 | 0.0177 | 0.0173 |

Panel E. Net benefit at a threshold probability of 20 percent
| Outcome | Target | Bank | Frozen logistic | Frozen CatBoost | Isotonic recalibration | Retrieval on logistic | Retrieval on CatBoost | Local logistic | Pooled logistic | Local CatBoost | Retrieval on local logistic |
| --- | --- | --- | --- | --- | --- | --- | --- | --- | --- | --- | --- |
| Acute kidney injury | MIMIC-IV | full pool | 0.0398 | 0.0398 | 0.0403 | 0.0413 | 0.0393 | 0.0394 | 0.0399 | 0.0374 | 0.0404 |
| Acute kidney injury | MIMIC-III | full pool | 0.0435 | 0.0473 | 0.0434 | 0.0467 | 0.0474 | 0.0440 | 0.0440 | 0.0463 | 0.0453 |
| 168 h mortality | MIMIC-IV | full pool | 0.0077 | 0.0096 | 0.0071 | 0.0078 | 0.0095 | 0.0074 | 0.0074 | 0.0083 | 0.0073 |
| 168 h mortality | MIMIC-III | full pool | 0.0058 | 0.0088 | 0.0061 | 0.0061 | 0.0087 | 0.0060 | 0.0056 | 0.0081 | 0.0062 |

Net benefit at a threshold probability of 20 percent. These two cohorts are closer to the development population than eICU-CRD, so the frozen models are less miscalibrated and the strategies separate less.

**Supplementary Table S6.**
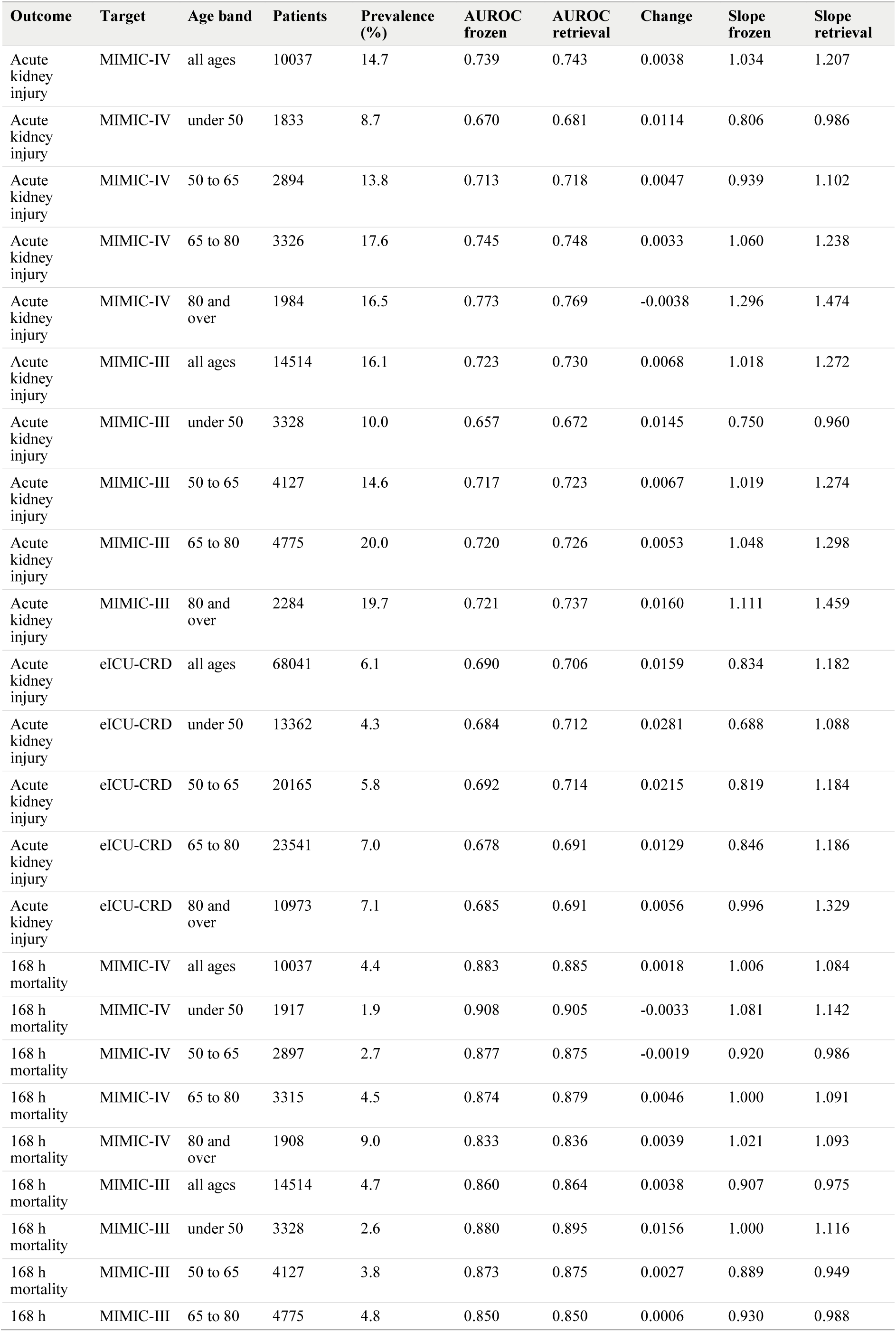
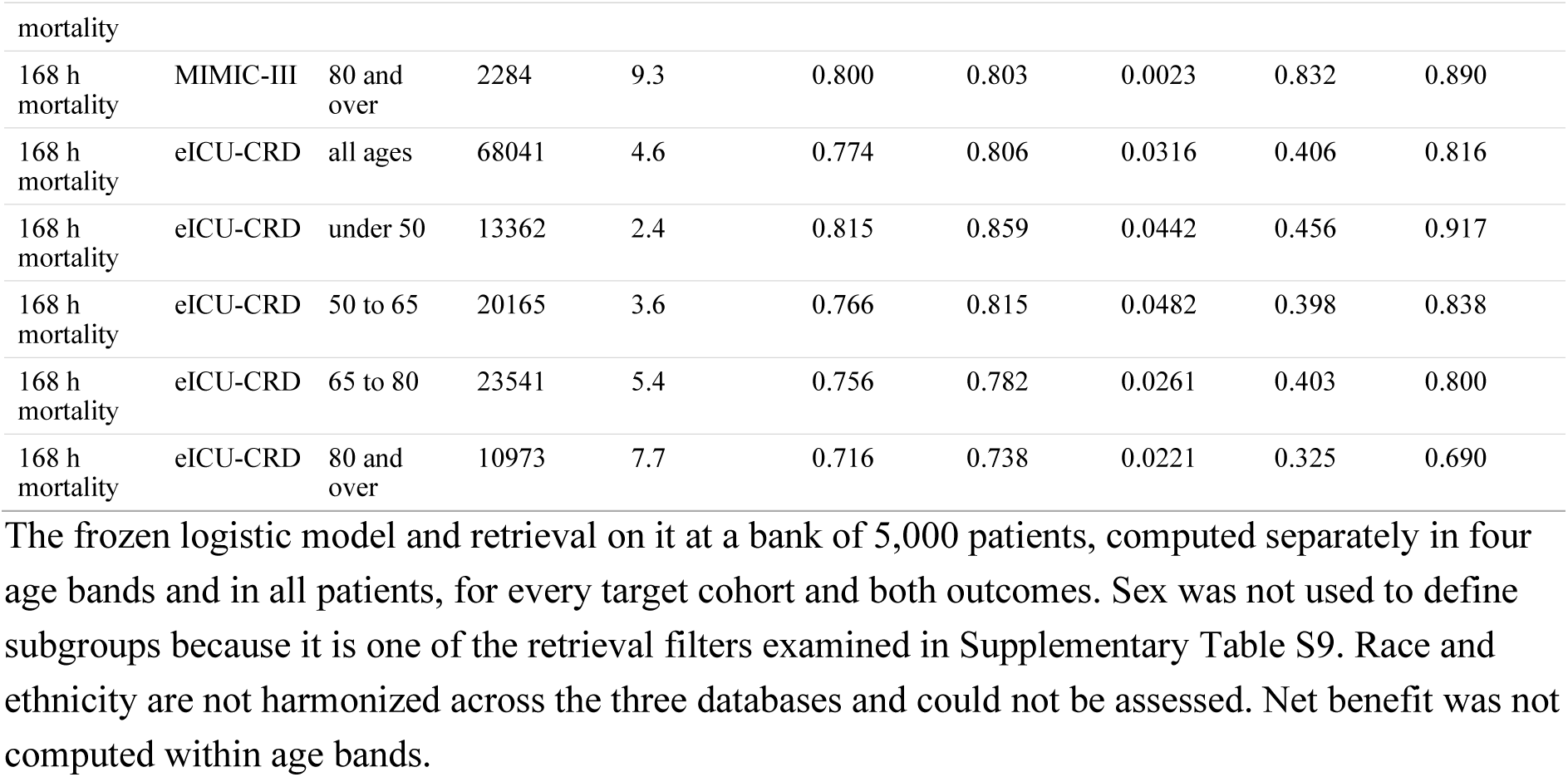
Age subgroups.

**Supplementary Table S7.** Bank composition, all rules and sizes.

| Outcome | Target | Bank rule | Bank size | Bank prevalence (%) | Mixing weight | Change in AUROC | SD |
| --- | --- | --- | --- | --- | --- | --- | --- |
| Acute kidney injury | MIMIC-III | Cluster representatives | 300 | 15.3 | 0.10 | -0.0013 | 0.0028 |
| Acute kidney injury | MIMIC-III | Cluster representatives | 1000 | 17.2 | 0.23 | 0.0027 | 0.0025 |
| Acute kidney injury | MIMIC-III | Cluster representatives | 5000 | 17.3 | 0.37 | 0.0074 | 0.0046 |
| Acute kidney injury | MIMIC-III | Coverage of the feature space | 300 | 22.0 | 0.17 | 0.0020 | 0.0022 |
| Acute kidney injury | MIMIC-III | Coverage of the feature space | 1000 | 20.5 | 0.20 | 0.0009 | 0.0044 |
| Acute kidney injury | MIMIC-III | Coverage of the feature space | 5000 | 17.3 | 0.44 | 0.0106 | 0.0047 |
| Acute kidney injury | MIMIC-III | Least certain under the frozen model | 300 | 53.5 | 0.40 | -0.0192 | 0.0037 |
| Acute kidney injury | MIMIC-III | Least certain under the frozen model | 1000 | 47.3 | 0.50 | -0.0141 | 0.0201 |
| Acute kidney injury | MIMIC-III | Least certain under the frozen model | 5000 | 25.9 | 0.40 | 0.0065 | 0.0050 |
| Acute kidney injury | MIMIC-III | Most recent | 300 | 17.2 | 0.15 | 0.0019 | 0.0036 |
| Acute kidney injury | MIMIC-III | Most recent | 1000 | 16.8 | 0.27 | 0.0065 | 0.0023 |
| Acute kidney injury | MIMIC-III | Most recent | 5000 | 16.2 | 0.35 | 0.0087 | 0.0026 |
| Acute kidney injury | MIMIC-III | Prevalence balanced | 300 | 50.0 | 0.21 | 0.0031 | 0.0039 |
| Acute kidney injury | MIMIC-III | Prevalence balanced | 1000 | 50.0 | 0.22 | 0.0047 | 0.0038 |
| Acute kidney injury | MIMIC-III | Prevalence balanced | 5000 | 37.4 | 0.36 | 0.0107 | 0.0032 |
| Acute kidney injury | MIMIC-III | Random | 300 | 16.0 | 0.20 | 0.0011 | 0.0012 |
| Acute kidney injury | MIMIC-III | Random | 1000 | 16.1 | 0.25 | 0.0026 | 0.0025 |
| Acute kidney injury | MIMIC-III | Random | 5000 | 16.1 | 0.43 | 0.0080 | 0.0043 |
| Acute kidney injury | eICU-CRD | Cluster representatives | 300 | 7.6 | 0.28 | 0.0035 | 0.0060 |
| Acute kidney injury | eICU-CRD | Cluster representatives | 1000 | 8.6 | 0.40 | 0.0130 | 0.0022 |
| Acute kidney injury | eICU-CRD | Cluster representatives | 5000 | 8.4 | 0.50 | 0.0228 | 0.0035 |
| Acute kidney injury | eICU-CRD | Coverage of the feature space | 300 | 13.8 | 0.36 | -0.0220 | 0.0158 |
| Acute kidney injury | eICU-CRD | Coverage of the feature space | 1000 | 12.3 | 0.43 | 0.0022 | 0.0063 |
| Acute kidney injury | eICU-CRD | Coverage of the feature space | 5000 | 9.6 | 0.49 | 0.0210 | 0.0053 |
| Acute kidney injury | eICU-CRD | Least certain under the frozen model | 300 | 26.1 | 0.47 | 0.0021 | 0.0195 |
| Acute kidney injury | eICU-CRD | Least certain under the frozen model | 1000 | 25.8 | 0.50 | -0.0210 | 0.0090 |
| Acute kidney injury | eICU-CRD | Least certain under the frozen model | 5000 | 17.9 | 0.49 | 0.0148 | 0.0079 |
| Acute kidney injury | eICU-CRD | Most recent | 300 | 5.9 | 0.34 | 0.0025 | 0.0059 |
| Acute kidney injury | eICU-CRD | Most recent | 1000 | 6.4 | 0.35 | 0.0071 | 0.0049 |
| Acute kidney injury | eICU-CRD | Most recent | 5000 | 6.5 | 0.50 | 0.0175 | 0.0039 |
| Acute kidney injury | eICU-CRD | Prevalence balanced | 300 | 50.0 | 0.16 | 0.0049 | 0.0071 |
| Acute kidney injury | eICU-CRD | Prevalence balanced | 1000 | 50.0 | 0.27 | 0.0194 | 0.0026 |
| Acute kidney injury | eICU-CRD | Prevalence balanced | 5000 | 50.0 | 0.29 | 0.0254 | 0.0030 |
| Acute kidney injury | eICU-CRD | Random | 300 | 6.0 | 0.29 | 0.0032 | 0.0032 |
| Acute kidney injury | eICU-CRD | Random | 1000 | 6.1 | 0.23 | 0.0049 | 0.0048 |
| Acute kidney injury | eICU-CRD | Random | 5000 | 6.1 | 0.45 | 0.0175 | 0.0053 |
| 168 h mortality | MIMIC-III | Cluster representatives | 300 | 6.9 | 0.10 | -0.0007 | 0.0025 |
| 168 h mortality | MIMIC-III | Cluster representatives | 1000 | 8.9 | 0.17 | 0.0016 | 0.0031 |
| 168 h mortality | MIMIC-III | Cluster representatives | 5000 | 8.1 | 0.19 | 0.0059 | 0.0038 |
| 168 h mortality | MIMIC-III | Coverage of the feature space | 300 | 18.3 | 0.19 | -0.0063 | 0.0098 |
| 168 h mortality | MIMIC-III | Coverage of the feature space | 1000 | 14.5 | 0.22 | -0.0011 | 0.0016 |
| 168 h mortality | MIMIC-III | Coverage of the feature space | 5000 | 8.5 | 0.20 | 0.0058 | 0.0036 |
| 168 h mortality | MIMIC-III | Least certain under the frozen model | 300 | 32.3 | 0.47 | -0.1252 | 0.0453 |
| 168 h mortality | MIMIC-III | Least certain under the frozen model | 1000 | 25.5 | 0.43 | -0.0558 | 0.0444 |
| 168 h mortality | MIMIC-III | Least certain under the frozen model | 5000 | 10.1 | 0.27 | 0.0030 | 0.0044 |
| 168 h mortality | MIMIC-III | Most recent | 300 | 4.3 | 0.08 | 0.0001 | 0.0001 |
| 168 h mortality | MIMIC-III | Most recent | 1000 | 4.4 | 0.15 | 0.0013 | 0.0023 |
| 168 h mortality | MIMIC-III | Most recent | 5000 | 4.8 | 0.11 | 0.0031 | 0.0021 |
| 168 h mortality | MIMIC-III | Prevalence balanced | 300 | 50.0 | 0.05 | 0.0021 | 0.0012 |
| 168 h mortality | MIMIC-III | Prevalence balanced | 1000 | 50.0 | 0.04 | 0.0023 | 0.0027 |
| 168 h mortality | MIMIC-III | Prevalence balanced | 5000 | 11.0 | 0.09 | 0.0044 | 0.0032 |
| 168 h mortality | MIMIC-III | Random | 300 | 4.7 | 0.10 | -0.0009 | 0.0031 |
| 168 h mortality | MIMIC-III | Random | 1000 | 4.7 | 0.12 | 0.0005 | 0.0019 |
| 168 h mortality | MIMIC-III | Random | 5000 | 4.7 | 0.14 | 0.0034 | 0.0026 |
| 168 h mortality | eICU-CRD | Cluster representatives | 300 | 8.5 | 0.27 | 0.0089 | 0.0061 |
| 168 h mortality | eICU-CRD | Cluster representatives | 1000 | 8.7 | 0.43 | 0.0251 | 0.0054 |
| 168 h mortality | eICU-CRD | Cluster representatives | 5000 | 9.1 | 0.50 | 0.0323 | 0.0033 |
| 168 h mortality | eICU-CRD | Coverage of the feature space | 300 | 22.5 | 0.50 | 0.0060 | 0.0082 |
| 168 h mortality | eICU-CRD | Coverage of the feature space | 1000 | 18.1 | 0.50 | 0.0117 | 0.0096 |
| 168 h mortality | eICU-CRD | Coverage of the feature space | 5000 | 10.1 | 0.50 | 0.0316 | 0.0036 |
| 168 h mortality | eICU-CRD | Least certain under the frozen model | 300 | 15.0 | 0.50 | 0.0220 | 0.0103 |
| 168 h mortality | eICU-CRD | Least certain under the frozen model | 1000 | 15.8 | 0.50 | 0.0253 | 0.0078 |
| 168 h mortality | eICU-CRD | Least certain under the frozen model | 5000 | 15.8 | 0.50 | 0.0378 | 0.0070 |
| 168 h mortality | eICU-CRD | Most recent | 300 | 5.1 | 0.28 | 0.0094 | 0.0087 |
| 168 h mortality | eICU-CRD | Most recent | 1000 | 5.0 | 0.50 | 0.0149 | 0.0025 |
| 168 h mortality | eICU-CRD | Most recent | 5000 | 5.5 | 0.50 | 0.0267 | 0.0029 |
| 168 h mortality | eICU-CRD | Prevalence balanced | 300 | 50.0 | 0.40 | 0.0258 | 0.0083 |
| 168 h mortality | eICU-CRD | Prevalence balanced | 1000 | 50.0 | 0.50 | 0.0398 | 0.0071 |
| 168 h mortality | eICU-CRD | Prevalence balanced | 5000 | 50.0 | 0.50 | 0.0444 | 0.0065 |
| 168 h mortality | eICU-CRD | Random | 300 | 4.7 | 0.38 | 0.0092 | 0.0097 |
| 168 h mortality | eICU-CRD | Random | 1000 | 4.6 | 0.40 | 0.0194 | 0.0083 |
| 168 h mortality | eICU-CRD | Random | 5000 | 4.6 | 0.50 | 0.0300 | 0.0049 |

**Supplementary Table S8.** Bank recency.

| Target | Outcome | Bank | Oldest first | Newest first | Random |
| --- | --- | --- | --- | --- | --- |
| eICU-CRD | Acute kidney injury | 1,000 | +0.0058 | +0.0159 | +0.0080 |
| eICU-CRD | 168 h mortality | 1,000 | +0.0155 | +0.0224 | +0.0119 |
| MIMIC-III | Acute kidney injury | 1,000 | +0.0001 | +0.0101 | 0.0000 |
| MIMIC-III | Acute kidney injury | 2,000 | +0.0036 | +0.0110 | +0.0006 |

Change in AUROC when a bank of fixed size is filled from the oldest end of the pool, from the newest end, or at random. A site can act on recency with the admission date alone, by discarding old records.

**Supplementary Table S9.**
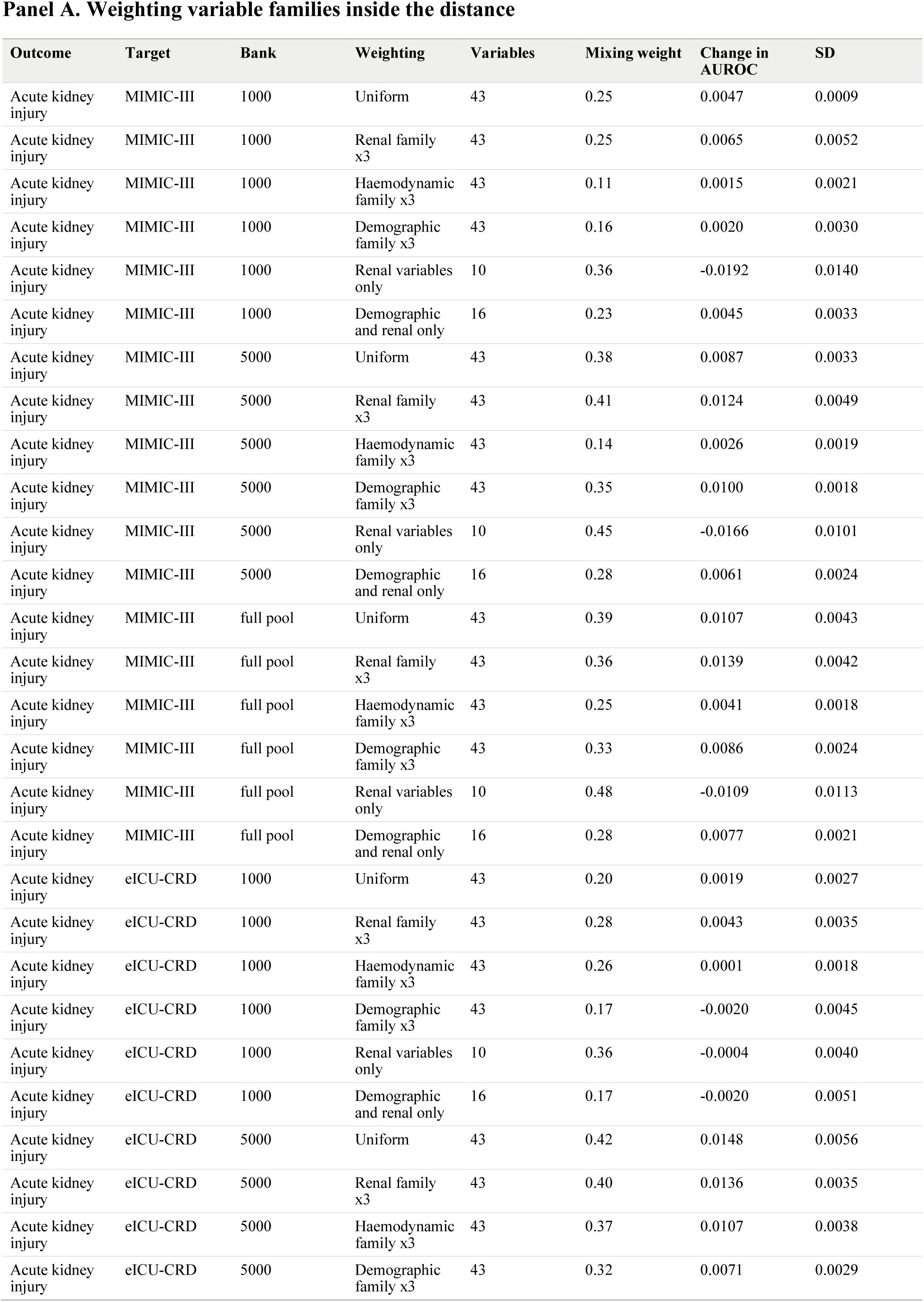
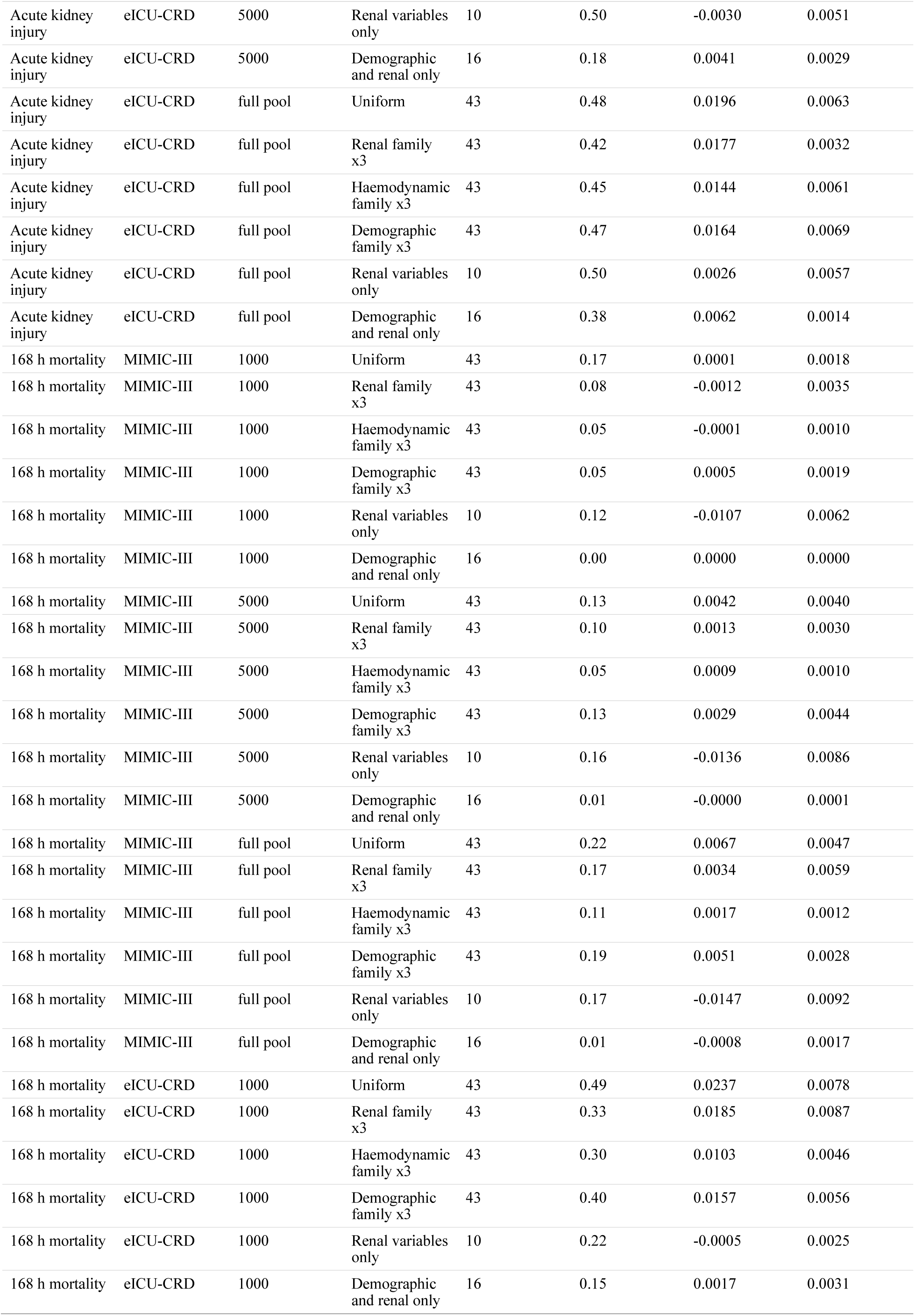
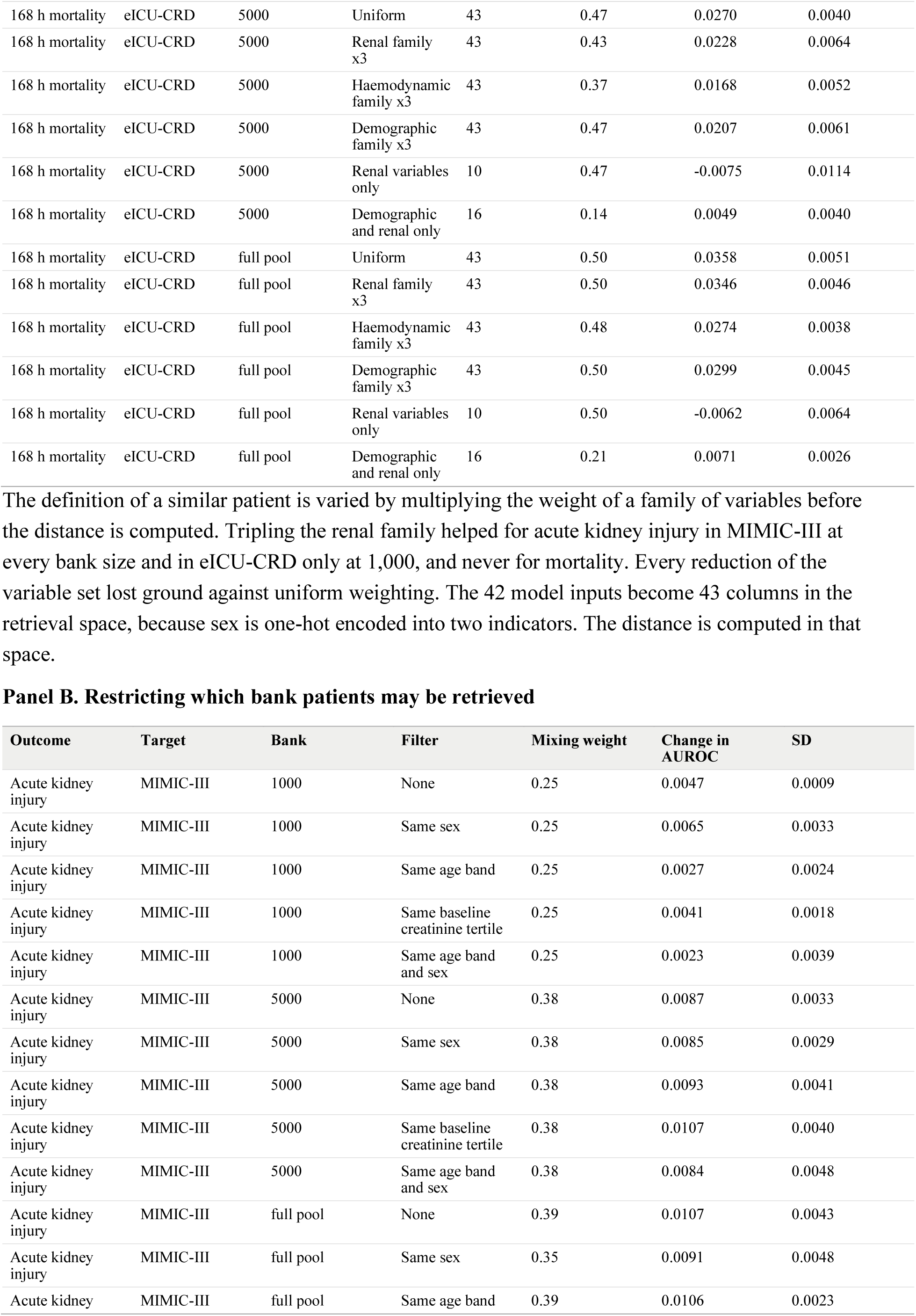
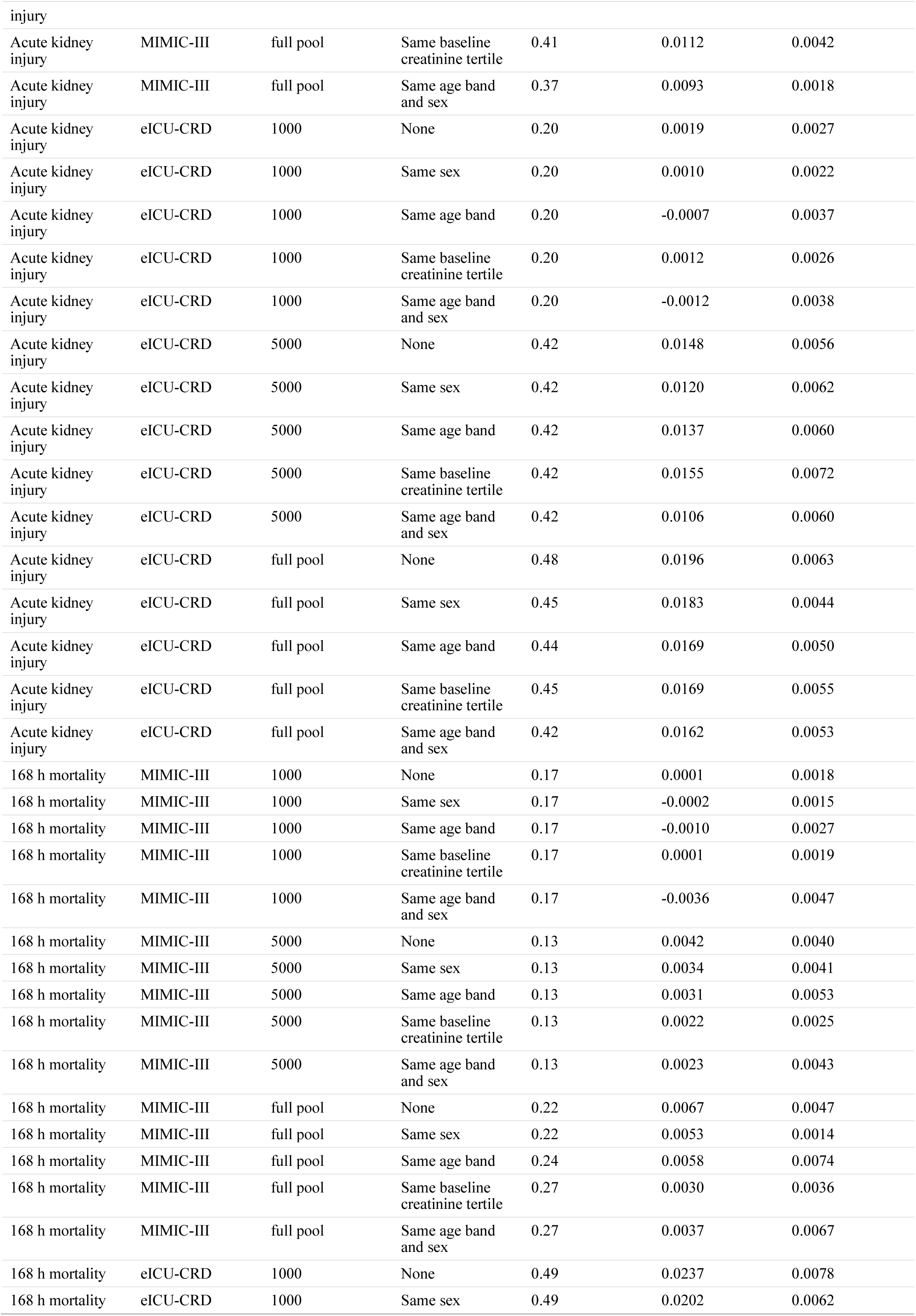
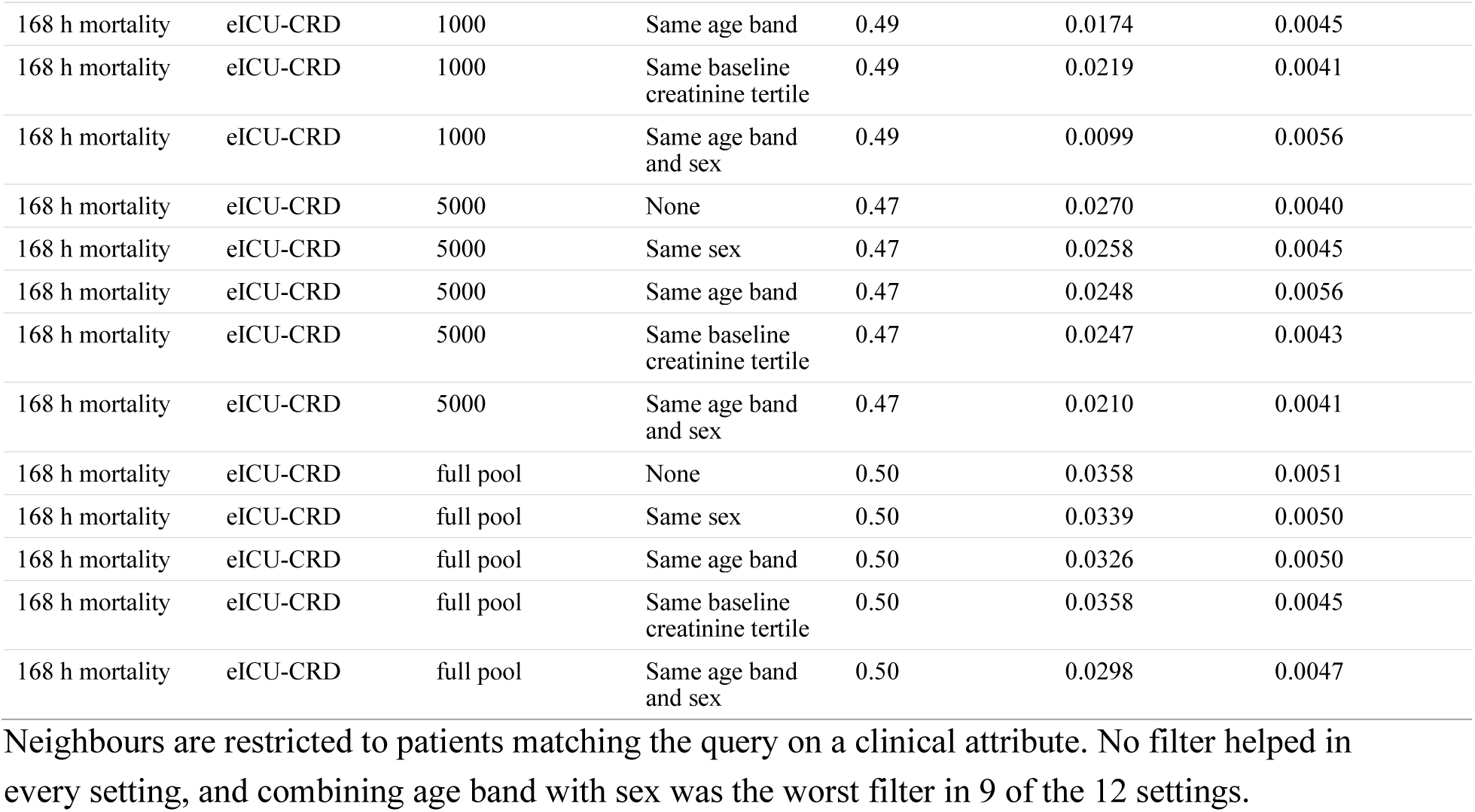
Similarity weighting and neighbour filters.

**Supplementary Table S10.** Base model strength and retrieval gain, ten development splits.

Panel A. Correlation between base model strength and retrieval gain
| Outcome | Units | Mean rho | SD | Range | Significance |
| --- | --- | --- | --- | --- | --- |
| Acute kidney injury | 5 base models | -0.727 | 0.121 | -0.90 to -0.50 | Mostly not significant |
| Acute kidney injury | 15 combinations | -0.692 | 0.105 | -0.86 to -0.51 | $p < 0.01$ in most splits |
| 168 h mortality | 5 base models | -0.800 | 0.141 | -1.00 to -0.50 | Significant in half of splits |
| 168 h mortality | 15 combinations | -0.836 | 0.062 | -0.93 to -0.77 | $p < 0.001$ in most splits |

Panel B. Each base model family with cosine retrieval
| Outcome | Base model | Base AUROC | Change from retrieval | Mixing weight |
| --- | --- | --- | --- | --- |
| Acute kidney injury | Logistic regression | 0.7368 | +0.0059 | 0.29 |
| Acute kidney injury | Random forest, small | 0.7286 | +0.0036 | 0.23 |
| Acute kidney injury | Random forest, full | 0.7298 | +0.0044 | 0.35 |
| Acute kidney injury | XGBoost | 0.7510 | +0.0003 | 0.15 |
| Acute kidney injury | CatBoost | 0.7539 | -0.0004 | 0.09 |
| 168 h mortality | Logistic regression | 0.8842 | +0.0040 | 0.27 |
| 168 h mortality | Random forest, small | 0.8742 | +0.0072 | 0.30 |
| 168 h mortality | Random forest, full | 0.8786 | +0.0074 | 0.34 |
| 168 h mortality | XGBoost | 0.8873 | +0.0013 | 0.16 |
| 168 h mortality | CatBoost | 0.8928 | +0.0001 | 0.13 |

**Supplementary Table S11.** Deployment with MIMIC-III as the development source.

| Development source | Outcome | Target | Bank | Frozen logistic | Frozen CatBoost | Retrieval on logistic | Retrieval minus frozen logistic |
| --- | --- | --- | --- | --- | --- | --- | --- |
| MIMIC-III | Acute kidney injury | MIMIC-IV | 0 | 0.7253 | 0.7319 | 0.7253 | 0.0000 |
| MIMIC-III | Acute kidney injury | MIMIC-IV | 1000 | 0.7253 | 0.7319 | 0.7283 | 0.0030 |
| MIMIC-III | Acute kidney injury | MIMIC-IV | 5000 | 0.7253 | 0.7319 | 0.7379 | 0.0126 |
| MIMIC-III | Acute kidney injury | MIMIC-IV | full pool | 0.7253 | 0.7319 | 0.7420 | 0.0167 |
| MIMIC-III | Acute kidney injury | eICU-CRD | 0 | 0.6703 | 0.6514 | 0.6703 | 0.0000 |
| MIMIC-III | Acute kidney injury | eICU-CRD | 1000 | 0.6703 | 0.6514 | 0.6918 | 0.0215 |
| MIMIC-III | Acute kidney injury | eICU-CRD | 5000 | 0.6703 | 0.6514 | 0.6957 | 0.0253 |
| MIMIC-III | Acute kidney injury | eICU-CRD | full pool | 0.6703 | 0.6514 | 0.7046 | 0.0343 |
| MIMIC-III | Acute kidney injury | MIMIC-III, internal | 0 | 0.7403 | 0.7549 | 0.7403 | 0.0000 |
| MIMIC-III | Acute kidney injury | MIMIC-III, internal | 1000 | 0.7403 | 0.7549 | 0.7451 | 0.0048 |
| MIMIC-III | Acute kidney injury | MIMIC-III, internal | 5000 | 0.7403 | 0.7549 | 0.7411 | 0.0007 |
| MIMIC-III | Acute kidney injury | MIMIC-III, internal | full pool | 0.7403 | 0.7549 | 0.7454 | 0.0050 |
| MIMIC-III | 168 h mortality | MIMIC-IV | 0 | 0.8738 | 0.8820 | 0.8738 | 0.0000 |
| MIMIC-III | 168 h mortality | MIMIC-IV | 1000 | 0.8738 | 0.8820 | 0.8753 | 0.0015 |
| MIMIC-III | 168 h mortality | MIMIC-IV | 5000 | 0.8738 | 0.8820 | 0.8790 | 0.0051 |
| MIMIC-III | 168 h mortality | MIMIC-IV | full pool | 0.8738 | 0.8820 | 0.8822 | 0.0084 |
| MIMIC-III | 168 h mortality | eICU-CRD | 0 | 0.7688 | 0.8152 | 0.7688 | 0.0000 |
| MIMIC-III | 168 h mortality | eICU-CRD | 1000 | 0.7688 | 0.8152 | 0.7916 | 0.0228 |
| MIMIC-III | 168 h mortality | eICU-CRD | 5000 | 0.7688 | 0.8152 | 0.7995 | 0.0307 |
| MIMIC-III | 168 h mortality | eICU-CRD | full pool | 0.7688 | 0.8152 | 0.8059 | 0.0371 |
| MIMIC-III | 168 h mortality | MIMIC-III, internal | 0 | 0.8615 | 0.8658 | 0.8615 | 0.0000 |
| MIMIC-III | 168 h mortality | MIMIC-III, internal | 1000 | 0.8615 | 0.8658 | 0.8603 | -0.0012 |
| MIMIC-III | 168 h mortality | MIMIC-III, internal | 5000 | 0.8615 | 0.8658 | 0.8523 | -0.0092 |
| MIMIC-III | 168 h mortality | MIMIC-III, internal | full pool | 0.8615 | 0.8658 | 0.8614 | -0.0001 |

AUROC at bank sizes of none, 1,000, 5,000 and the full pool when the frozen models are trained on MIMIC-III, which is smaller and older than MIMIC-IV, and deployed to MIMIC-IV, eICU-CRD and the MIMIC-III internal test split. Reversing the development source gives six further deployment scenarios and tests whether the relationship between base model strength and retrieval gain holds at cohort level.

**Supplementary Table S12.** Adaptive mixing weight and abstention.

Panel A. Settings in which the adaptive weight improved discrimination by more than 0.001 AUROC
| Settings examined | Adaptive weight better |
| --- | --- |
| 18 | 1 (eICU-CRD mortality at a bank of 1,000, +0.005) |

Panel B. Proportion of patients whose prediction was made worse
| Setting | Fixed weight | Adaptive weight |
| --- | --- | --- |
| MIMIC-IV acute kidney injury, bank 5,000 | 49.1% | 26.0% |
| MIMIC-IV acute kidney injury, full bank | 47.1% | 28.3% |
| MIMIC-III mortality, full bank | 34.2% | 29.1% |

The proportion of patients whose predicted probability moved away from their observed outcome. The adaptive weight did not improve discrimination and it halved this proportion. Any probability adjustment moves some patients in the wrong direction, so this is reported as a property of the mechanism and not as a claim about individual patients.

**Supplementary Table S13.** Learned distance metric.

| **Metric** | **eICU-CRD acute kidney injury** | **eICU-CRD 168 h mortality** |
| --- | --- | --- |
| M0 cosine | +0.0197 | +0.0358 |
| M1 mutual information weighted Euclidean | +0.0216 | +0.0367 |
| M2 neighbourhood components analysis,<br>fitted on the development cohort | +0.0105 | +0.0268 |
| M3 neighbourhood components analysis,<br>fitted on the target bank | +0.0131 | +0.0262 |

Change in AUROC against the frozen logistic model at the full bank in eICU-CRD for four retrieval distances: plain cosine (M0, the pre-specified rule), a mutual information weighted Euclidean distance (M1) and two neighbourhood components analyses (M2, M3). M2 was fitted on the development cohort and transferred; M3 was fitted on the target bank itself and is therefore optimistic. Both neighbourhood components variants were worse than cosine, and the mutual information weighted distance was marginally better.

**Supplementary Table S14.**
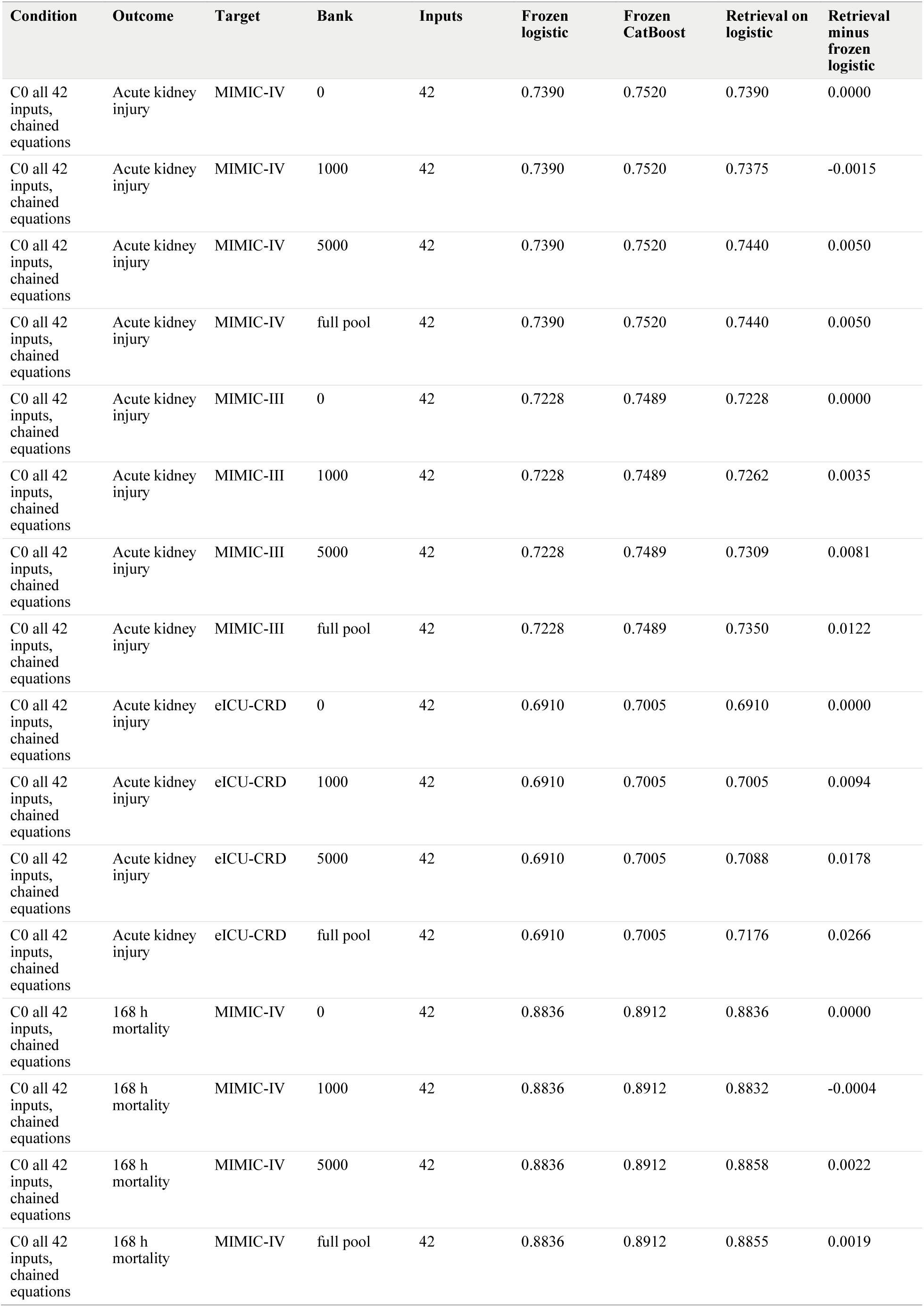
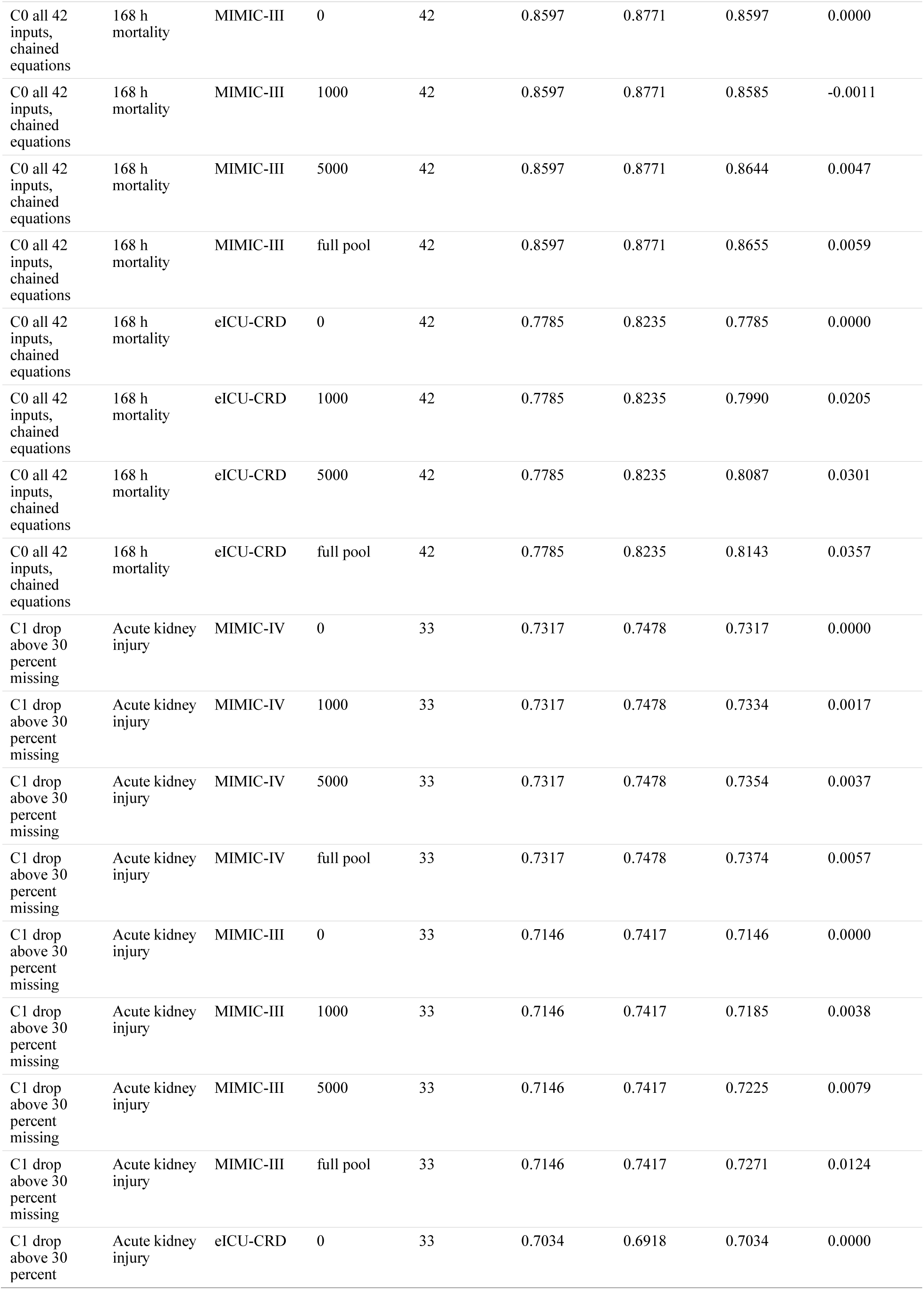
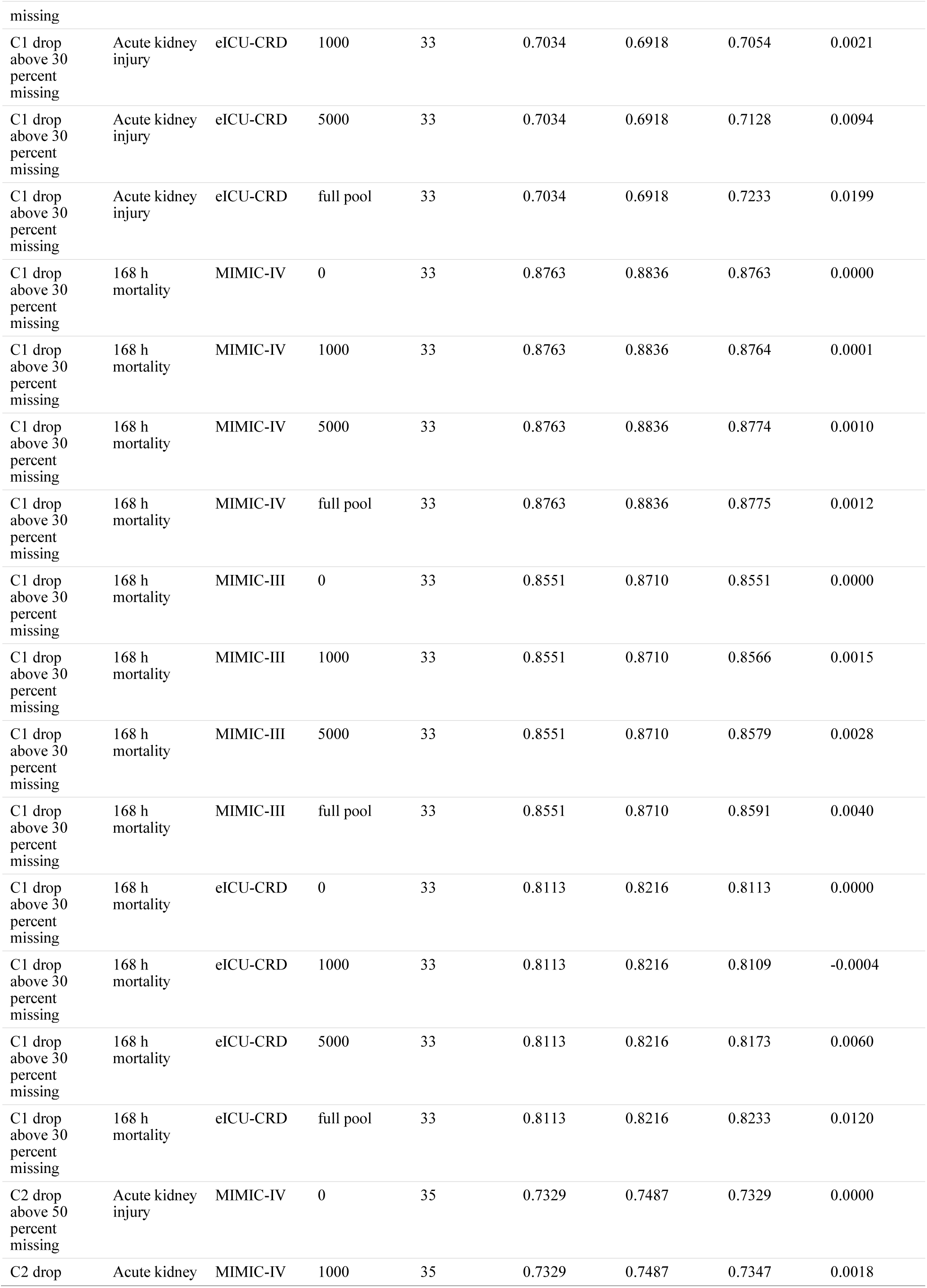
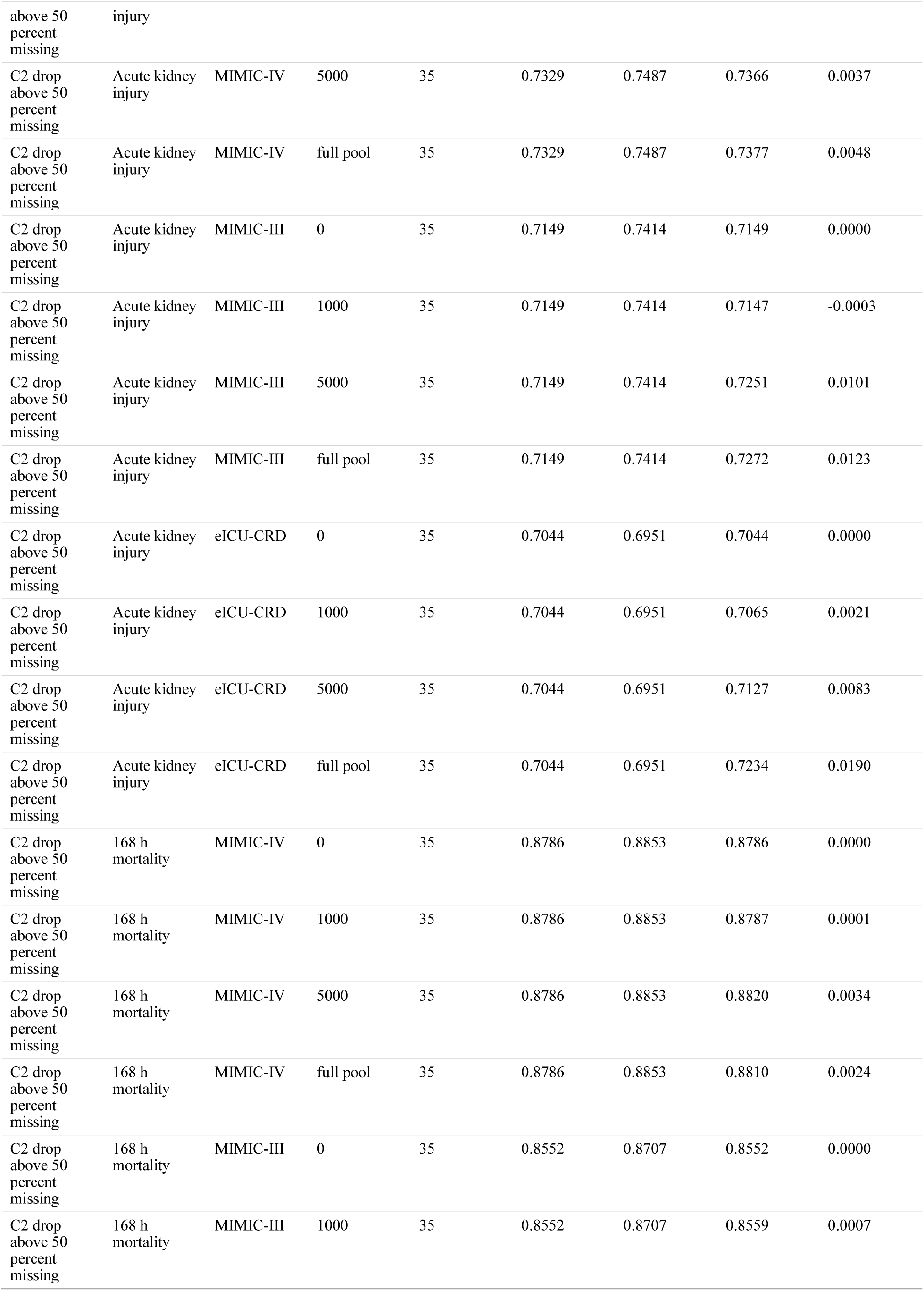
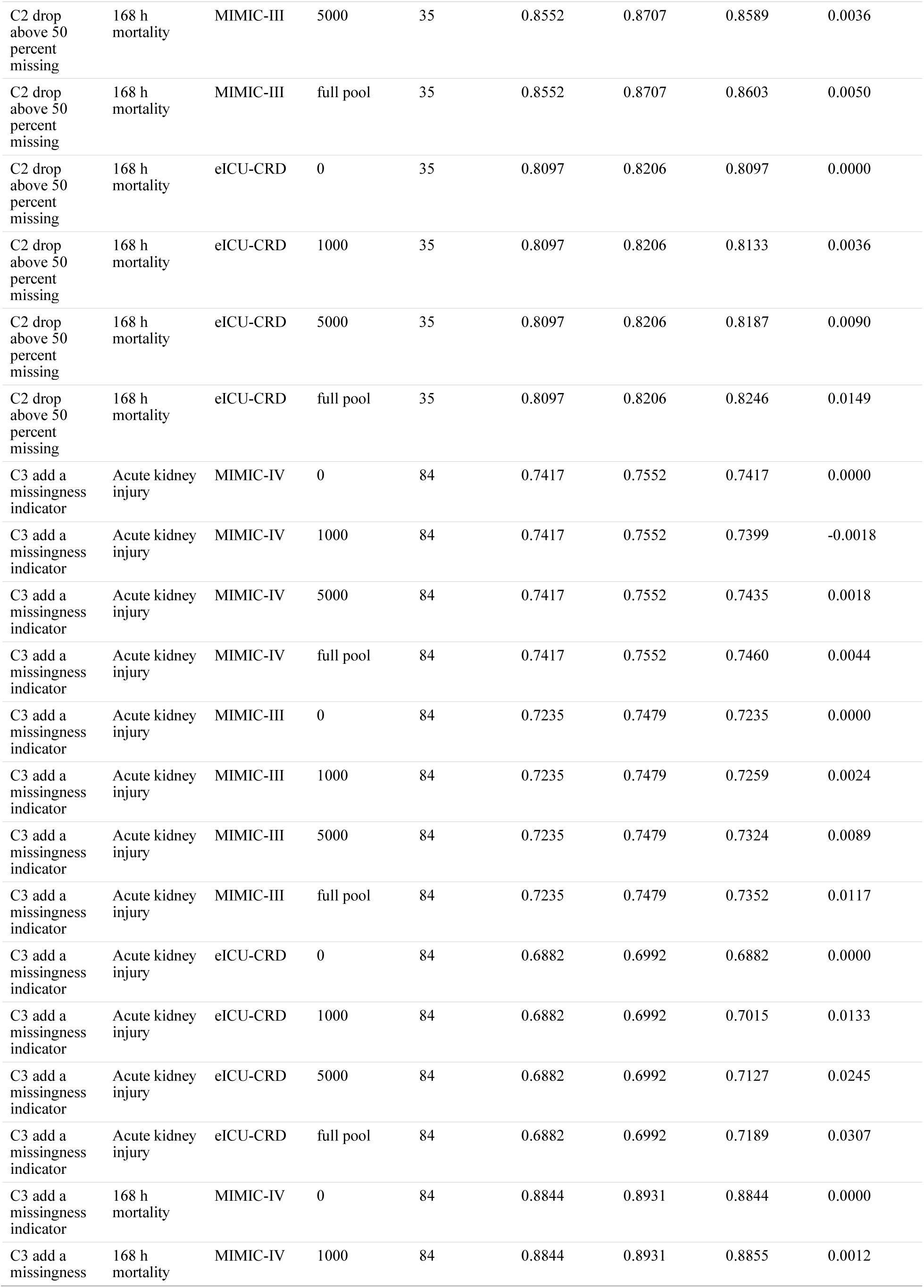
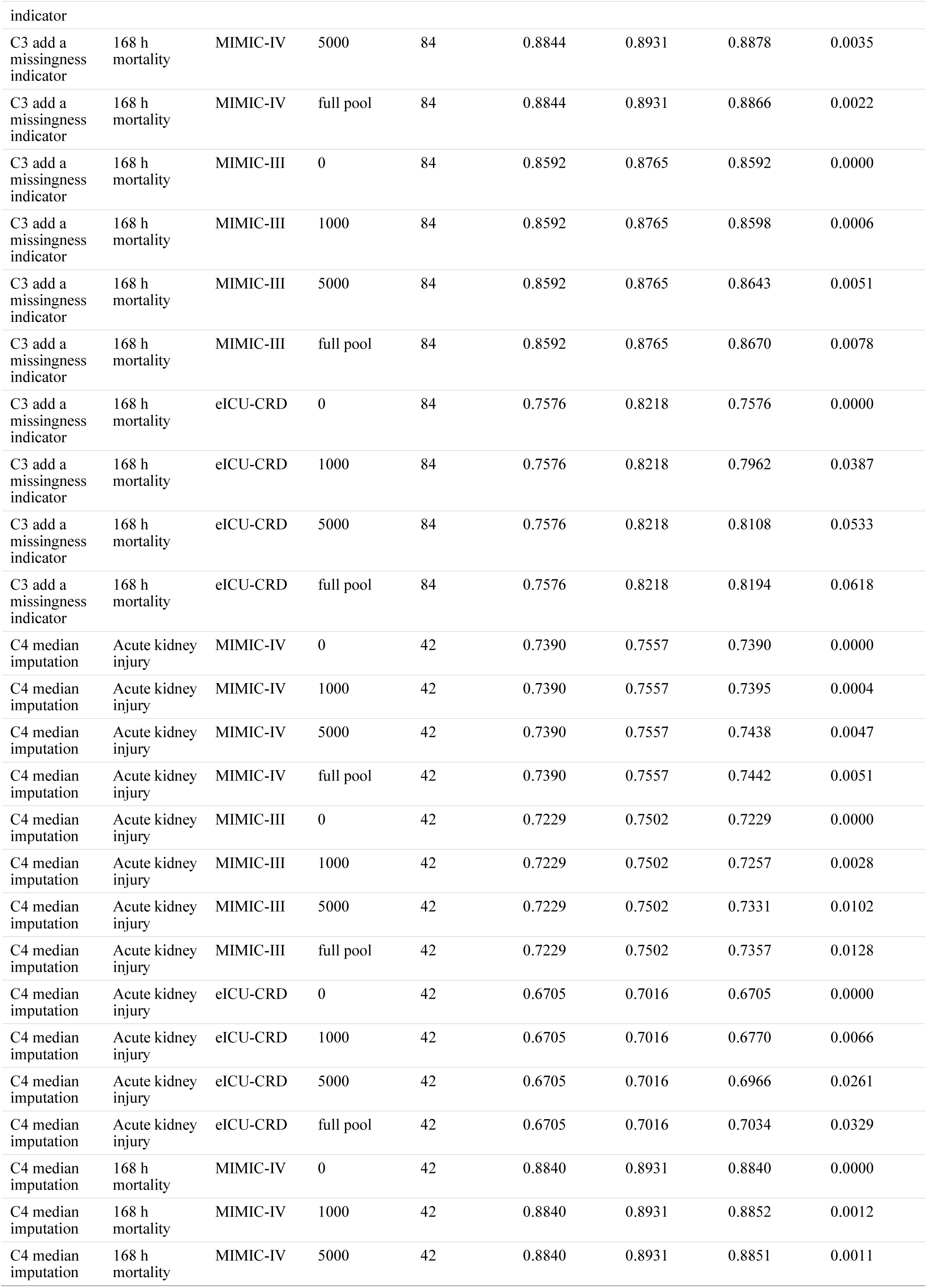
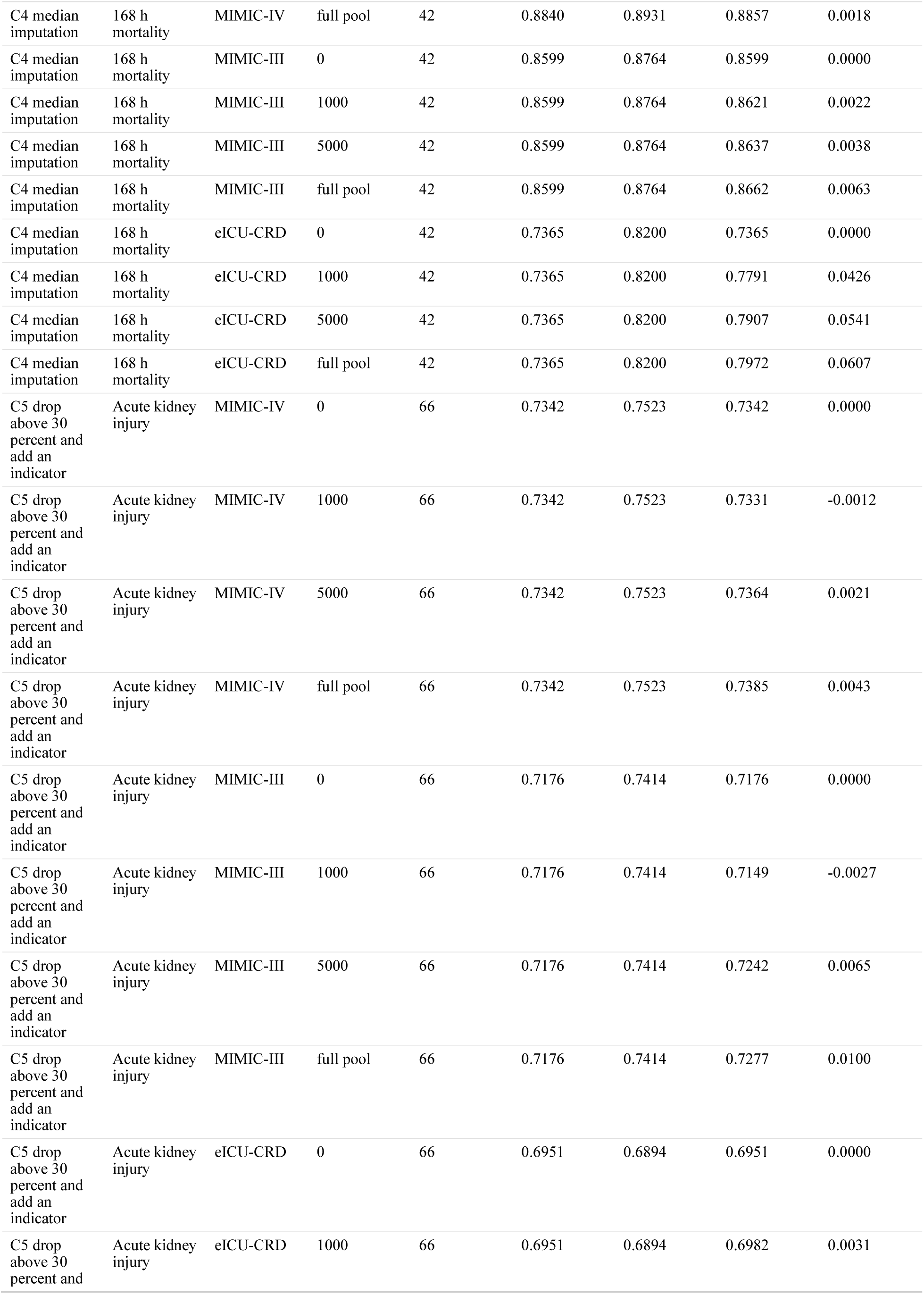
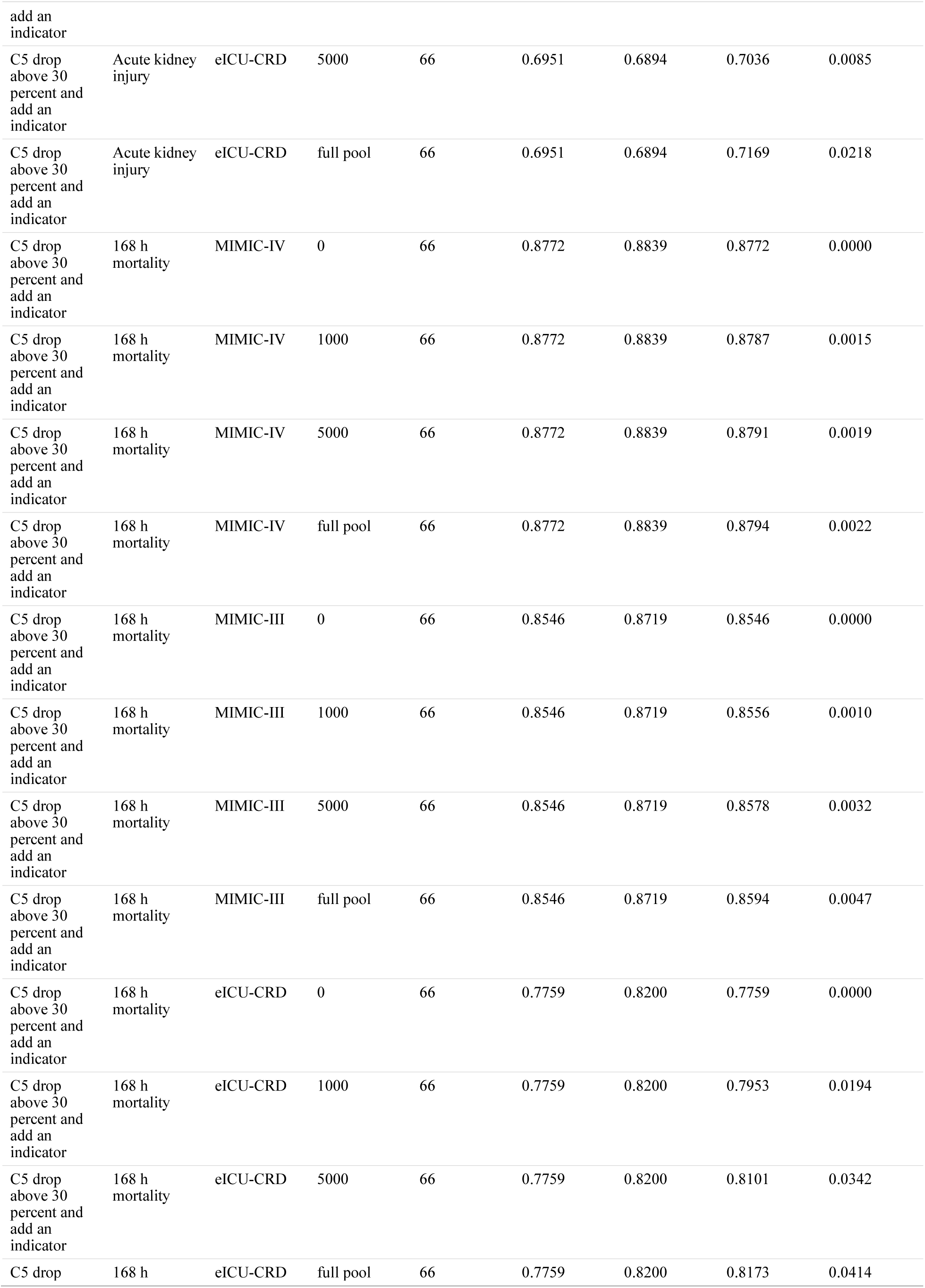
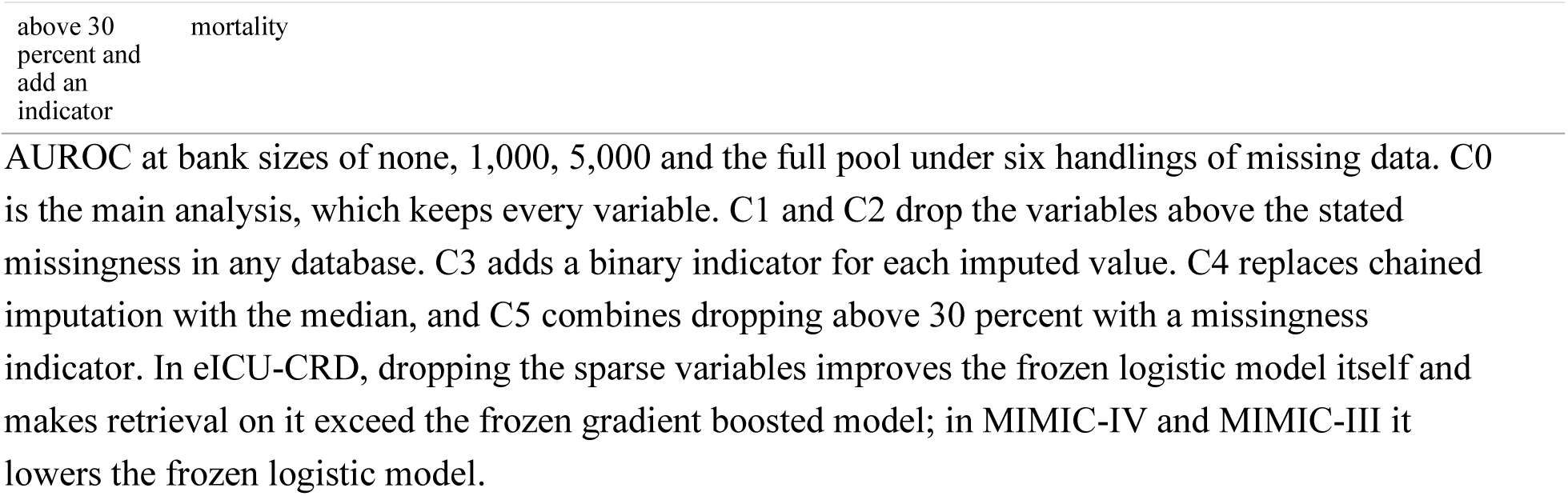
Missing data handling, all conditions.

**Supplementary Table S15.**
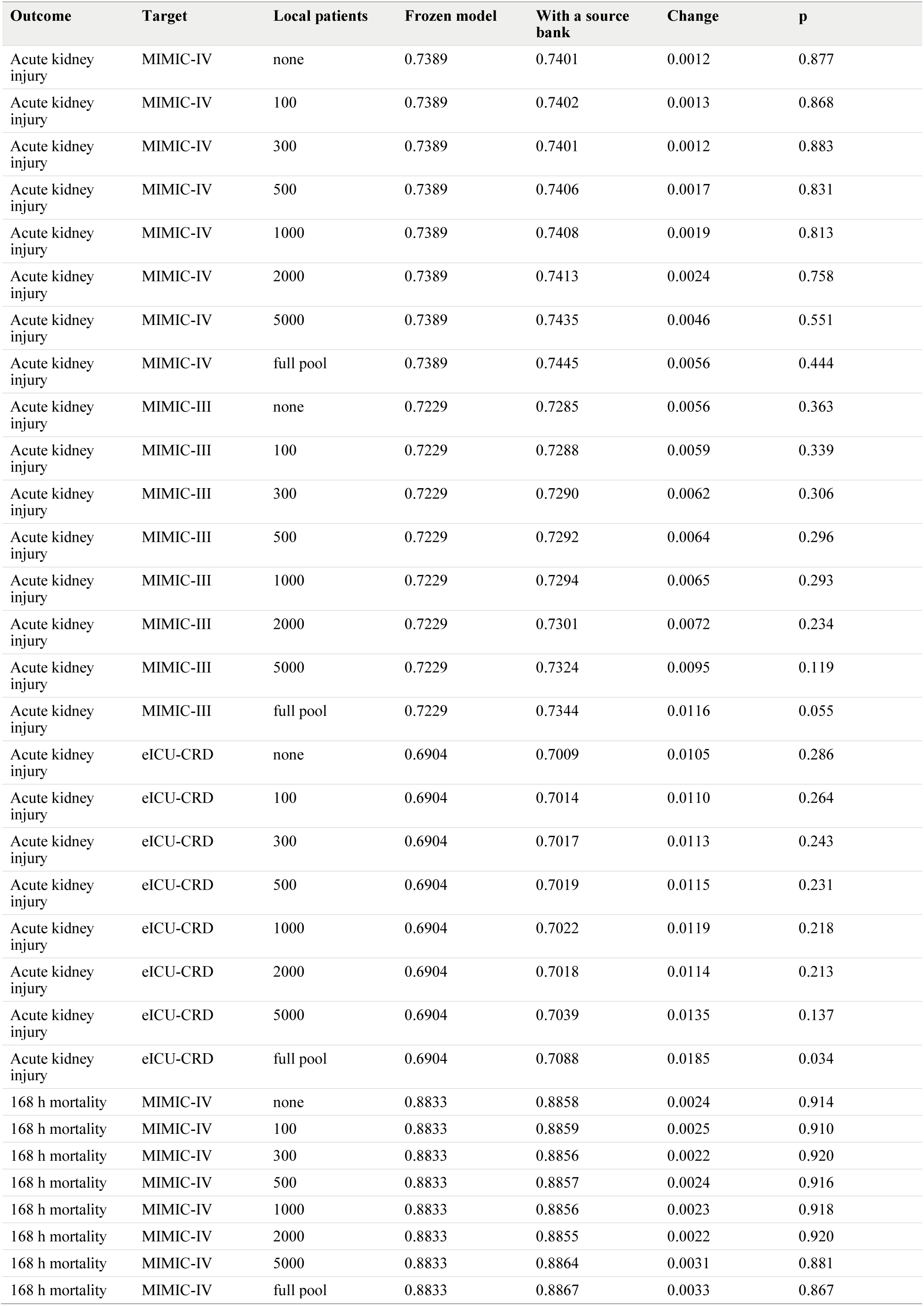
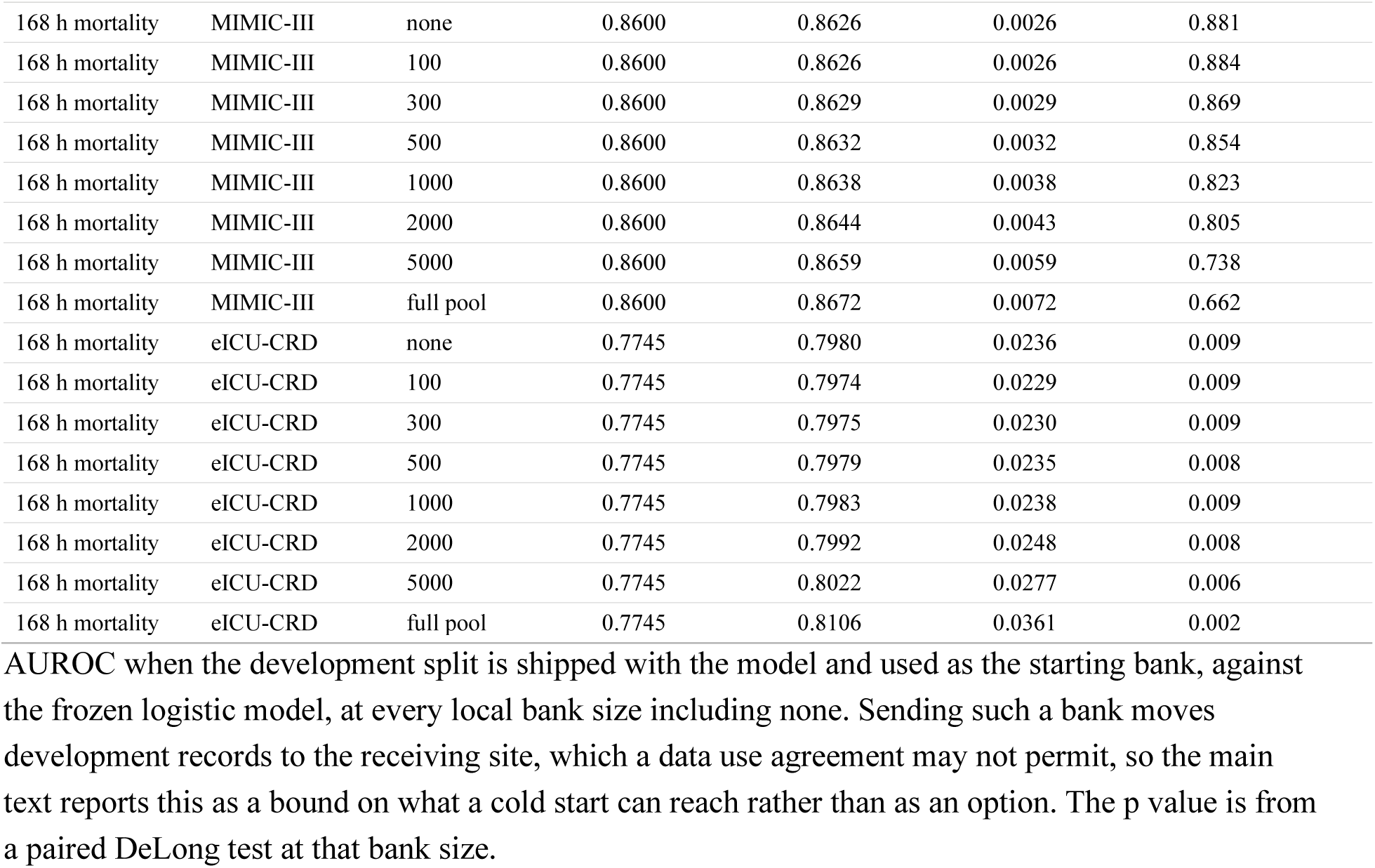
Source bank cold start.

**Supplementary Figure S2.**
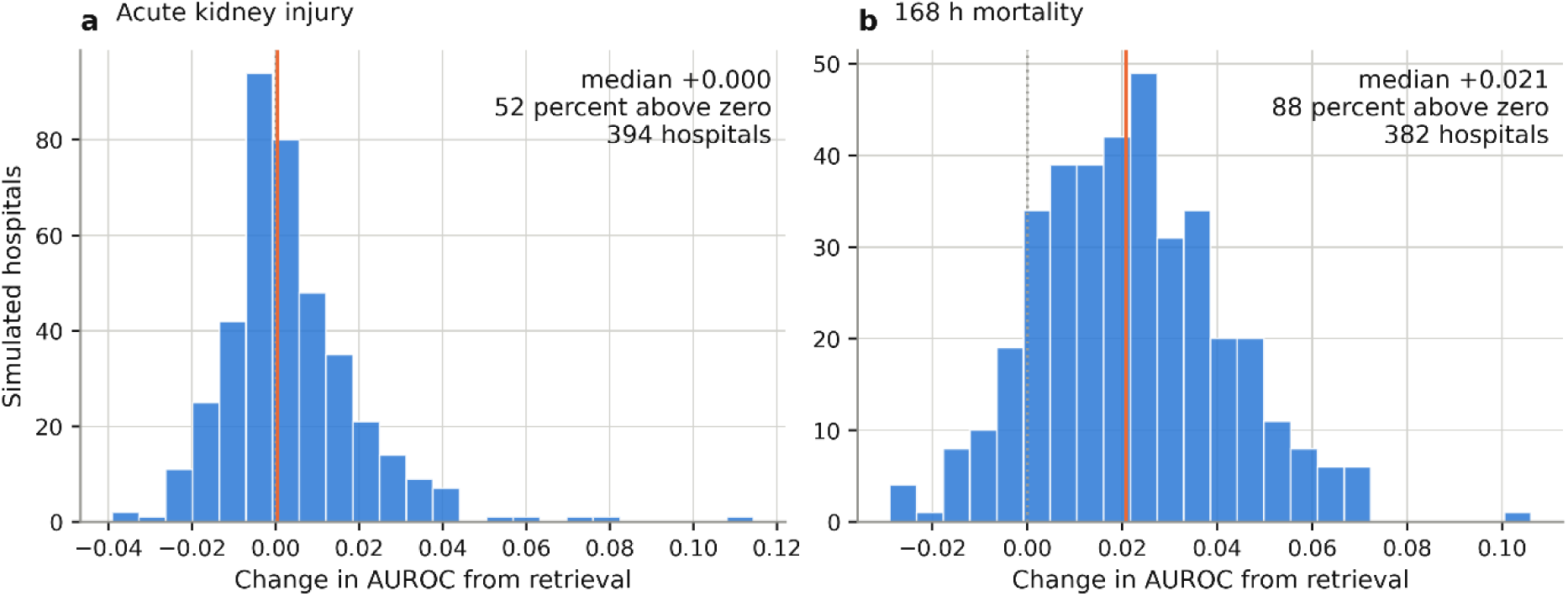
Simulated hospitals. Distribution of the change in AUROC from retrieval across simulated hospitals drawn from eICU-CRD, for acute kidney injury and 168 hour mortality. Two settings of case mix heterogeneity were drawn and are pooled in the figure, which gives 394 simulated hospitals for acute kidney injury and 382 for mortality. Each simulated hospital holds 300 to 3,000 patients whose case mix is drawn from a Dirichlet distribution over ten strata of the feature space, then splits its own patients into a bank and a test set. The simulation varies case mix only. It does not reproduce differences in practice, equipment or recording habits between real hospitals, and is not a substitute for site identifiers. Under the heterogeneous setting retrieval improved 53 percent of 196 hospitals for acute kidney injury and 88 percent of 187 for mortality; under the homogeneous setting it improved 50 percent of 198 and 89 percent of 195. Varying case mix heterogeneity moved neither the proportion improved nor the median.

**Supplementary Table S16.** Hospital-level results, all strategies.

| Outcome | Strategy | Hospitals | Pooled delta AUROC | 95% CI | I<sup>2</sup> | Hospitals improved | Calibration slope | Net benefit at 0.20 |
| --- | --- | --- | --- | --- | --- | --- | --- | --- |
| Acute kidney injury | Frozen logistic | 67 | reference |  |  |  | 0.845 | -0.0014 |
| Acute kidney injury | Frozen CatBoost | 67 | +0.0077 | -0.0042 to +0.0197 | 27% | 49% | 0.843 | -0.0051 |
| Acute kidney injury | Isotonic recalibration | 67 | -0.0267 | -0.0307 to -0.0227 | 0% | 0% | 0.235 | 0.0000 |
| Acute kidney injury | Retrieval on logistic | 67 | +0.0092 | +0.0040 to +0.0144 | 0% | 61% | 1.020 | +0.0012 |
| Acute kidney injury | Retrieval on CatBoost | 67 | +0.0140 | +0.0025 to +0.0256 | 27% | 57% | 1.006 | -0.0021 |
| Acute kidney injury | Local logistic | 67 | -0.0499 | -0.0677 to -0.0320 | 67% | 15% | 0.326 | -0.0037 |
| Acute kidney injury | Pooled logistic | 67 | +0.0068 | +0.0054 to +0.0082 | 0% | 79% | 0.962 | +0.0003 |
| Acute kidney injury | Local CatBoost | 67 | -0.0368 | -0.0532 to -0.0204 | 62% | 25% | 0.439 | -0.0004 |
| 168 h mortality | Frozen logistic | 59 | reference |  |  |  | 0.430 | -0.0036 |
| 168 h mortality | Frozen CatBoost | 59 | +0.0486 | +0.0403 to +0.0568 | 0% | 95% | 0.793 | +0.0029 |
| 168 h mortality | Isotonic recalibration | 59 | -0.0232 | -0.0267 to -0.0196 | 0% | 0% | 0.371 | -0.0005 |
| 168 h mortality | Retrieval on logistic | 59 | +0.0201 | +0.0141 to +0.0262 | 0% | 76% | 0.731 | -0.0018 |
| 168 h mortality | Retrieval on CatBoost | 59 | +0.0484 | +0.0393 to +0.0576 | 0% | 90% | 0.884 | +0.0031 |
| 168 h mortality | Local logistic | 59 | -0.0272 | -0.0463 to -0.0081 | 76% | 36% | 0.428 | +0.0011 |
| 168 h mortality | Pooled logistic | 59 | +0.0302 | +0.0247 to +0.0356 | 0% | 93% | 0.687 | +0.0019 |
| 168 h mortality | Local CatBoost | 59 | +0.0146 | +0.0022 to +0.0270 | 39% | 56% | 0.635 | +0.0011 |

**Supplementary Figure S3.**
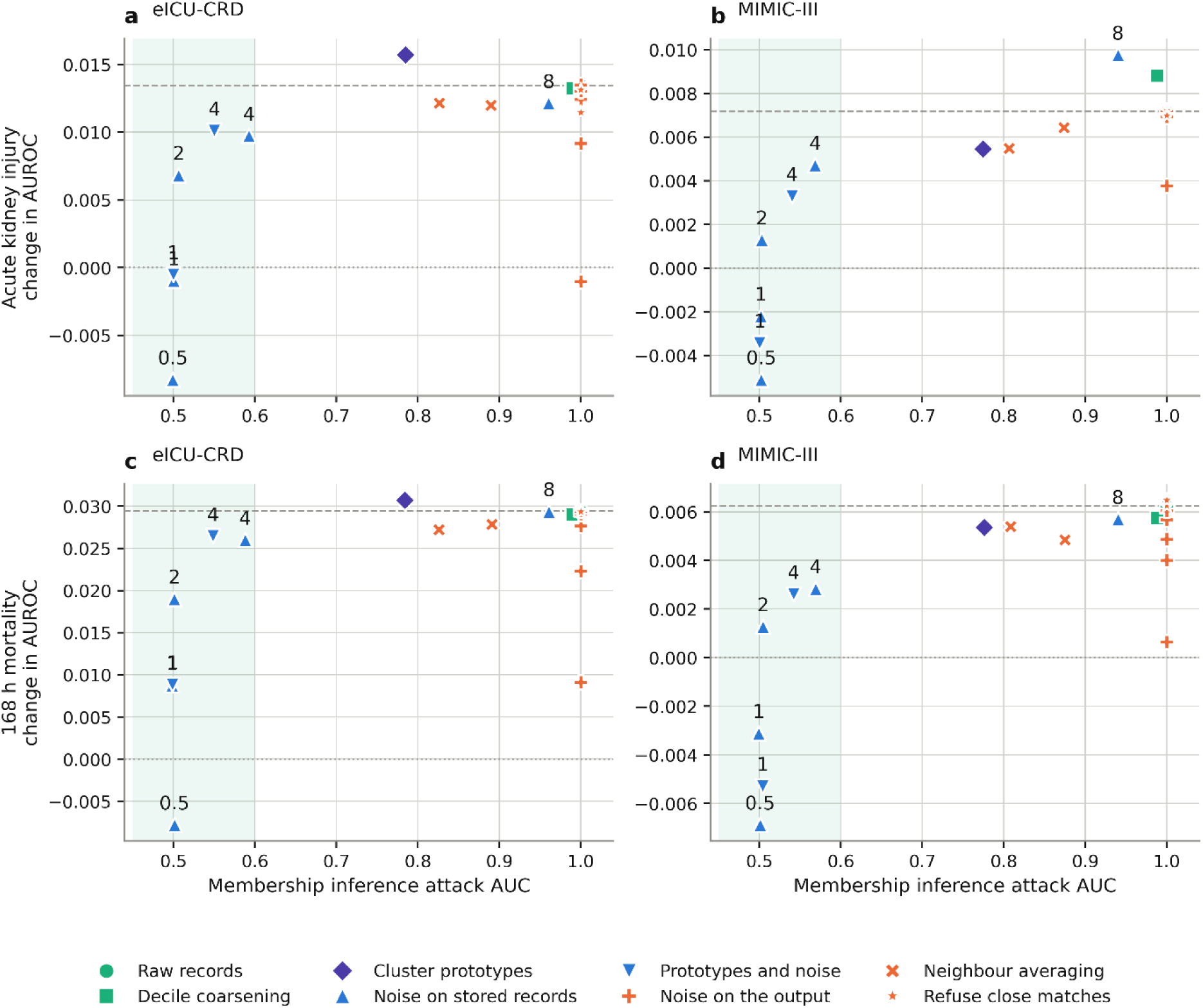
Bank defences, privacy against performance. Each point is one defence applied to the bank. The horizontal axis is the area under the curve of a membership inference attack, where 0.50 is chance and 1.00 is perfect identification of bank members. The vertical axis is the retrieval gain in AUROC against the frozen logistic model under that defence. The shaded band marks attack values between 0.45 and 0.60, which is the region where the attack is close to uninformative. The dashed line is the gain obtained by the raw bank, so a point on that line has given up no performance. Points for differentially private variants are labelled with the privacy budget. Defences that alter the stored records move left; defences applied after retrieval stay at 1.00. Values are in Supplementary Table S17.

**Supplementary Table S17.** Bank defences, representative conditions.

| Outcome | Target | Defence | Parameter | Attack AUC | Delta AUROC |
| --- | --- | --- | --- | --- | --- |
| Acute kidney injury | eICU-CRD | Raw records |  | 1.00 | +0.0135 |
| Acute kidney injury | eICU-CRD | Decile coarsening |  | 0.99 | +0.0133 |
| Acute kidney injury | eICU-CRD | k-anonymity | k = 10 | 0.83 | +0.0122 |
| Acute kidney injury | eICU-CRD | Prototypes | 1,000 | 0.78 | +0.0157 |
| Acute kidney injury | eICU-CRD | Noise on records | epsilon = 4 | 0.59 | +0.0097 |
| Acute kidney injury | eICU-CRD | Prototypes with noise | epsilon = 4 | 0.55 | +0.0101 |
| Acute kidney injury | eICU-CRD | Noise on the output | epsilon = 4 | 1.00 | +0.0130 |
| Acute kidney injury | eICU-CRD | Refusal | closest 5% | 1.00 | +0.0115 |
| 168 h mortality | eICU-CRD | Raw records |  | 1.00 | +0.0294 |
| 168 h mortality | eICU-CRD | Decile coarsening |  | 0.99 | +0.0290 |
| 168 h mortality | eICU-CRD | k-anonymity | k = 10 | 0.83 | +0.0272 |
| 168 h mortality | eICU-CRD | Prototypes | 1,000 | 0.78 | +0.0307 |
| 168 h mortality | eICU-CRD | Noise on records | epsilon = 4 | 0.59 | +0.0260 |
| 168 h mortality | eICU-CRD | Prototypes with noise | epsilon = 4 | 0.55 | +0.0264 |
| 168 h mortality | eICU-CRD | Noise on the output | epsilon = 4 | 1.00 | +0.0291 |
| 168 h mortality | eICU-CRD | Refusal | closest 5% | 1.00 | +0.0293 |
| Acute kidney injury | MIMIC-III | Raw records |  | 1.00 | +0.0072 |
| Acute kidney injury | MIMIC-III | Noise on records | epsilon = 4 | 0.57 | +0.0047 |
| Acute kidney injury | MIMIC-III | Prototypes with noise | epsilon = 4 | 0.54 | +0.0033 |
| 168 h mortality | MIMIC-III | Raw records |  | 1.00 | +0.0062 |
| 168 h mortality | MIMIC-III | Noise on records | epsilon = 4 | 0.57 | +0.0028 |
| 168 h mortality | MIMIC-III | Prototypes with noise | epsilon = 4 | 0.54 | +0.0026 |

**Supplementary Figure S4.**
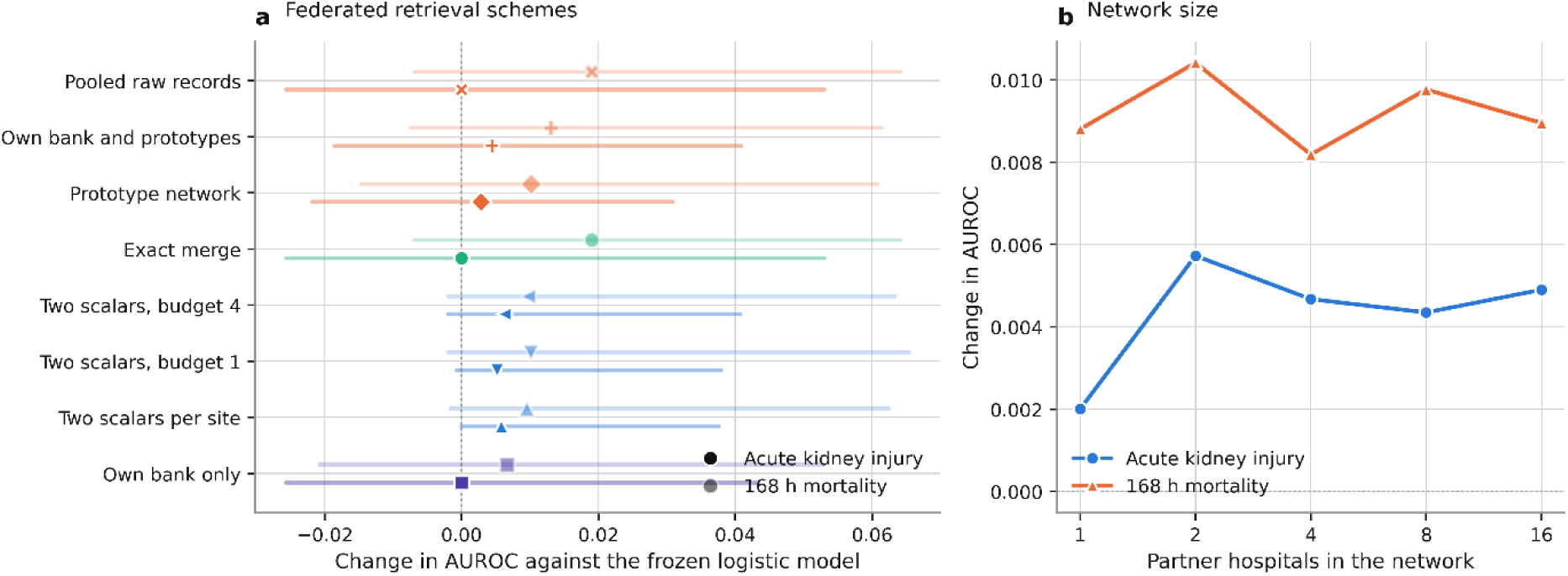
Federated retrieval schemes and network size. a, Change in AUROC against the frozen logistic model for each federated scheme, across the same eICU-CRD hospitals as Figure 3. The exact merge and the pooled raw bank lie on top of one another, which is the empirical form of the identity described in the Results. b, Change in AUROC against the hospital’s own bank as the number of partner hospitals increases from 1 to 16. Most of the available gain arrives with the first one or two partners. Values are in Supplementary Table S18.

**Supplementary Table S18.**
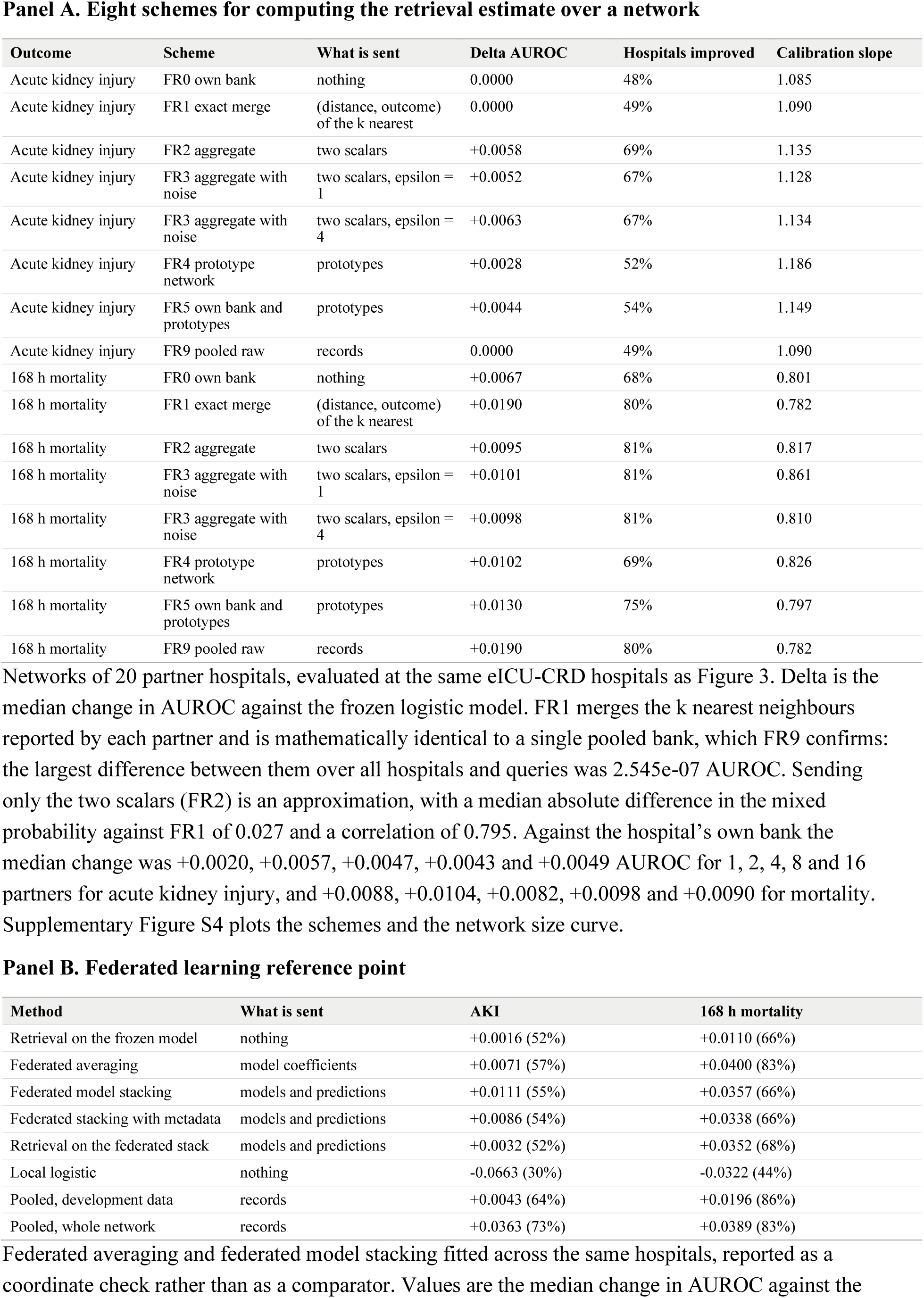
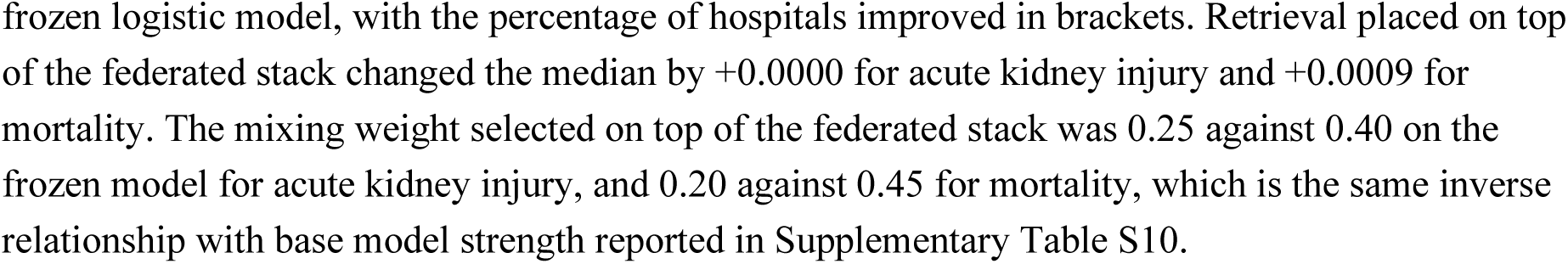
Federated retrieval schemes, all conditions.

Panel A. Eight schemes for computing the retrieval estimate over a network
| Outcome | Scheme | What is sent | Delta AUROC | Hospitals improved | Calibration slope |
| --- | --- | --- | --- | --- | --- |
| Acute kidney injury | FR0 own bank | nothing | 0.0000 | 48% | 1.085 |
| Acute kidney injury | FR1 exact merge | (distance, outcome) of the k nearest | 0.0000 | 49% | 1.090 |
| Acute kidney injury | FR2 aggregate | two scalars | +0.0058 | 69% | 1.135 |
| Acute kidney injury | FR3 aggregate with noise | two scalars, epsilon = 1 | +0.0052 | 67% | 1.128 |
| Acute kidney injury | FR3 aggregate with noise | two scalars, epsilon = 4 | +0.0063 | 67% | 1.134 |
| Acute kidney injury | FR4 prototype network | prototypes | +0.0028 | 52% | 1.186 |
| Acute kidney injury | FR5 own bank and prototypes | prototypes | +0.0044 | 54% | 1.149 |
| Acute kidney injury | FR9 pooled raw | records | 0.0000 | 49% | 1.090 |
| 168 h mortality | FR0 own bank | nothing | +0.0067 | 68% | 0.801 |
| 168 h mortality | FR1 exact merge | (distance, outcome) of the k nearest | +0.0190 | 80% | 0.782 |
| 168 h mortality | FR2 aggregate | two scalars | +0.0095 | 81% | 0.817 |
| 168 h mortality | FR3 aggregate with noise | two scalars, epsilon = 1 | +0.0101 | 81% | 0.861 |
| 168 h mortality | FR3 aggregate with noise | two scalars, epsilon = 4 | +0.0098 | 81% | 0.810 |
| 168 h mortality | FR4 prototype network | prototypes | +0.0102 | 69% | 0.826 |
| 168 h mortality | FR5 own bank and prototypes | prototypes | +0.0130 | 75% | 0.797 |
| 168 h mortality | FR9 pooled raw | records | +0.0190 | 80% | 0.782 |

Panel B. Federated learning reference point
| Method | What is sent | AKI | 168 h mortality |
| --- | --- | --- | --- |
| Retrieval on the frozen model | nothing | +0.0016 (52%) | +0.0110 (66%) |
| Federated averaging | model coefficients | +0.0071 (57%) | +0.0400 (83%) |
| Federated model stacking | models and predictions | +0.0111 (55%) | +0.0357 (66%) |
| Federated stacking with metadata | models and predictions | +0.0086 (54%) | +0.0338 (66%) |
| Retrieval on the federated stack | models and predictions | +0.0032 (52%) | +0.0352 (68%) |
| Local logistic | nothing | -0.0663 (30%) | -0.0322 (44%) |
| Pooled, development data | records | +0.0043 (64%) | +0.0196 (86%) |
| Pooled, whole network | records | +0.0363 (73%) | +0.0389 (83%) |

Federated averaging and federated model stacking fitted across the same hospitals, reported as a coordinate check rather than as a comparator. Values are the median change in AUROC against the

**Supplementary Table S19.** Retrieval network, combination rules and local bank sizes.

Panel A. Combination rules at the full local bank
| Outcome | Rule | Delta AUROC | Hospitals improved | Calibration slope | ECE | Baseline slope | Baseline ECE |
| --- | --- | --- | --- | --- | --- | --- | --- |
| Acute kidney injury | Own bank only | +0.0080 | 66% | 1.075 | 0.0340 | 0.861 | 0.0439 |
| Acute kidney injury | Mean | +0.0155 | 80% | 1.144 | 0.0347 | 0.861 | 0.0439 |
| Acute kidney injury | Size weighted | +0.0142 | 80% | 1.134 | 0.0349 | 0.861 | 0.0439 |
| Acute kidney injury | Similarity weighted | +0.0157 | 80% | 1.141 | 0.0344 | 0.861 | 0.0439 |
| Acute kidney injury | Precision weighted | +0.0166 | 79% | 1.086 | 0.0302 | 0.861 | 0.0439 |
| Acute kidney injury | Median | +0.0152 | 81% | 1.136 | 0.0345 | 0.861 | 0.0439 |
| Acute kidney injury | Trimmed mean | +0.0154 | 81% | 1.144 | 0.0346 | 0.861 | 0.0439 |
| Acute kidney injury | Mean in logit space | +0.0162 | 82% | 1.111 | 0.0325 | 0.861 | 0.0439 |
| Acute kidney injury | Local stack | +0.0118 | 68% | 1.093 | 0.0354 | 0.861 | 0.0439 |
| Acute kidney injury | Best single partner | +0.0133 | 72% | 1.084 | 0.0350 | 0.861 | 0.0439 |
| 168 h mortality | Own bank only | +0.0202 | 74% | 0.880 | 0.0341 | 0.481 | 0.0532 |
| 168 h mortality | Mean | +0.0234 | 85% | 0.891 | 0.0370 | 0.481 | 0.0532 |
| 168 h mortality | Size weighted | +0.0230 | 87% | 0.889 | 0.0370 | 0.481 | 0.0532 |
| 168 h mortality | Similarity weighted | +0.0242 | 86% | 0.891 | 0.0370 | 0.481 | 0.0532 |
| 168 h mortality | Precision weighted | +0.0218 | 80% | 0.824 | 0.0338 | 0.481 | 0.0532 |
| 168 h mortality | Median | +0.0230 | 85% | 0.881 | 0.0363 | 0.481 | 0.0532 |
| 168 h mortality | Trimmed mean | +0.0221 | 86% | 0.884 | 0.0366 | 0.481 | 0.0532 |
| 168 h mortality | Mean in logit space | +0.0230 | 85% | 0.846 | 0.0341 | 0.481 | 0.0532 |
| 168 h mortality | Local stack | +0.0221 | 78% | 0.814 | 0.0352 | 0.481 | 0.0532 |
| 168 h mortality | Best single partner | +0.0172 | 73% | 0.852 | 0.0373 | 0.481 | 0.0532 |

Panel B. Local bank size, mean rule against own bank only
| Outcome | Local patients | Network delta | Network improved | Network slope | Own bank delta | Own bank improved |
| --- | --- | --- | --- | --- | --- | --- |
| Acute kidney injury | 0 | +0.0168 | 84% | 1.058 | not available |  |
| Acute kidney injury | 100 | +0.0108 | 76% | 1.101 | +0.0035 | 61% |
| Acute kidney injury | 300 | +0.0173 | 84% | 1.177 | +0.0111 | 72% |
| Acute kidney injury | 1,000 | +0.0197 | 78% | 1.177 | +0.0080 | 66% |
| Acute kidney injury | full pool | +0.0198 | 78% | 1.177 | +0.0082 | 67% |
| 168 h mortality | 0 | +0.0174 | 88% | 0.757 | not available |  |
| 168 h mortality | 100 | +0.0207 | 86% | 0.926 | +0.0112 | 66% |
| 168 h mortality | 300 | +0.0249 | 85% | 0.922 | +0.0249 | 78% |
| 168 h mortality | 1,000 | +0.0253 | 83% | 0.928 | +0.0203 | 76% |
| 168 h mortality | full pool | +0.0255 | 83% | 0.928 | +0.0203 | 76% |

Panel C. Robustness to one partner with permuted outcome labels
| Outcome | Combination rule | All partners clean | One partner corrupted | AUROC lost |
| --- | --- | --- | --- | --- |
| Acute kidney injury | Median | 0.7113 | 0.7121 | -0.0009 |
| Acute kidney injury | Mean in logit space | 0.7136 | 0.7144 | -0.0008 |
| Acute kidney injury | Trimmed mean | 0.7108 | 0.7110 | -0.0002 |
| Acute kidney injury | Size weighted | 0.7129 | 0.7116 | +0.0014 |
| Acute kidney injury | Similarity weighted | 0.7128 | 0.7114 | +0.0015 |
| Acute kidney injury | Mean | 0.7135 | 0.7097 | +0.0038 |
| Acute kidney injury | Precision weighted | 0.7159 | 0.7095 | +0.0064 |
| Acute kidney injury | Best single partner | 0.7161 | 0.7061 | +0.0100 |
| Acute kidney injury | Local stack | 0.7183 | 0.6932 | +0.0250 |
| Acute kidney injury | Own bank only | 0.7081 | not applicable | not applicable |
| 168 h mortality | Median | 0.8198 | 0.8171 | +0.0027 |
| 168 h mortality | Mean in logit space | 0.8223 | 0.8229 | -0.0006 |
| 168 h mortality | Trimmed mean | 0.8204 | 0.8192 | +0.0011 |
| 168 h mortality | Size weighted | 0.8164 | 0.8149 | +0.0015 |
| 168 h mortality | Similarity weighted | 0.8192 | 0.8144 | +0.0048 |
| 168 h mortality | Mean | 0.8194 | 0.8167 | +0.0027 |
| 168 h mortality | Precision weighted | 0.8154 | 0.8063 | +0.0091 |
| 168 h mortality | Best single partner | 0.8135 | 0.8119 | +0.0016 |
| 168 h mortality | Local stack | 0.8155 | 0.8117 | +0.0038 |
| 168 h mortality | Own bank only | 0.8101 | not applicable | not applicable |

At the full local bank, one partner returns probabilities computed from permuted outcomes and the querying hospital combines the answers as before. A positive value in the last column is AUROC lost, and a negative value means the rule was not harmed. For acute kidney injury the loss grows from the rules that summarize the partner answers to the rules that weight or select among them, and reaches 0.025 for the locally fitted stack. Losses were smaller for mortality, where no rule lost more than 0.010. Own bank only is listed for reference and has no partner to corrupt.

**Supplementary Table S20.** Label-free prediction smoothing.

| Target | Outcome | Bank | Label-free smoothing | Retrieval with labels |
| --- | --- | --- | --- | --- |
| eICU-CRD | Acute kidney injury | 1,000 | +0.0107 | +0.0045 |
| eICU-CRD | Acute kidney injury | 5,000 | +0.0103 | +0.0124 |
| eICU-CRD | 168 h mortality | 1,000 | +0.0207 | +0.0174 |
| eICU-CRD | 168 h mortality | 5,000 | +0.0213 | +0.0264 |
| MIMIC-IV | 168 h mortality | Full | -0.0058 | +0.0042 |

Change in AUROC against the frozen logistic model in the bank simulation, for a variant that averages the frozen model output over the retrieved neighbours instead of their outcomes, so that no local outcome label is needed, compared with retrieval using labels. The variant can be switched on the day the model is installed, before any local outcome has been observed. Where the shift is large the label-free variant recovers most of the gain, and at a bank of 1,000 it is ahead. Where the shift is small it is a small loss.

**Supplementary Figure S5.**
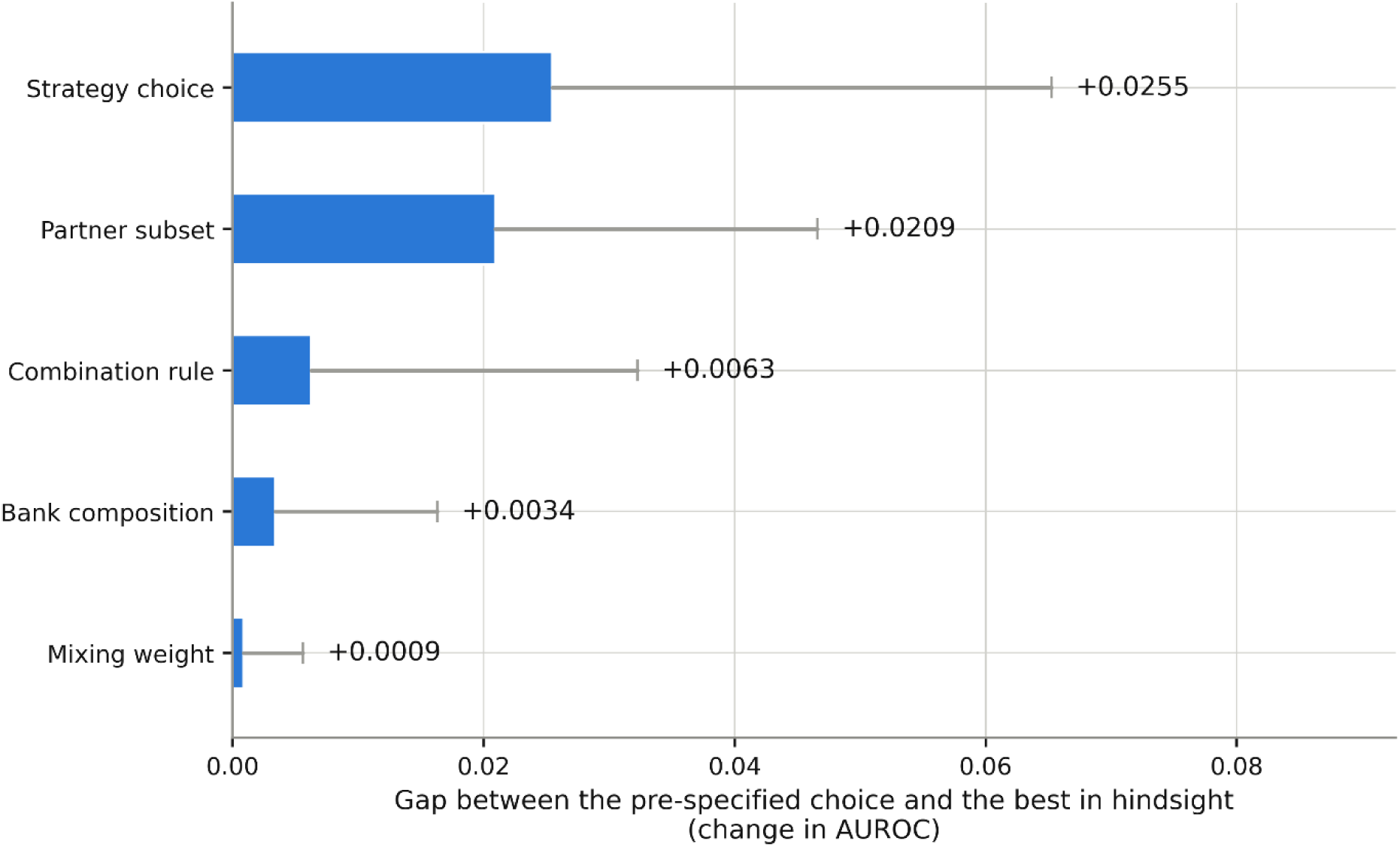
Gap to the best choice in hindsight. The gap between each pre-specified choice and the choice that would have been best in hindsight, in AUROC. The bar is the median and the thin line extends to the 90th percentile; the annotated value is the median. None of these can be obtained at deployment, so the bars are a measurement of remaining room rather than of achievable performance. The two largest gaps, strategy selection and partner selection, both concern which information to use; the smallest, the mixing weight, concerns how to weight it. Values are in Supplementary Table S21.

**Supplementary Table S21.** Oracle bounds, all five choices.

| Choice | Pre-specified as | Median gap in AUROC | Notes |
| --- | --- | --- | --- |
| Strategy, per hospital | Retrieval on the frozen logistic model | +0.0255 | Gap against the frozen model is +0.0216 for acute kidney injury and +0.0508 for mortality |
| Partner subset | All 20 partners | +0.0209 | The hindsight-best network held a median of 2 partners for acute kidney injury and 1 for mortality |
| Combination rule | Mean of the partner probabilities | +0.0063 | Best rule in hindsight varies by hospital; no rule was best at more than a quarter of them |
| Bank composition rule | Random sampling | +0.0034 | Prevalence balancing was the hindsight winner in 10 of 12 settings |
| Mixing weight | Three-fold cross-validation inside the bank | +0.0009 | The cross-validated weight cost less than 0.002 AUROC against the oracle weight in 17 of 18 settings; the exception was eICU-CRD mortality at the full bank |

**Supplementary Figure S6.**
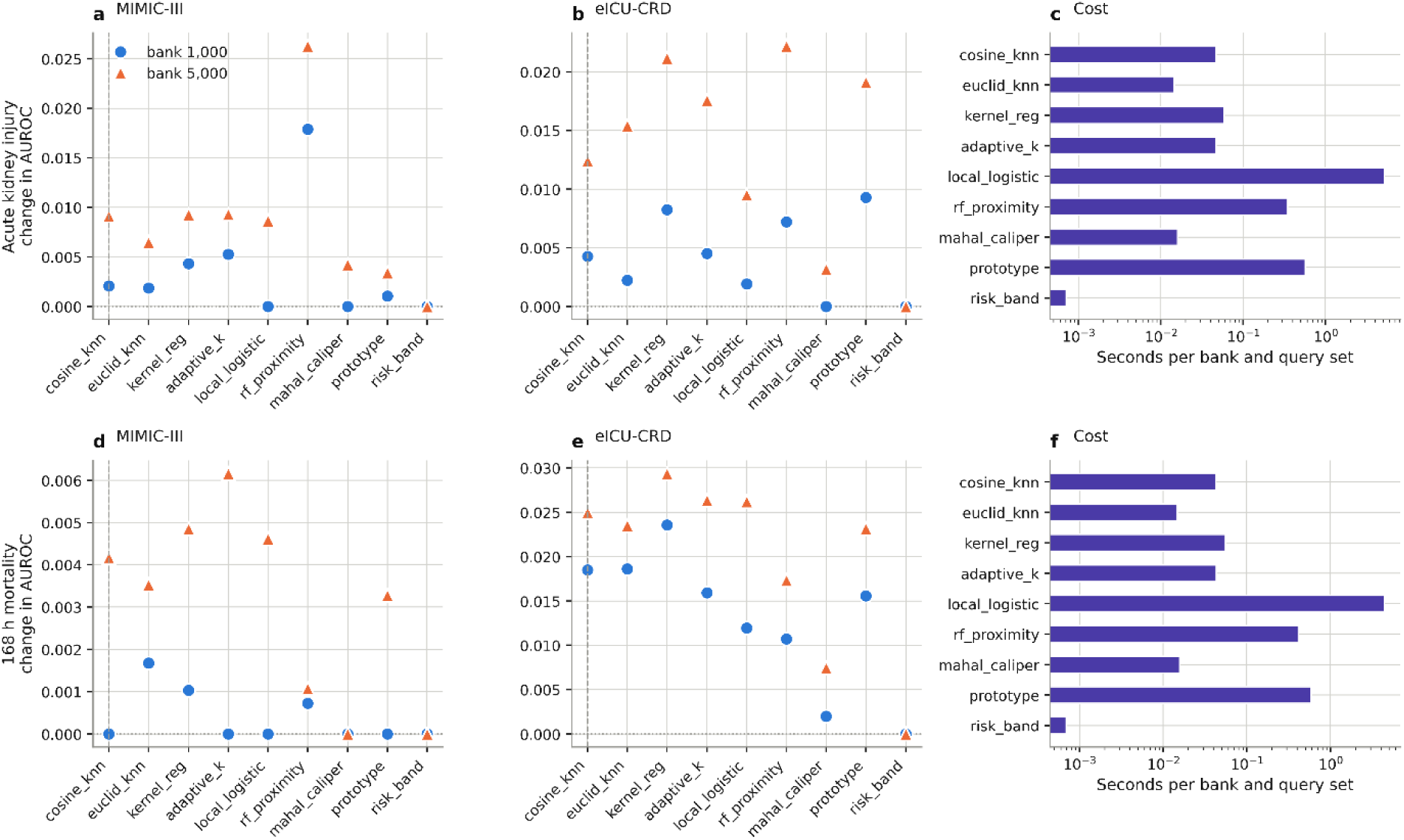
Reference class estimators and their cost. a, b, d, e, Change in AUROC against the frozen base model for nine ways of deciding which bank patients count as similar to the query, in each cohort, outcome and bank size. The pre-specified cosine rule is marked by the dashed line. c, f, Seconds to answer 2,000 queries, on a logarithmic axis. No estimator was best everywhere, and the two most expensive were not the two best. Values are in Supplementary Table S22.

**Supplementary Table S22.** Reference class estimators, all conditions.

| Estimator | AKI<br>MIMIC-<br>III 1k | AKI<br>MIMIC-<br>III 5k | AKI eICU<br>1k | AKI eICU<br>5k | Mortality<br>MIMIC-III<br>1k | Mortality<br>MIMIC-<br>III 5k | Mortality<br>eICU 1k | Mortality<br>eICU 5k | Seconds<br>per 2,000<br>queries |
| --- | --- | --- | --- | --- | --- | --- | --- | --- | --- |
| S0 cosine<br>kNN (pre-specified) | +0.0021 | +0.0091 | +0.0043 | +0.0124 | 0.0000 | +0.0042 | +0.0185 | +0.0250 | 0.044 |
| S1<br>Euclidean<br>kNN | +0.0019 | +0.0065 | +0.0022 | +0.0154 | +0.0017 | +0.0035 | +0.0186 | +0.0235 | 0.015 |
| S2 kernel<br>regression | +0.0043 | +0.0093 | +0.0083 | +0.0212 | +0.0010 | +0.0049 | +0.0236 | +0.0294 | 0.060 |
| S3 adaptive<br>k | +0.0053 | +0.0093 | +0.0045 | +0.0176 | 0.0000 | +0.0062 | +0.0159 | +0.0264 | 0.045 |
| S4 local<br>logistic | 0.0000 | +0.0086 | +0.0019 | +0.0095 | 0.0000 | +0.0046 | +0.0120 | +0.0262 | 4.859 |
| S5 random<br>forest<br>proximity | +0.0179 | +0.0263 | +0.0072 | +0.0222 | +0.0007 | +0.0011 | +0.0107 | +0.0173 | 0.353 |
| S6<br>Mahalanobis<br>caliper | 0.0000 | +0.0042 | 0.0000 | +0.0032 | 0.0000 | 0.0000 | +0.0020 | +0.0074 | 0.016 |
| S7 learned<br>prototypes | +0.0010 | +0.0034 | +0.0093 | +0.0191 | 0.0000 | +0.0033 | +0.0156 | +0.0232 | 0.584 |
| S8 predicted<br>risk band | 0.0000 | 0.0000 | 0.0000 | 0.0000 | 0.0000 | 0.0000 | 0.0000 | 0.0000 | 0.001 |

**Supplementary Figure S7.**
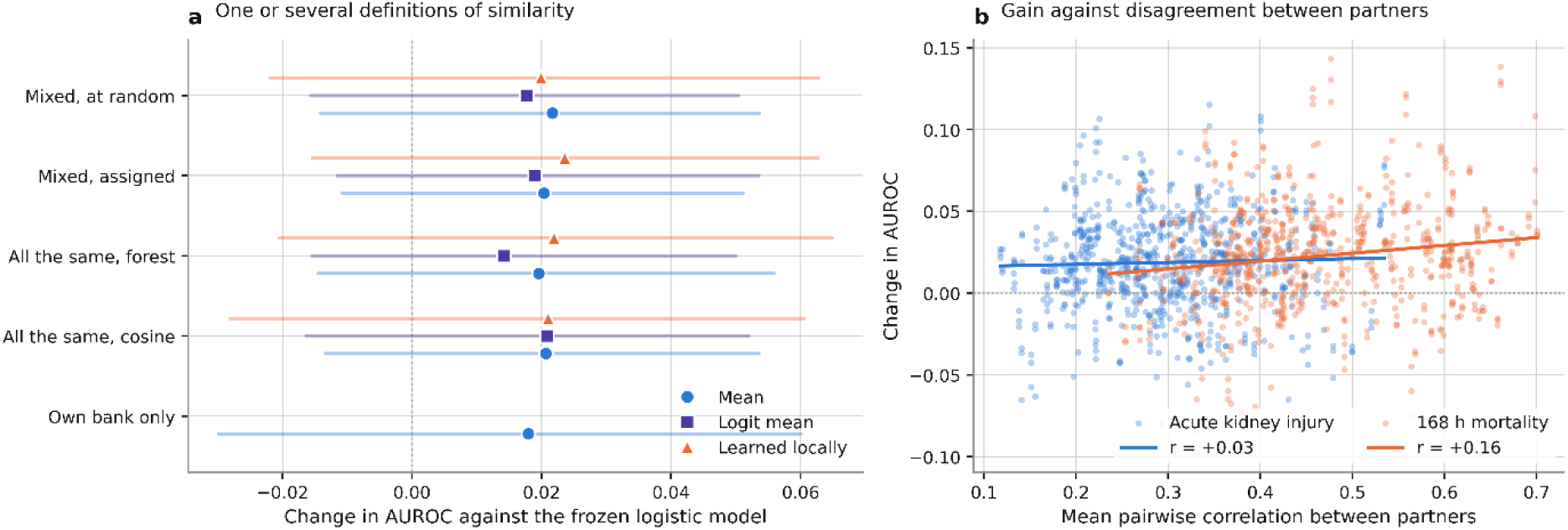
Networks with one or several definitions of similarity. a, Change in AUROC against the frozen logistic model for homogeneous and heterogeneous networks under three combination rules. b, For every querying hospital, the gain plotted against the mean pairwise correlation between the probabilities its partners returned, with the correlation coefficient annotated. If disagreement between partners carried extra information, the points would slope downward. They do not. Values are in Supplementary Table S23.

**Supplementary Table S23.** Heterogeneous similarity networks.

| Outcome | Network | Combiner | Definitions | Pairwise correlation | Delta AUROC | Hospitals improved |
| --- | --- | --- | --- | --- | --- | --- |
| Acute kidney injury | Homogeneous, cosine | Mean | 1 | 0.365 | +0.0185 | 79% |
| Acute kidney injury | Homogeneous, forest proximity | Mean | 1 | 0.324 | +0.0195 | 78% |
| Acute kidney injury | Heterogeneous, assigned in turn | Mean | 5 | 0.246 | +0.0198 | 79% |
| Acute kidney injury | Heterogeneous, assigned at random | Mean | 5 | 0.230 | +0.0189 | 79% |
| 168 h mortality | Homogeneous, cosine | Mean | 1 | 0.605 | +0.0234 | 80% |
| 168 h mortality | Homogeneous, forest proximity | Mean | 1 | 0.465 | +0.0213 | 75% |
| 168 h mortality | Heterogeneous, assigned in turn | Mean | 5 | 0.391 | +0.0223 | 80% |
| 168 h mortality | Heterogeneous, assigned at random | Mean | 5 | 0.398 | +0.0242 | 78% |

**Supplementary Table S24.**
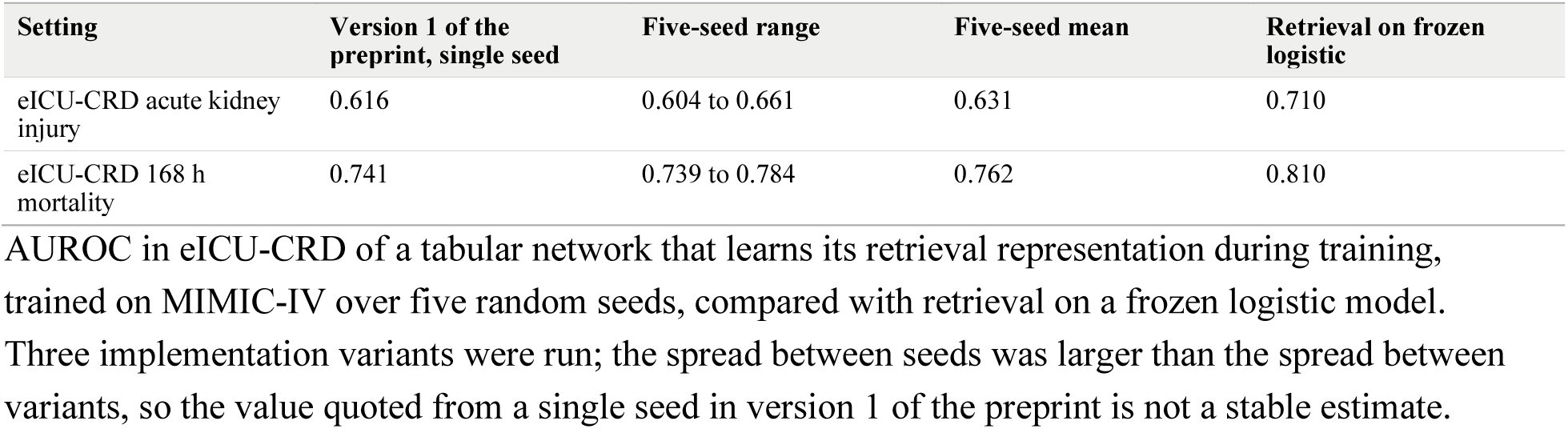
Five-seed retrieval-based network baseline.

| Setting | Version 1 of the preprint, single seed | Five-seed range | Five-seed mean | Retrieval on frozen logistic |
| --- | --- | --- | --- | --- |
| eICU-CRD acute kidney injury | 0.616 | 0.604 to 0.661 | 0.631 | 0.710 |
| eICU-CRD 168 h mortality | 0.741 | 0.739 to 0.784 | 0.762 | 0.810 |

AUROC in eICU-CRD of a tabular network that learns its retrieval representation during training, trained on MIMIC-IV over five random seeds, compared with retrieval on a frozen logistic model. Three implementation variants were run; the spread between seeds was larger than the spread between variants, so the value quoted from a single seed in version 1 of the preprint is not a stable estimate.

**Supplementary Table S25.**
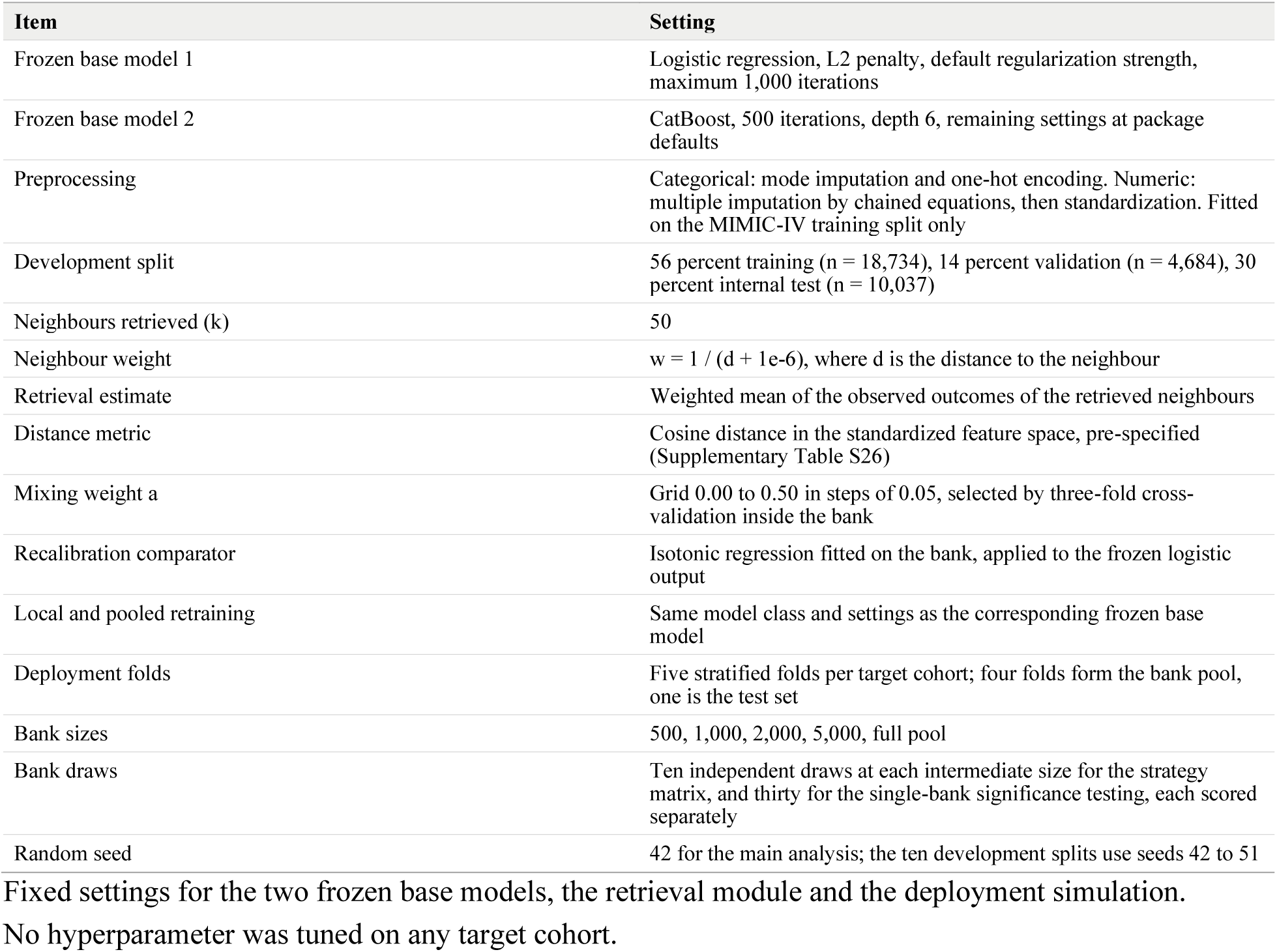
Model and retrieval settings.

| Item | Setting |
| --- | --- |
| Frozen base model 1 | Logistic regression, L2 penalty, default regularization strength, maximum 1,000 iterations |
| Frozen base model 2 | CatBoost, 500 iterations, depth 6, remaining settings at package defaults |
| Preprocessing | Categorical: mode imputation and one-hot encoding. Numeric: multiple imputation by chained equations, then standardization. Fitted on the MIMIC-IV training split only |
| Development split | 56 percent training (n = 18,734), 14 percent validation (n = 4,684), 30 percent internal test (n = 10,037) |
| Neighbours retrieved (k) | 50 |
| Neighbour weight | $w = 1 / (d + 1e-6)$, where d is the distance to the neighbour |
| Retrieval estimate | Weighted mean of the observed outcomes of the retrieved neighbours |
| Distance metric | Cosine distance in the standardized feature space, pre-specified (Supplementary Table S26) |
| Mixing weight a | Grid 0.00 to 0.50 in steps of 0.05, selected by three-fold cross-validation inside the bank |
| Recalibration comparator | Isotonic regression fitted on the bank, applied to the frozen logistic output |
| Local and pooled retraining | Same model class and settings as the corresponding frozen base model |
| Deployment folds | Five stratified folds per target cohort; four folds form the bank pool, one is the test set |
| Bank sizes | 500, 1,000, 2,000, 5,000, full pool |
| Bank draws | Ten independent draws at each intermediate size for the strategy matrix, and thirty for the single-bank significance testing, each scored separately |
| Random seed | 42 for the main analysis; the ten development splits use seeds 42 to 51 |

Fixed settings for the two frozen base models, the retrieval module and the deployment simulation.
No hyperparameter was tuned on any target cohort.

**Supplementary Table S26.** Retrieval distance metrics and stability of mutual information weights.

Panel A. Mean change in AUROC over ten development splits, frozen logistic model
| Outcome | R1 cosine | R2 mutual information weighted Euclidean | R3 mutual information random subspace | Between-split SD |
| --- | --- | --- | --- | --- |
| Acute kidney injury | 0.0059 | 0.0048 | 0.0054 | 0.0014 to 0.0023 |
| 168 h mortality | 0.0040 | 0.0049 | 0.0033 | 0.0021 to 0.0029 |

Panel B. Metric with the highest validation AUROC, in the first five of the ten splits
| Split seed | 42 | 43 | 44 | 45 | 46 |
| --- | --- | --- | --- | --- | --- |
| Acute kidney injury | R3 | R1 | R2 | R1 | R3 |
| 168 h mortality | R3 | R2 | R1 | R2 | R1 |

Panel C. Spearman correlation of bank-estimated with cohort-estimated mutual information
| Bank size | 50 | 100 | 300 | 500 | 1,000 | 2,000 | 5,000 |
| --- | --- | --- | --- | --- | --- | --- | --- |
| Acute kidney injury | 0.07 | 0.03 | 0.10 | 0.29 | 0.25 | 0.47 | 0.67 |
| 168 h mortality | -0.02 | 0.11 | 0.07 | 0.30 | 0.38 | 0.50 | 0.68 |

Correlation between the mutual information ranking of the 42 features estimated inside a bank of the stated size and the ranking estimated on the whole cohort. At a bank of 300 patients an average of 3.0 to 3.3 of the top ten features agreed with the top ten estimated on the whole cohort, so the two metrics that use mutual information have no basis at small bank sizes. Cosine distance was therefore pre-specified for all deployment analyses, and the neighbour weight was checked for instability: no neighbour was at zero distance in any setting, the median share of the single nearest neighbour in the total weight was 0.024 to 0.030, and its maximum was 0.11.

**Supplementary Note 1. TRIPOD+AI checklist**

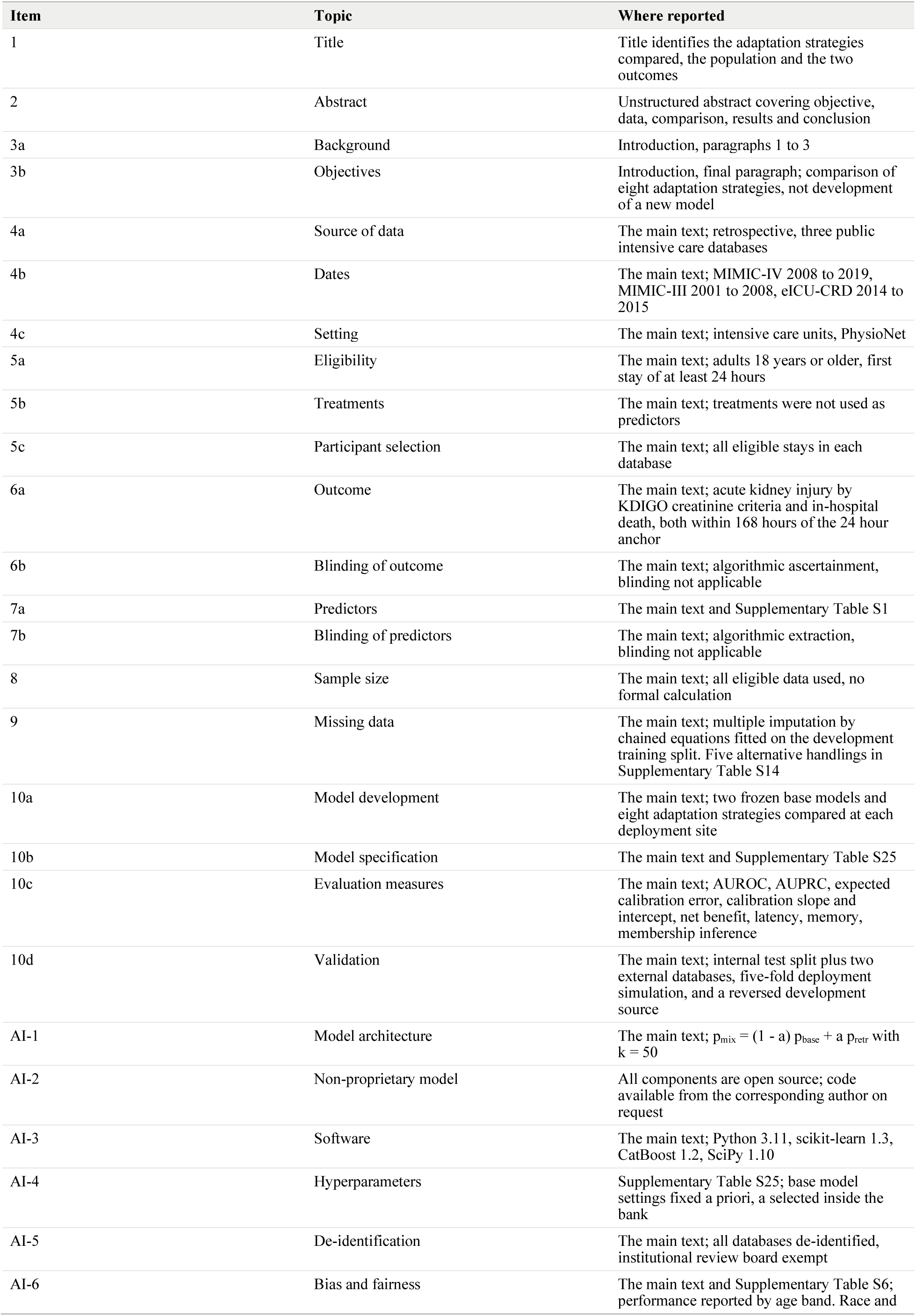
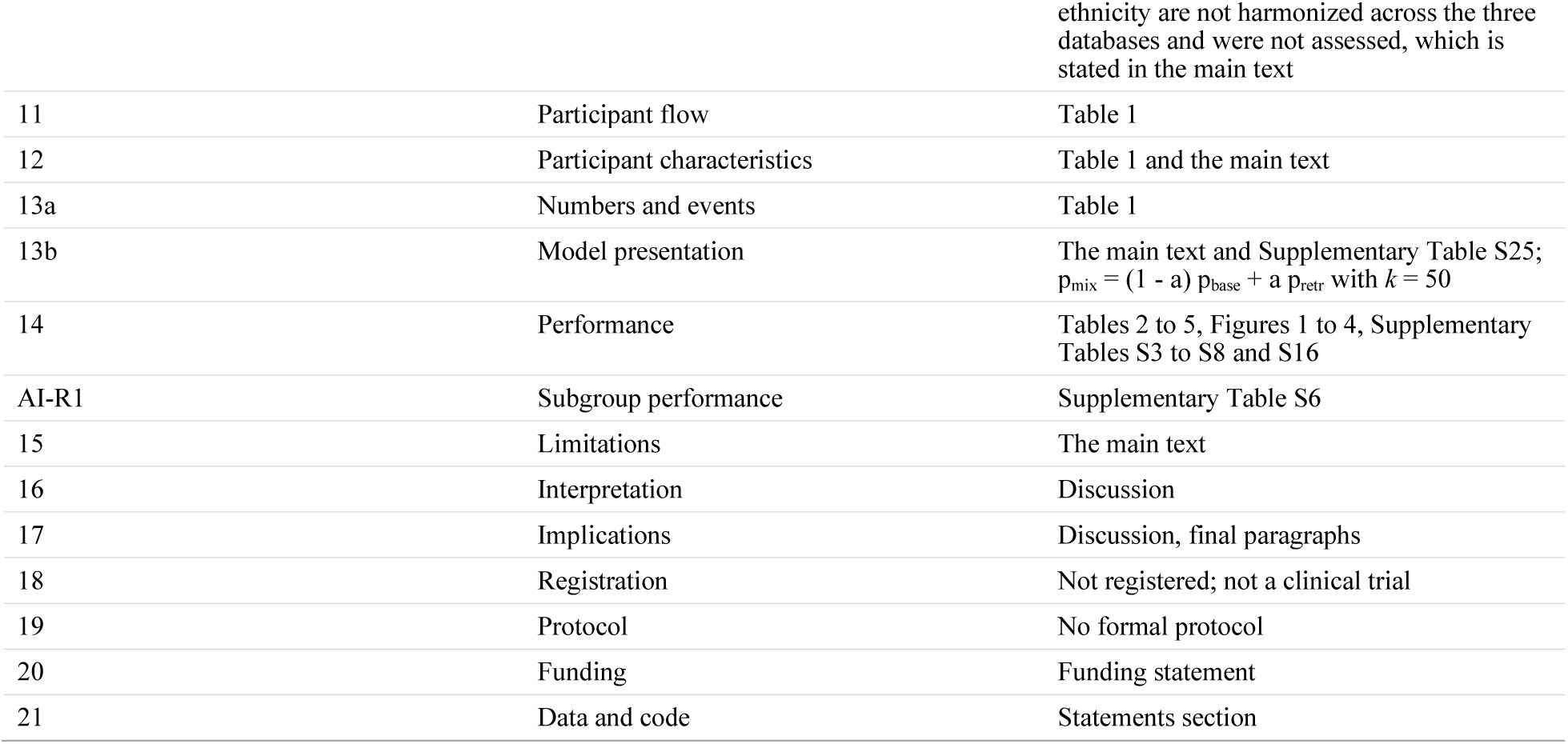

## References

1. Rajkomar, A. et al. Scalable and accurate deep learning with electronic health records. NPJ digital medicine. 1, 18 (2018).

2. Tomašev, N. et al. A clinically applicable approach to continuous prediction of future acute kidney injury. Nature. 572, 116–119 (2019).

3. Cho, N.-J. et al. Machine Learning to Assist in Managing Acute Kidney Injury in General Wards: Multicenter Retrospective Study. Journal of Medical Internet Research. 27, e66568 (2025).

4. Subbaswamy, A., Schulam, P. & Saria, S. Preventing failures due to dataset shift: Learning predictive models that transport. in The 22nd International Conference on Artificial Intelligence and Statistics (3118–3127 (PMLR, 2019).

5. Finlayson, S. G. et al. The clinician and dataset shift in artificial intelligence. New England Journal of Medicine. 385, 283–286 (2021).

6. Yang, J., Soltan, A. A. & Clifton, D. A. Machine learning generalizability across healthcare settings: insights from multi-site COVID-19 screening. NPJ digital medicine. 5, 69 (2022).

7. Rockenschaub, P. et al. The impact of multi-institution datasets on the generalizability of machine learning prediction models in the ICU. Critical Care Medicine. 52, 1710–1721 (2024).

8. Guo, L. L. et al. Evaluation of domain generalization and adaptation on improving model robustness to temporal dataset shift in clinical medicine. Scientific reports. 12, 2726 (2022).

9. Jeong, I., Cho, N.-J., Ahn, S.-J., Lee, H. & Gil, H.-W. Machine learning approaches toward an understanding of acute kidney injury: current trends and future directions. The Korean Journal of Internal Medicine. 39, 882 (2024).

10. Rosen, K. L., Sui, M., Ong, A. Y. & Kvedar, J. C. The portability paradox of foundation models for clinical decision support. NPJ Digital Medicine. 9, 285 (2026).

11. Van De Water, R. et al. Yet another ICU benchmark: A flexible multi-center framework for clinical ML. in International Conference on Learning Representations (44638–44693 (2024).

12. Wardi, G. et al. Predicting progression to septic shock in the emergency department using an externally generalizable machine-learning algorithm. Annals of emergency medicine. 77, 395–406 (2021).

13. Brown, N. A., Carey, C. H. & Gerry, E. I. FDA releases action plan for artificial intelligence/machine learning-enabled software as a medical device. *The Journal of Robotics*, Artificial Intelligence & Law. 4 (2021).

14. Gerke, S., Babic, B., Evgeniou, T. & Cohen, I. G. The need for a system view to regulate artificial intelligence/machine learning-based software as medical device. NPJ digital medicine. 3, 53 (2020).

15. Administration, U. S. F. a. D. Artificial Intelligence/Machine Learning (AI/ML)-Based Software as a Medical Device (SaMD) Action Plan. ((U.S. Food and Drug Administration, 2021).

16. Gulrajani, I. & Lopez-Paz, D. In search of lost domain generalization. arXiv preprint arXiv:2007.01434. (2020).

17. Koh, P. W., et al. Wilds: A benchmark of in-the-wild distribution shifts. in International conference on machine learning (5637–5664 (PMLR, 2021).

18. Zhang, T., Chen, M. & Bui, A. A. AdaDiag: Adversarial domain adaptation of diagnostic prediction with clinical event sequences. Journal of biomedical informatics. 134, 104168 (2022).

19. Jeong, I., Kim, Y., Cho, N.-J., Gil, H.-W. & Lee, H. A novel method for medical predictive models in small data using out-of-distribution data and transfer learning. Mathematics. 12, 237 (2024).

20. Gupta, P., Malhotra, P., Narwariya, J., Vig, L. & Shroff, G. Transfer learning for clinical time series analysis using deep neural networks. Journal of Healthcare Informatics Research. 4, 112–137 (2020).

21. Londschien, M., Burger, M., Rätsch, G. & Bühlmann, P. Domain generalization and adaptation in intensive care with anchor regression. RSS: Data Science and Artificial Intelligence. udag001 (2026).

22. Lewis, P. et al. Retrieval-augmented generation for knowledge-intensive nlp tasks. Advances in neural information processing systems. 33, 9459–9474 (2020).

23. Khandelwal, U., Levy, O., Jurafsky, D., Zettlemoyer, L. & Lewis, M. Generalization through memorization: Nearest neighbor language models. arXiv preprint arXiv:1911.00172. (2019).

24. Khandelwal, U., Fan, A., Jurafsky, D., Zettlemoyer, L. & Lewis, M. Nearest neighbor machine translation. arXiv preprint arXiv:2010.00710. (2020).

25. Zheng, X. et al. Adaptive nearest neighbor machine translation. in Proceedings of the 59th Annual Meeting of the Association for Computational Linguistics and the 11th International Joint Conference on Natural Language Processing (Volume 2: Short Papers) (368–374 (2021).

26. Gorishniy, Y. et al. Tabr: Tabular deep learning meets nearest neighbors. in International Conference on Learning Representations (18209–18249 (2024).

27. Ye, H.-J., Yin, H.-H., Zhan, D.-C. & Chao, W.-L. Revisiting nearest neighbor for tabular data: A deep tabular baseline two decades later. arXiv preprint arXiv*:2407.03257*. (2024).

28. Xu, R. et al. Ram-ehr: Retrieval augmentation meets clinical predictions on electronic health records. in Proceedings of the 62nd Annual Meeting of the Association for Computational Linguistics (Volume 2: Short Papers) (754–765 (2024).

29. Van Calster, B., McLernon, D. J., Van Smeden, M., Wynants, L. & Steyerberg, E. W. Calibration: the Achilles heel of predictive analytics. BMC medicine. 17, 230 (2019).

30. Vickers, A. J. & Elkin, E. B. Decision curve analysis: a novel method for evaluating prediction models. Medical Decision Making. 26, 565–574 (2006).

31. Lee, K. H., Yoon, D., Lim, H., Lee, K.-B. & Lee, Y. K. Deep learning models for acute kidney injury prediction: multi-center external validation and evaluation under simulated continuous monitoring conditions. npj Digital Medicine. (2026).

32. Rieke, N. et al. The future of digital health with federated learning. NPJ digital medicine. 3, 119 (2020).

33. Cao, J. et al. Exploring the limits of localization: federated model stacking improves hospital-level prediction in a national research network. npj Digital Medicine. (2026).

34. Rubachev, I., Kartashev, N., Gorishniy, Y. & Babenko, A. Tabred: Analyzing pitfalls and filling the gaps in tabular deep learning benchmarks. arXiv preprint arXiv*:2406.19380*. (2024).

35. Cai, H.-R. & Ye, H.-J. Understanding the limits of deep tabular methods with temporal shift. arXiv preprint arXiv:2502.20260. (2025).

36. Hripcsak, G. et al. Observational Health Data Sciences and Informatics (OHDSI): opportunities for observational researchers. Studies in health technology and informatics. 216, 574 (2015).

37. Bonawitz, K. et al. Practical secure aggregation for privacy-preserving machine learning. in proceedings of the 2017 ACM SIGSAC Conference on Computer and Communications Security (1175–1191 (2017).

38. Lundberg, S. M. & Lee, S.-I. A unified approach to interpreting model predictions. Advances in neural information processing systems. 30 (2017).

39. Ribeiro, M. T., Singh, S. & Guestrin, C. “ Why should i trust you?” Explaining the predictions of any classifier. in Proceedings of the 22nd ACM SIGKDD international conference on knowledge discovery and data mining (1135–1144 (2016).

40. Pelaccia, T., Tardif, J., Triby, E. & Charlin, B. An analysis of clinical reasoning through a recent and comprehensive approach: the dual-process theory. Medical education online. 16, 5890 (2011).

41. Rawson, T. M. et al. A real-world evaluation of a case-based reasoning algorithm to support antimicrobial prescribing decisions in acute care. Clinical Infectious Diseases. 72, 2103–2111 (2021).

42. Hájek, A. The reference class problem is your problem too. Synthese. 156, 563–585 (2007).

43. Breiman, L. Random forests. Machine learning. 45, 5–32 (2001).

44. Goldberger, A. L., et al. PhysioBank, PhysioToolkit, and PhysioNet: components of a new research resource for complex physiologic signals. circulation. 101, e215–e220 (2000).

45. Johnson, A. E. et al. MIMIC-IV, a freely accessible electronic health record dataset. Scientific data. 10, 1 (2023).

46. Johnson, A. E. et al. MIMIC-III, a freely accessible critical care database. Scientific data. 3, 1–9 (2016).

47. Pollard, T. J. et al. The eICU Collaborative Research Database, a freely available multi-center database for critical care research. Scientific data. 5, 180178 (2018).

48. Hoste, E. A. et al. Global epidemiology and outcomes of acute kidney injury. Nature Reviews Nephrology. 14, 607–625 (2018).

49. Mehta, R. et al. Diagnosis, evaluation, and management of acute kidney injury: A KDIGO summary (Part 1). (2013).

50. Van Buuren, S. & Groothuis-Oudshoorn, K. mice: Multivariate imputation by chained equations in R. Journal of statistical software. 45, 1–67 (2011).

51. Prokhorenkova, L., Gusev, G., Vorobev, A., Dorogush, A. V. & Gulin, A. CatBoost: unbiased boosting with categorical features. Advances in neural information processing systems. 31 (2018).

52. Higgins, J. P., Thompson, S. G., Deeks, J. J. & Altman, D. G. Measuring inconsistency in meta-analyses. bmj. 327, 557–560 (2003).

53. Dwork, C., McSherry, F., Nissim, K. & Smith, A. Calibrating noise to sensitivity in private data analysis. in Theory of cryptography conference (265–284 (Springer, 2006).

54. Shokri, R., Stronati, M., Song, C. & Shmatikov, V. Membership inference attacks against machine learning models. in 2017 IEEE symposium on security and privacy (SP) (3–18 (IEEE, 2017).

55. DeLong, E. R., DeLong, D. M. & Clarke-Pearson, D. L. Comparing the areas under two or more correlated receiver operating characteristic curves: a nonparametric approach. Biometrics. 837–845 (1988).

56. Benjamini, Y. & Hochberg, Y. Controlling the false discovery rate: a practical and powerful approach to multiple testing. Journal of the Royal statistical society: series B (Methodological). 57, 289–300 (1995).

57. Collins, G. S. et al. TRIPOD+ AI statement: updated guidance for reporting clinical prediction models that use regression or machine learning methods. bmj. 385 (2024).

